# Fertility decline in the later phase of the COVID-19 pandemic: The role of policy interventions, vaccination programmes, and economic uncertainty

**DOI:** 10.1101/2024.04.26.24306444

**Authors:** Maria Winkler-Dworak, Kryštof Zeman, Tomáš Sobotka

**Affiliations:** Vienna Institute of Demography (OeAW), Wittgenstein Centre for Demography and Global Human Capital (IIASA, OeAW, University of Vienna)

## Abstract

**BACKGROUND:** During the COVID-19 pandemic, birth rates in most higher-income countries first briefly declined and then shortly recovered, showing no common trends afterwards until early 2022, when they unexpectedly dropped.

**STUDY FOCUS:** We analyse monthly changes in total fertility rates in higher-income countries during the COVID-19 pandemic, with a special focus on 2022, when birth rates declined in most countries. We consider three broader sets of explanatory factors: economic uncertainty, policy interventions restricting mobility and social activities outside the home, and the role of vaccination programmes.

**STUDY DESIGN, DATA:** This study uses population-wide data on monthly total fertility rates adjusted for seasonality and calendar effects provided in the Human Fertility Database (HFD, 2023). Births taking place between November 2020 and October 2022 correspond to conceptions occurring between February 2020 and January 2022, i.e., after the onset of the pandemic but prior to the Russian invasion of Ukraine. The data cover 26 countries, including 21 countries in Europe, the United States, Canada, Israel, Japan and the Republic of Korea.

**METHODS:** First, we provide a descriptive analysis of the monthly changes in the total fertility rate (TFR). Second, we estimate the effects of the explanatory factors on the observed fertility swings using linear fixed effects (within) regression models.

**MAIN RESULTS:** We find that birth trends during the COVID-19 pandemic were associated with economic uncertainty, as measured by increased inflation, the stringency of pandemic policy interventions, and the progression of the COVID-19 vaccination campaign, whereas unemployment did not show any link to fertility during the pandemic.

**LIMITATIONS, REASONS FOR CAUTION:** Our research is restricted to higher-income countries with relatively strong social support policies provided by the government as well as wide access to modern contraception. Our data do not allow analysing fertility trends by key characteristics, such as age, birth order and social status.

**WIDER IMPLICATIONS OF THE FINDINGS:** This is the first multi-country study of the drivers of birth trends in a later phase of the COVID-19 pandemic. In the past, periods following epidemics and health crises were typically associated with a recovery in fertility. In contrast, our results show that the gradual phasing out of pandemic containment measures, allowing increased mobility and a return to more normal work and social life, contributed to declining birth rates in most countries. In addition, our analysis indicates that some women avoided pregnancy during the initial vaccination roll-out.

## Introduction

The COVID-19 pandemic of 2020–2022 contributed to distinct swings in birth rates. The initial shock was linked in most countries to a short-term decrease in the number of births around December 2020 to January 2021, followed by an equally brief recovery around March 2021 and a more differentiated development in the subsequent months that varied across countries (e.g., Bailey et al., 2023; Fallesen & Cozzani, 2023; Gietel-Basten & Chen, 2023; Gray et al., 2022; Kearney & Levine, 2023; Lappegård et al., 2023; Nisén et al., 2022; OECD, 2021a, 2021b; Plach et al., 2023; Sobotka et al., 2023). Some countries, including the Nordic countries, the Netherlands, Switzerland, Germany, Israel and the United States, even experienced a small “baby bump” during the second pandemic year, 2021. On balance, the changes in birth rates were smaller than initially expected when considering the unprecedented impact of COVID-19 and of the government responses to it on everyday lives, the labour market and social relations (Mayer, 2022; Settersten et al., 2020).

However, in the later phase of the pandemic, many higher-income countries experienced yet another shift in birth rates, an unexpected and robust downturn since early 2022 (Bujard & Andersson, 2024; Jasilioniene et al., 2024; Le Vu et al., 2023; Sobotka et al., 2023) that often persisted or even accelerated later in 2022 and in 2023 (HFD, 2023).

What could be the drivers of the unexpected decrease in births starting around January 2022? Going back nine months in time, to account for a typical length of pregnancy, we arrive in spring 2021, a time that can be considered a gradual “return to normality”. The disruptive impact of the pandemic diminished markedly in most countries. Lockdowns and social distancing measures were gradually phased out and were eventually lifted in 2022. As a consequence, people’s mobility and social contacts increased. Moreover, economic and labour market indicators had largely recovered from the initial pandemic shock. This return to normality was also achieved thanks to the COVID-19 vaccination programme, which was eventually becoming accessible to the whole population around mid-2021 in most countries. At the same time, inflation gradually increased in 2021 due to the increase in oil prices and the global supply chain crisis (supply and demand shocks brought on by the pandemic). Later, surging energy prices and wider uncertainty in the wake of the Russian invasion of Ukraine started fully affecting birth trends in late 2022. As a result of this renewed decline in birth rates, some countries reported record-low period fertility rates in 2022 and 2023 (e.g., Nordic Statistic Database, 2023).

While fertility trends during the early stages of the COVID-19 pandemic have been extensively researched (e.g., Bailey et al., 2023; Kearney & Levine, 2023; Plach et al., 2023; Sobotka et al., 2023), the majority of studies have focused on single countries and on conceptions in the first year of the pandemic. The most recent fertility declines and their drivers are not yet well documented and understood. Our study focuses on three sets of factors that can be empirically assessed and that may explain the unexpected drop in births starting around January 2022: (1) economic uncertainty, (2) phasing out of policy interventions restricting mobility and social contacts outside of the immediate family, and (3) the role of the vaccination programme.

There is ample evidence on the link between economic factors and fertility, where unemployment, inflation, and economic uncertainty mostly depress fertility rates (e.g., Comolli, 2017; Goldstein et al., 2013; Matysiak et al., 2021; Neels et al., 2024; Schneider, 2015; Sobotka et al., 2011). Economic uncertainty, including job disruptions and worries about unemployment and income loss jumped in the initial stage of the pandemic (OECD, 2021a, 2021b). In response, some women decided to delay or forego motherhood (Matsushima et al., 2023). Starting in (late) spring 2020, governments invested massively in job retention and income support schemes to mitigate the negative impact of the pandemic on the labour market, household income and economic output. In most of the higher-income countries, the unemployment rate returned to pre-pandemic levels by early 2021 and continued to decline until mid-2022. However, inflation rates started rising from mid-2021 during the period of economic recovery, which may have counteracted any fertility-enhancing response from economic recovery (OECD, 2023b).

To combat the spread of the virus and to support the economy, governments issued nonpharmaceutical policy interventions, such as work and school closures, travel restrictions, lockdowns, as well as income support and special subsidies for businesses affected by these interventions. These containment measures had led to major disruptions in social and family life (Mayer, 2022; OECD, 2021a, 2021b). They also resulted in increased stress and relationship struggles (Bellani & Vignoli, 2022)—factors that are negatively associated with an intention to become pregnant (Manning et al., 2022; Tasneem et al., 2023). At the same time, economic support cushioned financial pressure and economic uncertainty. Plach et al. (2023) found that containment measures led to a postponement of births, while economic support policies were positively associated with birth rates, but only in countries with low pre-pandemic social support policies (measured by public expenditures on family, health and unemployment support). The authors argued that pre-pandemic support policies broadly reflected the level of social trust, which might mitigate the negative consequences of pandemic-related uncertainty on fertility. However, their analysis was mostly based on data pertaining to the early phase of the pandemic. As the pandemic progressed, individuals developed coping strategies (Toffolutti et al., 2022), which may have altered the link between policy responses and fertility in the later part of the pandemic.

With limited opportunities for leisure, recreation and socialization, people started spending much more time at home with their partners and families. Working from home and saving commuting time to the workplace contributed to a better work-life balance. Opportunity costs of having a child declined. Under favourable circumstances, especially when feeling economically and socially secure, some couples might have rethought their priorities and decided to have (a)nother child or, more likely, have their next planned child earlier (Berrington et al., 2022; Neyer et al., 2022; Lappegård et al., 2023). Bujard and Andersson (2024) term this a “cocooning effect” (less poetically, Kearney and Levine (2023) referred to people who were “stuck home” with their romantic partners). As lockdowns and other restrictions gradually eased, especially after the COVID-19 vaccine became widely available in March– June 2021, people resumed work-related, leisure and socializing activities outside the home. In countries where COVID-19 containment measures resulted in fewer births, their ending would be expected to boost birth rates. In contrast, in countries where the “cocooning effect” contributed to rising birth rates, the end of pandemic-related restrictions would be expected to depress birth rates.

The decline in birth rates in early 2022 could also be linked to the first COVID-19 vaccination campaign in 2021. A large body of literature shows that COVID-19 vaccination does not lead to infertility problems among women or men or to increased adverse pregnancy outcomes (Chen et al., 2021; Wang et al., 2023; Zaçe et al., 2022); vaccination also does not increase the risk of miscarriage among pregnant women (Rimmer et al., 2023). However, it is likely that vaccination affected birth rates indirectly: couples might had decided to temporarily put pregnancy plans on hold during vaccination to reduce any potential harm to their foetus’s health (Bujard & Andersson, 2024). Such a decision would not be completely irrational—when vaccines were developed and introduced in late 2020, national health organisations and associations were hesitant to recommend vaccination during pregnancy until conclusive evidence was reached that COVID-19 vaccines are perfectly safe for pregnant women.^1^ Until early 2021, health authorities, including the US Centre for Disease Control and Prevention (CDC), adopted a cautious approach, suggesting that “pregnant women may choose to get any of the vaccines and should discuss risks and benefits with their healthcare providers” (CDC, 2021, p. 3). In mid-2021, only 22 out of 224 countries or territories recommended and 78 permitted (with qualifications) vaccination of pregnant women (Berman Institute of Bioethics & Center for Immunization Research, 2023). Most of the vaccines available in 2021 required two doses scheduled 3 to 12 weeks apart (or even longer intervals, especially when the supply of vaccines was still restricted) to complete the full course of vaccination^2^. Thus, some couples might have postponed their planned pregnancy for several months until finishing the full vaccination course.

In addition to these factors, fertility trends might have been affected by the dynamics of the COVID-19 pandemic itself. Periods of higher infection rates and excess mortality might be associated with depressed birth rates due to the worries women may have about becoming infected while pregnant, a desire to avoid hospitals and healthcare systems during infection peaks, or a desire to avoid possible exposure to COVID-19 during routine check-ups (Berrington et al., 2023). Some studies have shown a negative association between reported COVID-19 infections, deaths or overall excess deaths and birth rates, especially in the earlier phases of the pandemic (Kearney & Levine, 2023).

## Materials and methods

### Study population and period

This study uses population-wide data on monthly births and fertility rates collected in the Short-term Fertility Fluctuations (STFF) data series within the Human Fertility Database (HFD, 2023). The STFF data provide up-to-date information on live births by month of occurrence in selected countries with complete registration of births and available monthly reporting. Their monthly format makes the STFF data especially suitable for studying changes in fertility rates that may arise in response to sudden economic, political or pandemic shocks and changing policies, including the COVID-19 pandemic and the policies enacted to combat the spread of the virus. Our analysis focuses on birth trends from November 2020 to October 2022, covering conceptions occurring from February 2020 to January 2022. This period encompasses the onset of the pandemic but ends before the Russian invasion of Ukraine, which might have also affected fertility dynamics. The data cover 26 countries, including 21 countries in Europe, the United States, Canada, Israel, Japan and the Republic of Korea (hereafter South Korea). Table 1 provides an overview of the data, countries and regions covered. Among the data covered in the STFF data, we did not include Bulgaria, Lithuania or Russia due to missing data for some of the explanatory variables.

**Table 1:** Annual total fertility rates in 2019–2022, number of births, population size and trust in government before the pandemic by country.

| Country | TFR |  |  |  | Births<br>(thousand) | Population<br>(million) | Trust |
| --- | --- | --- | --- | --- | --- | --- | --- |
|  | 2019 | 2020 | 2021 | 2022 | 2022 | 2022 |  |
| Northern Europe |  |  |  |  |  |  |  |
| Denmark | 1.70 | 1.68 | 1.73 | 1.55 | 58.4 | 5.9 | higher |
| Finland | 1.35 | 1.37 | 1.46 | 1.32 | 45.0 | 5.6 | higher |
| Norway | 1.53 | 1.48 | 1.55 | 1.40 | 51.5 | 5.5 | higher |
| Sweden | 1.71 | 1.67 | 1.67 | 1.50 | 104.7 | 10.5 | higher |
| Western Europe |  |  |  |  |  |  |  |
| Belgium | 1.60 | 1.57 | 1.60 | 1.54 | 113.6 | 11.6 | lower |
| France | 1.83 | 1.79 | 1.80 | 1.76 | 686.9 | 65.5 | lower |
| Ireland | 1.71 | 1.66 | 1.72 | 1.60 | 54.4 | 5.0 | lower |
| Netherlands | 1.57 | 1.55 | 1.62 | 1.51 | 167.5 | 17.5 | higher |
| United Kingdom | 1.64 | 1.57 | 1.53 | 1.50 | 672.6 | 66.9 | lower |
| German-speaking countries |  |  |  |  |  |  |  |
| Austria | 1.46 | 1.44 | 1.48 | 1.41 | 82.6 | 9.0 | lower |
| Germany | 1.54 | 1.53 | 1.58 | 1.47 | 738.8 | 82.8 | higher |
| Switzerland | 1.48 | 1.47 | 1.52 | 1.39 | 82.4 | 8.7 | higher |
| Central-Eastern Europe |  |  |  |  |  |  |  |
| Czechia | 1.75 | 1.76 | 1.82 | 1.68 | 101.3 | 10.5 | lower |
| Hungary | 1.49 | 1.56 | 1.59 | 1.55 | 88.5 | 9.7 | lower |
| Latvia | 1.61 | 1.55 | 1.57 | 1.46 | 16.0 | 1.9 | lower |
| Poland | 1.41 | 1.38 | 1.33 | 1.25 | 305.1 | 38.3 | lower |
| Slovenia | 1.61 | 1.60 | 1.64 | 1.57 | 17.6 | 2.1 | lower |
| Southern Europe |  |  |  |  |  |  |  |
| Greece | 1.34 | 1.39 | 1.43 | 1.31 | 76.1 | 10.6 | lower |
| Italy | 1.27 | 1.25 | 1.25 | 1.24 | 392.6 | 58.7 | lower |
| Portugal | 1.43 | 1.41 | 1.34 | 1.45 | 83.7 | 10.3 | lower |
| Spain | 1.24 | 1.19 | 1.19 | 1.20 | 330.2 | 47.2 | lower |
| North America |  |  |  |  |  |  |  |
| Canada | 1.47 | 1.40 | 1.43 | 1.36 | 351.7 | 38.2 | higher |
| United States | 1.70 | 1.64 | 1.66 | 1.66 | 3665.0 | 332.0 | lower |
| East Asia |  |  |  |  |  |  |  |
| Japan | 1.34 | 1.33 | 1.30 | 1.25 | 770.8 | 122.3 | lower |
| South Korea | 0.92 | 0.84 | 0.81 | 0.78 | 249.2 | 51.2 | lower |
| Other regions |  |  |  |  |  |  |  |
| Israel | 3.01 | 2.90 | 3.00 | 2.93 | 181.2 | 9.5 | lower |

While the analysed data provide complete coverage of births, data for some countries might be affected by late reporting, reporting by month of registration rather than by month of birth, and different rules pertaining to the inclusion or exclusion of births by foreigners, refugees and asylum seekers residing in a country. Details about the birth data for individual countries are provided in the STFF Metadata document (HFD STFF, 2023).

### Indicators used: monthly total fertility rates

The analysis uses monthly series of total fertility rates (TFR) adjusted for both calendar^3^ and seasonal variation. The method of estimating the monthly TFRs from the absolute monthly number of births as well as the seasonal adjustment are explained in detail in the STFF Methodological Note (Jdanov et al., 2022).

The TFR is an indicator estimating the number of children that would be born per woman if age-specific fertility rates among women of reproductive age (15-49) remained the same indefinitely as in the observed period. Compared with the absolute number of births or crude birth rates, the TFR has the advantage of not being affected by changes in the size of the female population of reproductive age over time and can easily be compared across countries. However, the TFR may be affected by changes in the age at childbearing. When births are shifted to younger or older childbearing ages, the TFR does not properly reflect the ultimate family size among women at the end of their reproductive span (Bongaarts & Feeney, 1998; Sobotka & Lutz, 2011).

### Covariates

Explanatory variables include indicators reflecting economic uncertainty, nonpharmaceutical policy interventions (NPIs), as well as indicators of the vaccination rollout.

For economic uncertainty, we use the seasonally adjusted monthly harmonized unemployment rate (OECD, 2023d) as well as the monthly consumer price index, with 2015 as the base year (OECD, 2023a). For easier interpretation, we rescale the consumer price index relative to its value in the year 2019. We also considered other monthly economic indicators such as the consumer confidence index and the economic policy uncertainty index (Baker et al., 2016). However, these indicators were only available for all countries in this study.

We include two NPI indices—the stringency index and the economic support index—from the Oxford COVID-19 Government Response Tracker (Hale et al., 2021). The stringency index is a composite measure of nine policy responses: school closures, workplace closures, cancellation of public events, restrictions on public gatherings, closures of public transport, stay-at-home orders, restrictions on internal movement, international travel controls and public information campaigns. Each component is normalized to range from 0 to 100, and the index is computed as an average of the nine components. Higher values indicate more stringent measures, with 100 representing the most stringent response. Similarly, the economic support index is calculated as the average of the policy responses on income support as well as on debt relief and contract relief.

Building upon Plach et al. (2023) argument on the importance of social trust mediating the association between government policies and fertility, we differentiate countries according to the level of social trust using annual data on trust in government before the pandemic (2010-2019) from OECD (2023c). By employing partitioning around medoids methods (Kaufman, 1990),^4^ countries are grouped into those with lower and higher trust in government (see Table 1).^5^

Vaccination programmes in most countries were age-graded, with the oldest population being the first to receive the vaccine. Women and men of reproductive age became eligible several months later, usually in spring or early summer 2021 (Hale et al., 2021). Most vaccines required two doses to complete the primary protocol. To account for the potential pregnancy postponement during the initial COVID-19 vaccination programme, we use data on both the share of the population that received at least one dose and the share of the population who completed the initial COVID-19 vaccination protocol over time. If some women indeed decided to avoid or postpone pregnancies around the time of their vaccination, the former indicator, representing ongoing vaccination, should have a negative effect on fertility, while the latter, representing the completion of vaccination, should have a positive effect. The data come from Our World in Data (Mathieu et al., 2020), which collates and processes the up-to-date official vaccination data on a daily or weekly basis for the total population. We use interpolation techniques to derive the mid-month value. For an additional analysis, we use data available for broader age groups and select the share of the population having received the first dose and completing the primary course of vaccination, respectively, in the prime childbearing age group (25-49 years). These data are not available for four out of the 26 countries. Israel, Canada, and Switzerland use different age groups; we selected a narrower age group, specifically 30–39 years, to represent prime childbearing ages.

### Control variables

In addition, we control for the health emergency during the pandemic. We use excess mortality as reported by Our World in Data (Mathieu et al., 2020). Excess mortality is measured using a p-score, which corresponds to the relative difference between the reported number of deaths (HMD, 2023) and the projected (based on pre-pandemic trends) number of deaths from all causes (Karlinsky & Kobak, 2021). Data on excess mortality are provided on a monthly or weekly basis, which are in the latter case converted into monthly averages. In addition, we include a dummy variable in our statistical analysis for the first wave of the COVID-19 pandemic, running from February to April 2020, reflecting the high level of uncertainty just after the start of the pandemic.

Supplemental Figure S1. depicts trends in the covariates together with the seasonally adjusted monthly total fertility rate for all countries analysed.

### Statistical methods and models

We first analyse fertility trends in 26 high-income countries since the onset of the COVID-19 pandemic, documenting the accelerated fertility decline in many countries since early 2022. Next, we assess the correlations of the individual explanatory variables with the seasonally adjusted monthly TFR, shifted by nine months to the approximate time of conception, in each country. Then, we adopt a multivariable approach by estimating a linear fixed effects regression model of the relationship between the TFR and the explanatory variables.

The baseline model includes the indicators measuring economic uncertainty (the unemployment rate and the consumer price index), nonpharmaceutical policy interventions (the stringency index and the economic support index), vaccination rollout (the cumulative share of the population that received the first dose and that completed the initial vaccination protocol for the vaccination rollout), and controls for pandemic severity and stage (using excess mortality and a dummy variable for the first COVID-19 wave).

Next, we add the stringency index lagged by one month to allow for later fertility adjustments associated with the containment measures. For instance, a negative coefficient of the (non-lagged) stringency index in conjunction with a positive coefficient of the lagged stringency index would suggest that fertility was negatively associated nine months after restrictions were in place but that births were subsequently partly recovered one month later. Moreover, we examine whether the association between the NPI and fertility differs across countries depending on the level of social trust and whether these associations have changed over the course of the pandemic.

We estimate the fixed effects regression models by adopting Driscoll and Kraay (1998) standard errors, which are robust to disturbances being heteroskedastic, autocorrelated, and cross-sectional dependent (Hoechle, 2007).^6^ A p-value less than 0.05 was considered to indicate statistical significance. While the data and the descriptive analysis were carried out in R (R Core Team, 2023), Stata was used for the regression analysis (StataCorp, 2023).

## Results

Figure 1 depicts the monthly trends in the Total Fertility Rate for all countries analysed by both month of birth and the estimated month of conception. The country TFRs are shown relative to those in November 2021 to enhance comparability and to demonstrate the extent of fertility changes in early 2022 (absolute TFRs are depicted in Supplementary Figure S1). The figure reveals variation in fertility trends across countries and broader regions, with common periods of ups and downs. These include especially the initial pandemic dip in fertility around December 2020 and January 2021 (with conceptions around April 2020), a brief recovery two months later, and a decrease in fertility since early 2022. This recent decrease in fertility was pronounced in the Nordic countries, Western Europe (except in the United Kingdom), in German-speaking countries (Austria, Germany, Switzerland), and in Czechia, Hungary, Slovenia, Poland, Latvia and Greece. In contrast, countries outside Europe (Japan, South Korea, Canada, the United States and Israel) and Southern European countries (except for Greece) did not experience any sustained downward trend in fertility since early 2022.

**Figure 1:**
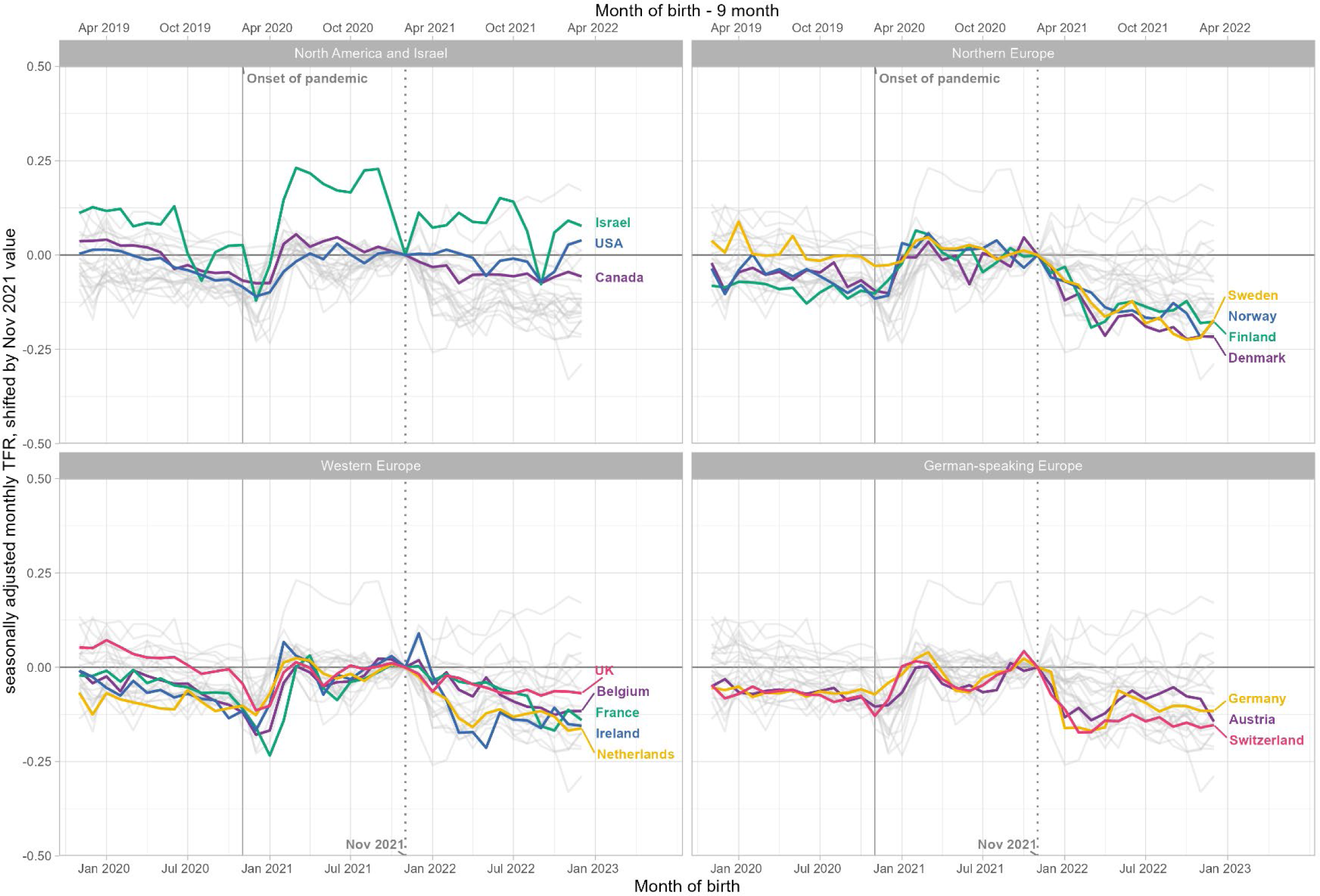
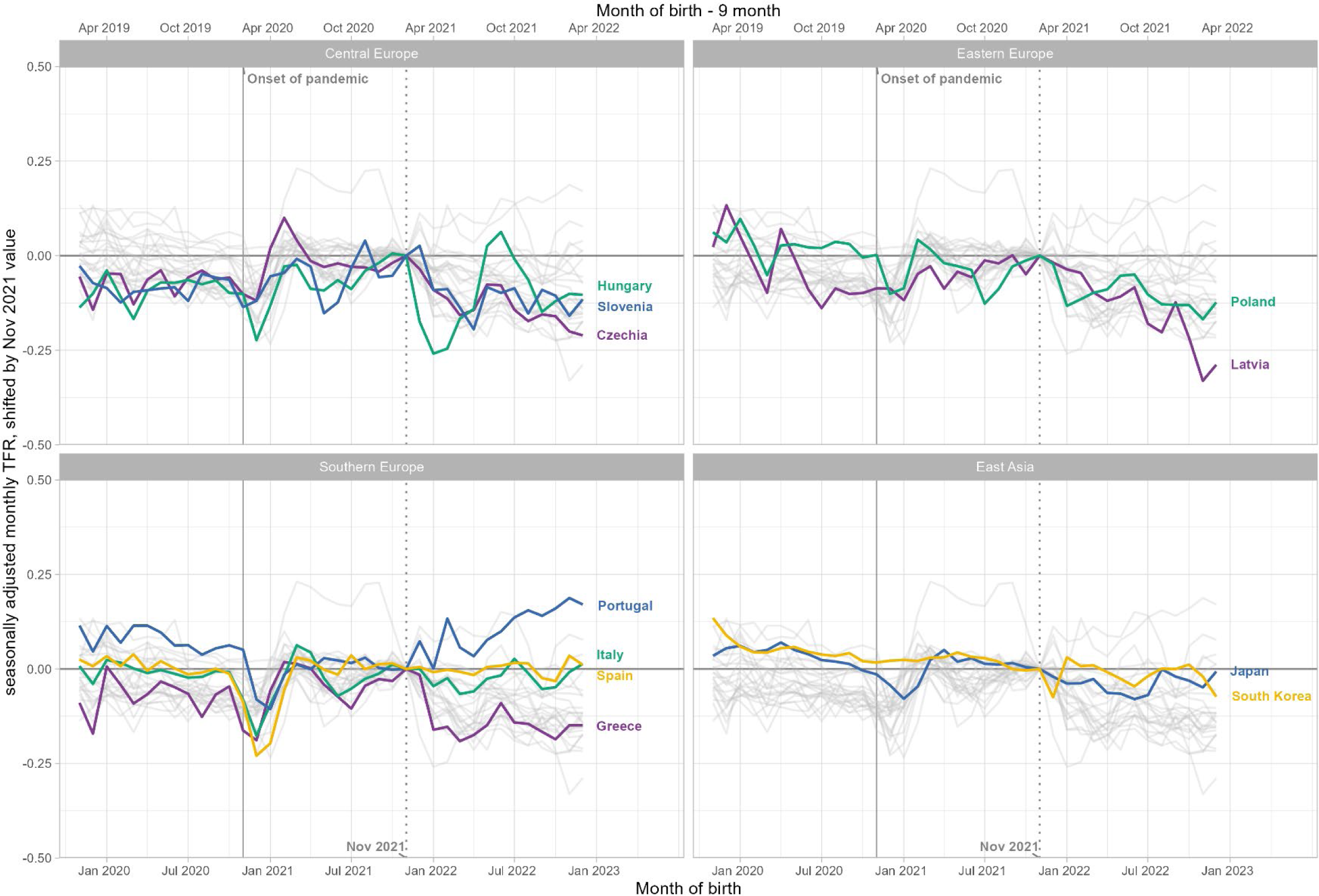
Seasonally adjusted monthly TFRs relative to the November 2021 level (Nov 2021 = 0), November 2019 to December 2022.

Fertility decline was especially sharp in the first months of 2022 (in a few countries it started already in December 2021, and in Israel in October 2021), often with a temporary recovery (e.g., in Hungary, Germany, Austria, Ireland, Denmark, Finland, Greece, and Poland) around June 2022. This was mostly followed by a renewed, albeit more gradual, fertility decline since August 2022. For instance, the estimated seasonally adjusted TFR in Germany decreased by approximately 10%, from 1.58 in December 2021 to 1.42-1.43 in January-April 2022, and then recovered to values at or above 1.50 in May-July 2022 before declining slightly again. Across all analysed countries, the TFR decreased on average by 0.10 between November 2021 and April 2022—a relative decline by 6%.

Figure 2 summarizes the correlation of the individual explanatory variables with the monthly seasonally adjusted TFR, where countries are clustered by geographical region. The colour and the angle of the ellipses indicate the direction of the correlation: purple and left-rotated ellipses represent negative correlations, and green and right-rotated ellipses denote positive correlations. The darker the colour and the “thinner” the shape of the ellipse are, the stronger the correlation. The pale solid circles indicate that the series of the respective explanatory variable is unrelated to the TFR time series during the pandemic.

**Figure 2:**
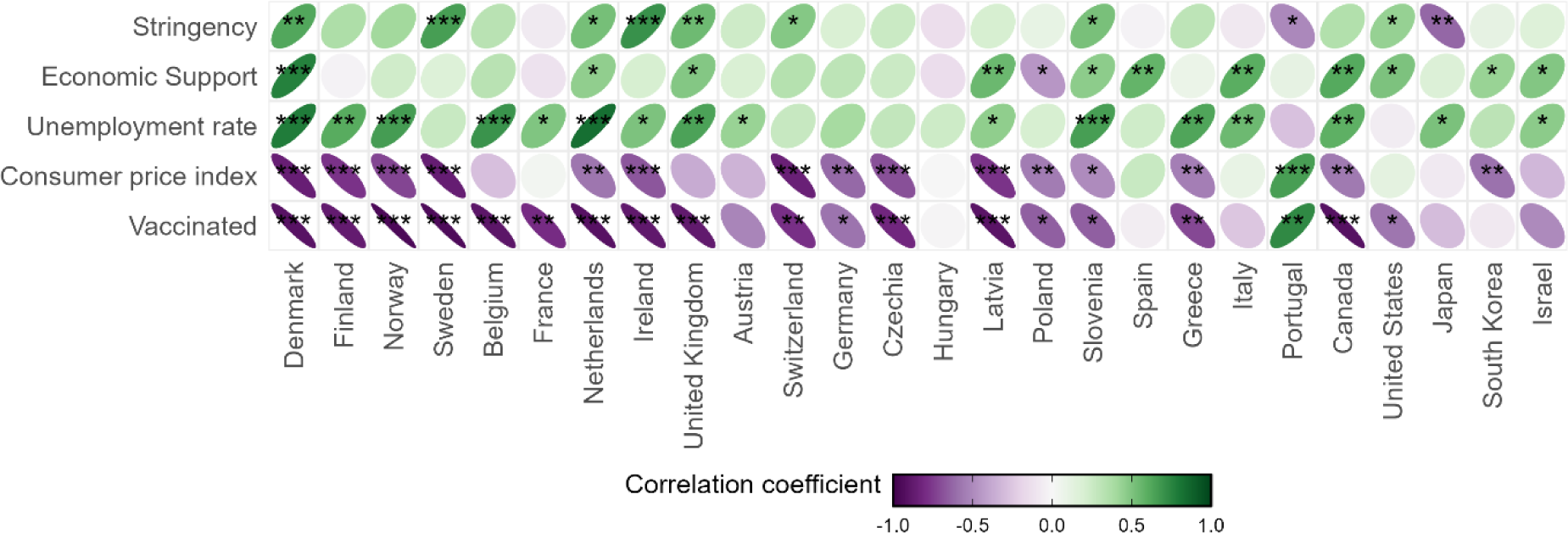
Summary of correlation coefficients of explanatory variables with the monthly seasonally adjusted TFR by country. Note: Own computations, visualization adapted from Cai et al. (2022). * p<0.05, ** p<0.01, *** p<0.001.

We find that the stringency index is positively associated with birth trends in most countries. The exceptions to this pattern are Japan and Portugal, where fertility was significantly lower during stricter containment measures. This suggests that the decline in the stringency of pandemic containment measures and the associated increase in mobility were associated with a decline in fertility.

Governmental economic support aimed to cushion economic uncertainty among the population and, accordingly, is positively correlated with fertility during the pandemic. Strikingly, the unemployment rate is also positively associated with birth trends in almost all countries, challenging well-established findings about the negative impact of unemployment on fertility. In line with past research, the consumer price index is strongly negatively correlated with childbearing in almost all countries.

Finally, vaccination rollout is also strongly negatively correlated with fertility in almost all countries, with the most notable exception being Portugal, where fertility increased along with vaccination take-up.

Figure 3 plots the estimated model coefficients of the fixed effects regression model of the seasonally adjusted TFR per 100 women (see also supplementary Table S1). The model includes economic indicators, two nonpharmaceutical policy intervention indicators and indicators of vaccination rollout while additionally controlling for health emergency and the first COVID-19 wave. For the economic indicators, we do not find the expected negative association with the unemployment rate (β= −0.051, 95% CI: −0.300 to 0.200). However, due to massive government interventions, the unemployment rate might not properly reflect economic uncertainty during the pandemic (OECD, 2020). In only a few countries, including Canada and the United States, unemployment surged after the onset of the pandemic (e.g., Kearney & Levine, 2023). The negative impact of the consumer price index on birth trends is confirmed in the multivariable analysis (β= −0.600, 95% CI: −0.887 to −0.312). On average, consumer prices increased for all countries by approximately five percentage points in 2021, with the Czech Republic and Poland experiencing a 10 percentage point increase. Considering the average increase of five percentage points, higher inflation would be associated with a monthly decline in the TFR of three births per 100 women in the period of October 2021 to September 2022.

**Figure 3:**
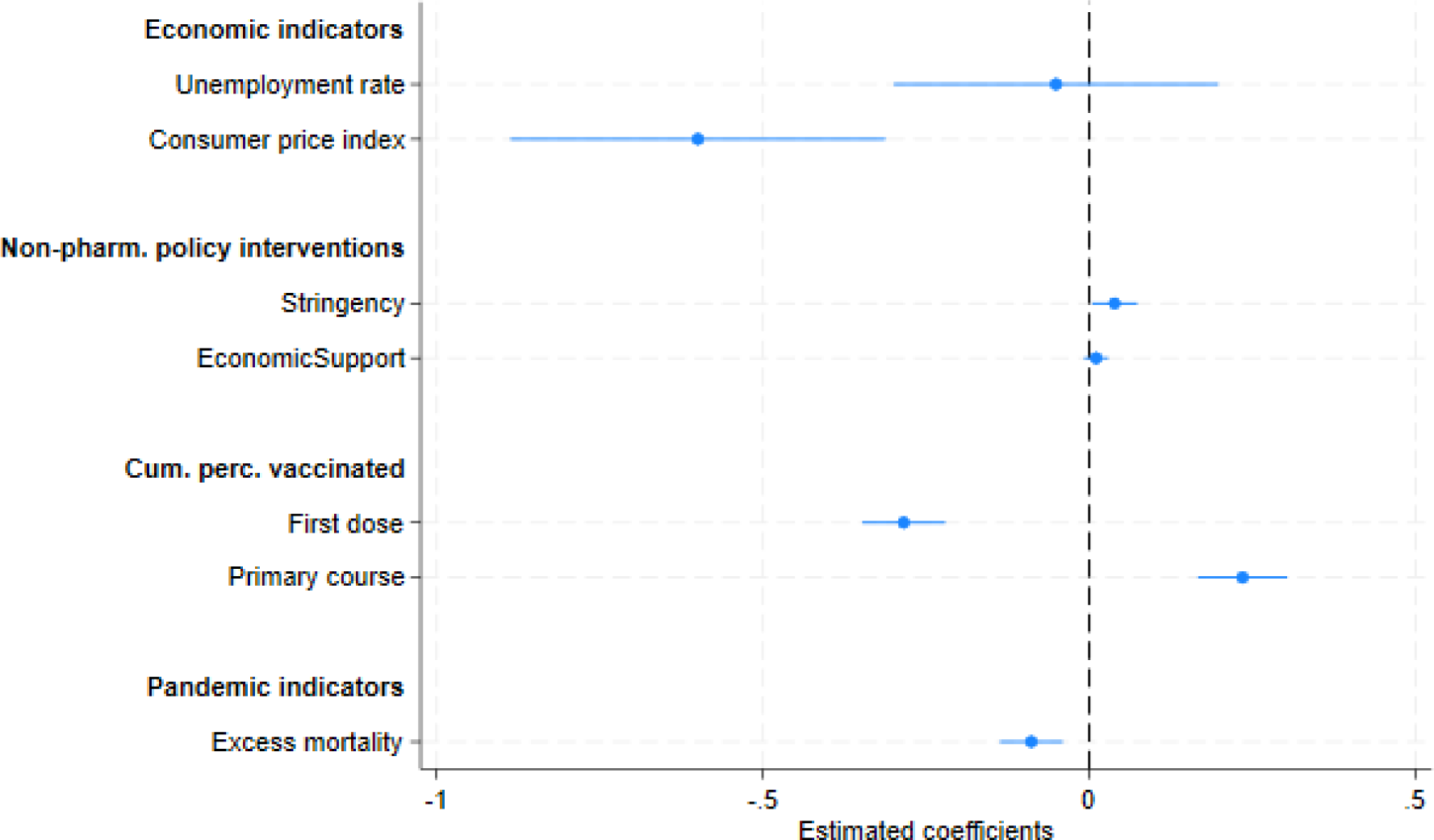
Estimated model coefficients of the fixed effect model of the monthly seasonally adjusted TFR per 100 women. Note: The regression model includes additional controls for the first COVID-19 wave and country fixed effects.

With regard to the nonpharmaceutical policy measures, we observe a small but statistically significant positive association between the stringency index and the total fertility rate (β= 0.039, 95% CI: 0.004 to 0.073), while economic support does not show the expected positive association (β= 0.011, 95% CI: −0.008 to 0.029). In an alternative model, the normalcy index developed by the Economist (2021) was included instead of the stringency index in the regression model (see also supplementary Table S2). The normalcy index was designed to reflect changes in population’s mobility and activities outside of the home.^7^ The estimated coefficient, is not significant (p=0.218). However, the normalcy index has several important caveats. It is relative to a pre-pandemic level, which is only measured in January and February 2020, and it may therefore be biased by seasonal variations in the indicators of the different domains. Furthermore, it is not available for all countries considered in this study.

Our results suggest that the age-graded, two-doses vaccination scheme resulted first in lower fertility rates and later was linked with their recovery. When the vaccination rollout gained momentum and the cumulative share of the population having received at least one dose of the COVID-19 vaccine increased, fertility decreased (β= −0.260, 95% CI: −0.324 to −0.197). This suggests that some women chose to delay their pregnancies until after completing their vaccination. Indeed, we find a statistically significant positive association between the cumulative percentage of the population that has completed the primary vaccination course and the TFR (β= 0.213, 95% CI: 0.144 to 0.281). It is worth noting that the estimated coefficient is almost equal in magnitude as the estimated negative coefficient of the indicator signalling the start of the vaccination rollout. In the spring of 2021, the share of the population that had received the first dose of the vaccination increased on average by 12 percentage points per month. This would be associated with an average decline of approximately three births per 100 women per month in the period from January to March 2022. In early summer, the share of the population completing the initial vaccination protocol similarly increased. This would be then associated with a compensatory average increase in the TFR of approximately 2.6 births per 100 women per month in the period from March to May 2022. However, the timing and pace of vaccine roll-out varied markedly across countries. In fact, some countries experienced increases in the share of the population vaccinated that were more than double the average numbers stated above.

Due to the age-graded nature of the vaccination roll-out, women of childbearing age were entitled to the vaccination in late spring or early summer 2021 in most countries. For a subset of countries, vaccination data is also available by broad age groups. As a robustness check, we included the share of the population of childbearing age instead of the total population in the regression model. This revealed a similar pattern of fertility delay and recovery during the primary course of vaccination (see also supplementary Table S2).

Finally, the regression model included two controls for the health emergency and the first COVID-19 wave (see supplementary Table S1). The health crisis, as measured by excess mortality, is negatively associated with fertility (β= −0.089, 95% CI: −0.127 to −0.051). The estimated coefficient for the early pandemic months, February to April 2020, is −8.275 (95% CI: −9.721 to −6.829), which is consistent with the markedly depressed fertility nine months later, i.e., November 2020 to January 2021 (not shown in Figure 3 due to the large effect size).

Next, in Figure 4, we inspect the association of nonpharmaceutical policy interventions with fertility by first including the lagged stringency index and adding an interaction between the policy responses (stringency, lagged stringency and economic support) and the level of trust in government in the country (see also supplementary Table S1). In countries with lower levels of trust in government, stricter containment measures were associated with a statistically significant decrease in fertility (β= - 0.107, 95% CI: −0.200 to −0.015). The positive coefficient of the lagged stringency variable (β= 0.105, 95% CI: 0.013 to 0.198) suggests that births subsequently partly recovered. However, we do not find any evidence of such containment measure-associated fertility postponement in countries with higher institutional trust. To the contrary, periods of stricter containment measures in higher-trust countries were associated with a statistically significant higher total fertility rate (β= 0.140, 95% CI: 0.023 to 0.256) without any indication of a later “compensatory” decline (β= −0.008, 95% CI: −0.123 to 0.108). Conversely, a decline in the stringency of measures would be associated with depressed fertility again. For instance, a reduction in the stringency index of approximately 10 points in one month would be associated with a decline in fertility of 1.4 births per 100 women in higher-trust countries nine months later.

**Figure 4:**
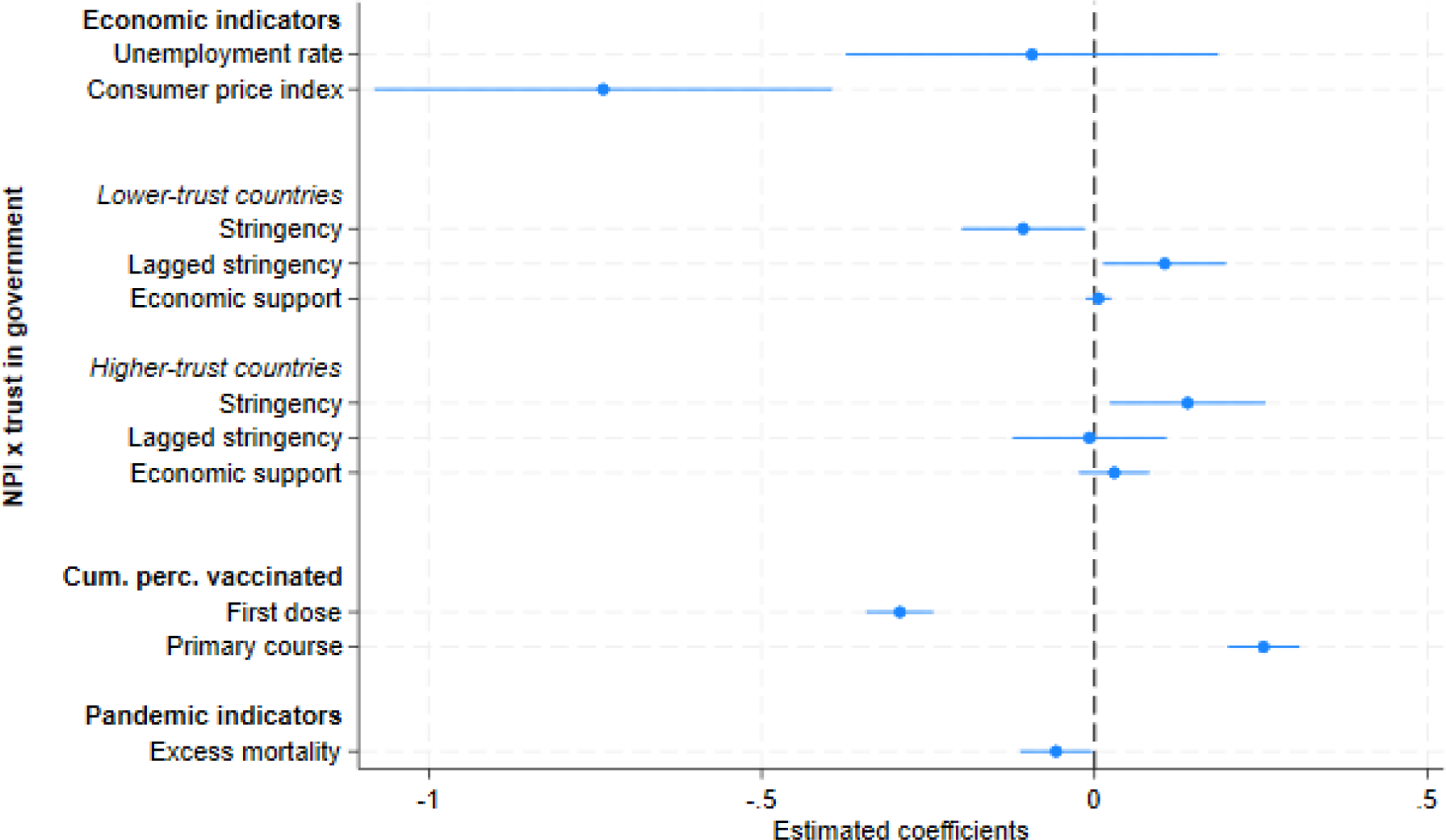
Estimated model coefficients of the fixed effect model of the monthly seasonally adjusted TFR per 100 women distinguishing stringency of containment measures in lower- and higher-trust countries. Note: The regression model additionally includes a time dummy for the first COVID-19 wave and country fixed effects.

Unlike the results for the stringency index, we do not find any visible country differences in the association between economic support and total fertility rates. In both country groups, the estimated effects are small and not significantly different from zero (β= 0.006, 95% CI: −0.013 to 0.198, and β= - 0.030, 95% CI: −0.023 to 0.083, in lower- and higher-trust countries, respectively). While economic support cushioned economic uncertainty and income loss, it is not linked to childbearing behaviour during the pandemic when simultaneously considering economic indicators, containment measures and vaccination rollout.

The vaccination rollout and the prospect of a return to normality may have altered the relationship between NPIs and fertility over the course of the pandemic. In a further analysis, we thus estimate the link between NPIs and fertility separately during the early and a later phase of the pandemic. Figure 5 displays the estimated coefficients for containment and economic support measures in lower- and higher-trust countries for two periods, February 2020 to December 2020, versus January 2021 to January 2022 (see also supplementary Table S1).^8^ We confirm the previously derived pattern of fertility decline and recovery in lower-trust countries, but only for the early phase of the pandemic. For conceptions from January 2021 to January 2022, we find no association between policy interventions and fertility in lower-trust countries. In higher-trust countries, the stringency index tends to be positively associated with fertility, but the statistical significance is weak (p=0.057).

**Figure 5:**
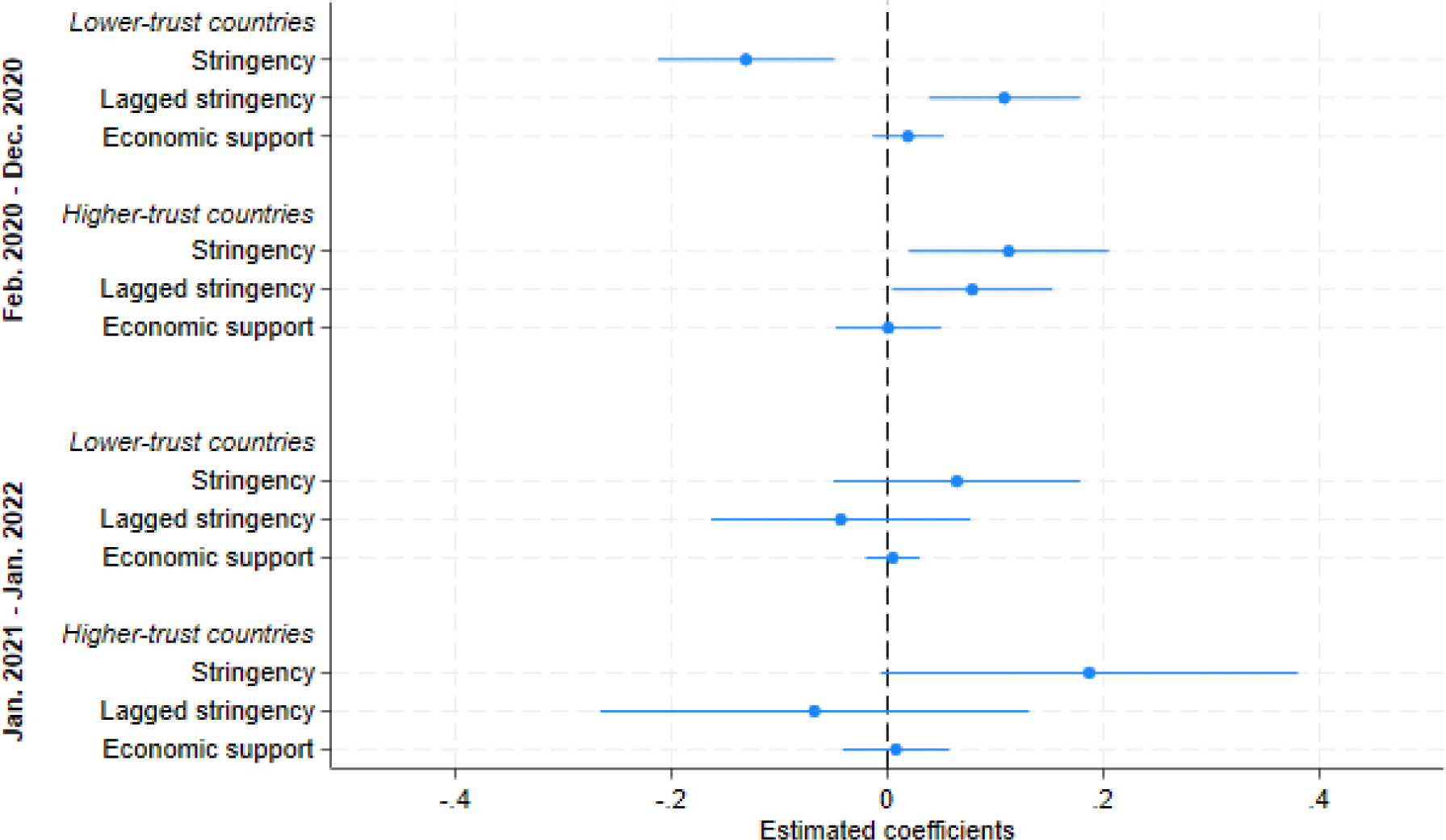
Estimated relationship of nonpharmaceutical policy interventions by level of trust and period of conception Note: The regression model also includes the covariates representing the economic indicators and the vaccination rollout indicators as well as the controls for the severity of the pandemic and country fixed effects.

## Discussion

The COVID-19 pandemic was associated with distinct short-term ups and downs in birth trends across low-fertility countries. This study sheds light on the factors driving these fluctuations. We focus especially on the later phase of the pandemic, when vaccination became widely available, lockdowns, school closures and social distancing measures gradually phased out, mobility and socialization outside of the home increased, and more people returned to their offices and workplaces and life partly returned to pre-pandemic “normality.” Unexpectedly, birth rates in many European countries dropped starting around January 2022, often putting an end to a minor pandemic era upturn in births during spring-autumn 2021. Our results suggest that economic uncertainty, nonpharmaceutical policy interventions and the first wave of the population-wide vaccination programme contributed to the decline in birth rates during 2022.

One measure of economic uncertainty, inflation, displayed a strong and significantly negative association with birth rates during the pandemic. The rate of inflation across higher-income countries typically decreased during the early phase of the pandemic in 2020 but then began to rise during 2021. The gradual but steady rise in inflation in 2021 thus contributed to the observed decline in birth rates. In 2022, inflation in many OECD countries was more than twice as high as that in 2021. We therefore expect inflation to have a negative and relatively strong impact on fertility beyond our study period, especially in 2023.

In contrast, the unemployment rate was not associated with birth trends during the COVID-19 pandemic. This unexpected finding could be partly explained by an anomalous labour market situation and policy interventions in the early stages of the pandemic. First, economic activity and unemployment rates in 2020-2021 were strongly affected by the government’s massive interventions to protect jobs and the economy. Most countries, therefore, did not experience strong swings in unemployment (the United States and Canada are the major exceptions among the countries analysed here). Second, government social and welfare policies helped reduce economic uncertainty among people potentially facing employment loss or unstable employment.

The association between containment and mobility restrictions on birth rates, as measured by the stringency index, differed between lower-trust countries and higher-trust countries. In countries with higher levels of trust, stricter policies were linked with higher birth rates, partly explaining the temporary upturn in births observed, for example, in the Nordic countries, Germany, and the Netherlands. For some couples, especially in countries with stronger economic and family policy support, greater trust in government and less disruptive impacts of COVID-19-related policies on everyday life, this was a favourable time to have children despite the pandemic (Bujard & Andersson, 2024; Lappegård et al., 2023; Nisén et al., 2022). In spring 2021, the stringency of the containment measures was gradually reduced, putting an end to the previously observed fertility-enhancing cocooning effect. Hence, fertility rates in 2022 were lower compared to the preceding year.

In contrast, stricter containment policies in lower-trust countries were associated with a decline in fertility and subsequent recovery, in line with the findings of Plach et al. (2023). Less stringent policies would thus be linked to a recovery in fertility. However, our analysis revealed that the pattern of fertility decline and recovery disappeared for conceptions in 2021. One possible interpretation could be that individuals developed copying strategies with regard to containment policies (Toffolutti et al., 2022), and thus, the link between the stringency index and fertility vanished. Apart from government policies, actual trends in mobility, work and socialization outside of the home may also affect birth trends. However, a separate analysis found no evidence for an association between pandemic-related behavioural trends, as reflected by the normalcy index, and the fertility rate. The analysis shows a negative association between the initial vaccination rollout and fertility, whereas the completion of the full first vaccination course (usually consisting of two doses) is linked to fertility recovery. Similar to Bujard and Andersson (2024), we interpret this finding as a behavioural response to the perceived potential risks of vaccination for pregnant women. The decrease in fertility during the introduction of vaccination was probably due to an ‘anticipatory’ postponement of births, as some women decided to complete their vaccination course before becoming pregnant to minimize the risks of potential COVID-19 infection to their health and pregnancy outcomes. Vaccination is strongly recommended for women with childbearing intentions because of the increased risk of severe illness during pregnancy and elevated risk of complications during pregnancy due to COVID-19 infection (Wei et al., 2021). Furthermore, women may be concerned about the hypothetical negative impact of vaccination on their health, fecundity or pregnancy outcomes. Such concerns have not been substantiated by scientific evidence (e.g., Chen et al., 2021) but have had a strong echo in social media. Successful vaccination campaigns are also closely linked to the “return to normal life” discussed above (Neyer et al., 2022): the launch of broad-based vaccination programs signalled that the pandemic restrictions are likely to be lifted soon and that the pandemic was getting under control. Decisions to postpone pregnancy until the full course of the vaccination had been completed contributed to the downturn in birth rates in January-April 2022 (and, in some countries, as early as December 2021) and later to their slight recovery in mid-2022.

In line with the scientific evidence, our analysis does not suggest a direct impact of vaccination on fertility trends. Specifically, conceptions in most countries started dropping already in the early stage of the vaccination process, when most women of reproductive age had not yet been eligible for receiving the first dose. If vaccines as such would directly contribute to the observed drop in births, birth rates in most countries would have started falling later than observed, and the decline would be of a similar magnitude across all countries during the peak of their vaccination campaigns.

Our research reveals large cross-country diversity in fertility responses to the COVID-19 pandemic (Plach et al., 2023; Sobotka et al., 2023). The downturn in birth rates since approximately January 2022 is visible in most, but not all, of the analysed countries: it did not occur or was only slight in most of the non-European countries studied. In Israel, birth rates decreased earlier, even in October and November 2021, which is consistent with its earlier and more intensive vaccination rollout, with women of reproductive age eligible for vaccination since January 2021 (Mathieu et al., 2020). Portugal is an outlier where birth rates have moved in the opposite direction and started to recover since early 2022, following their steep decline during the main phases of the COVID-19 pandemic. The diversity of birth trends between countries in the wake of the vaccination campaign, also documented by Jasilioniene et al. (2024) points out again that COVID-19 vaccination did not have any direct biological impact on birth trends.

Many countries with a mild “baby bump” during the periods of the most severe infection waves and government containment policies experienced an especially sharp downturn in births since early 2022. This includes the Nordic countries, Germany and Switzerland, Czechia, Greece and Hungary. To some extent, this suggests that a “return to normality” and the end of the “cocooning effect” – people socializing more outside of the home, attending cinemas, restaurants, sports venues and recreation facilities, travelling again for leisure and recreation, and employees returning to the offices – is a key part of the explanation of declining birth rates. In many countries, the birth trajectory has returned to pre-pandemic levels.

Our research demonstrates the usefulness of looking at birth trends and their drivers from a short-term (monthly or even more detailed) perspective rather than taking the usual approach of analysing annual data. Annual data do not have sufficient granularity to study the dynamics and drivers of fertility during sudden shocks such as the COVID-19 pandemic.

The COVID-19 pandemic has cast a long shadow on birth trends. This also includes factors we have not been able to analyse, including the disruptions to dating, intimate life, and partnership formation (e.g.,Ting & McLachlan, 2022), which may affect birth trends for an extended period of time. The changes in birth rates until autumn 2022 analysed here do not mark the end of the period of unstable, “rollercoaster” fertility in low-fertility countries. As the COVID-19 pandemic gradually weakened after the vaccination campaign, new crises and disruptions emerged, especially the Russian war against Ukraine since February 2022 and the resulting surge in inflation and economic uncertainty. This pushed birth rates in many of the analysed countries to a new downturn occurring since late 2022 and continuing through 2023 (HFD, 2023), with many countries recording new lows in fertility rates.

## Data Availability

All data produced are available online at different online databases: Human Fertility Database (https://www.humanfertility.org), OECD website (https://oecd.org), Our World in Data (https://ourworldindata.org) and the Oxford Covid-19 Government Response Tracker (OxCGRT) at GitHub (https://github.com/OxCGRT/covid-policy-dataset)

## Acknowledgement

Many thanks to Zuzanna Brzozowska, Ester González-Prieto, and Miguel Sánchez-Romero for helpful comments and suggestions.

## Footnotes

1 See, e.g., the “Updated advice on COVID-19 vaccination in pregnancy and women who are breastfeeding” by the Royal College of Midwifes issued on 30 December 2020 (https://www.rcm.org.uk/news-views/news/2020/december/updated-advice-on-covid-19-vaccination-in-pregnancy-and-women-who-are-breastfeeding/), and the UK government statement on 7 January 2021 based on Public Health England recommendation (https://www.gov.uk/drug-safety-update/covid-19-vaccines-pfizer-slash-biontech-and-covid-19-vaccine-astrazeneca-current-advice).

2 For a comparison see e.g., Fiolet et al. (2022).

3 Specifically, the monthly number of births are adjusted for weekday variations of births.

4 Using different clustering methods, such as k-Means or hierarchical clustering, does not yield different country clustering for the set of countries used in this study.

5 The clustering gives similar results to the ones based on pre-pandemic social support used by Plach et al. (2023). Canada and Ireland display lower social support but higher trust. Conversely, Austria, France, and Belgium show high social support but lower trust.

6 The Breusch‒Pagan test rejected the null of homoskedasticity. We performed the Woolridge test for serial correlation in panel-data models and the Pesaran’s CD test for testing for cross-sectional dependence. Both tests rejected the absence of autocorrelation and cross-section dependence, respectively.

7 The normalcy index is composed from eight indicators covering three different domains: transport and travel; recreation and entertainment; and retail and work, where each indicator is measured as a percentage of its pre-pandemic level (as the average values of each indicator in January and February 2020). The correlation between the normalcy index and the stringency index is high (rho=-0.88, p<0.001). Therefore, we do not include both indices in our analysis simultaneously.

8 In an additional analysis, we also included period interactions with the economic indicators. However, there were no visible differences in the estimated coefficients for the latter covariates between the two periods.

## Notes

### Competing Interest Statement

The authors have declared no competing interest.

### Funding Statement

This study did not receive any funding.

### Author Declarations

The study used (or will use) ONLY openly available human data that were originally located at the Human Fertility Database (https://www.humanfertility.org), OECD website (https://oecd.org), Our World in Data (https://ourworldindata.org) and the Oxford Covid-19 Government Response Tracker (OxCGRT) at GitHub (https://github.com/OxCGRT/covid-policy-dataset)

