## Supplemental Tables S1 and S2, and Supplemental Figure S1 for "Fertility decline in the later phase of the COVID-19 pandemic: The role of policy interventions, vaccination programmes, and economic uncertainty"

Table S1: Estimated model coefficients of the fixed effect models of the monthly seasonally adjusted TFR per 100 women.

| | (1)<br>Model 1 | (2)<br>Model 2 | (3)<br>Model 3 |
|---|---|---|---|
| **Economic indicators** | | | |
| Unemployment rate | -0.0510 | -0.0939 | -0.0235 |
| | (0.121) | (0.136) | (0.151) |
| Consumer price index | $-0.600^{***}$ | $-0.738^{***}$ | $-0.775^{***}$ |
| | (0.139) | (0.167) | (0.154) |
| **Non-pharmaceutical policy interventions** | | | |
| *All countries* | | | |
| Stringency | $0.0389^{*}$ | | |
| | (0.0168) | | |
| Economic Support | 0.0106 | | |
| | (0.00900) | | |
| *Lower-trust countries* | | | |
| Stringency | | $-0.107^{*}$ | |
| | | (0.0449) | |
| Lagged stringency | | $0.105^{*}$ | |
| | | (0.0448) | |
| Economic support | | 0.00590 | |
| | | (0.00922) | |
| *Higher-trust countries* | | | |
| Stringency | | $0.140^{*}$ | |
| | | (0.0566) | |
| Lagged stringency | | -0.00783 | |
| | | (0.0561) | |
| Economic support | | 0.0297 | |
| | | (0.0257) | |
| **Conceptions in Feb. 2020–Dec. 2020** | | | |
| *Lower-trust countries* | | | |
| Stringency | | | $-0.131^{**}$ |
| | | | (0.0398) |
| Lagged stringency | | | $0.108^{**}$ |
| | | | (0.0340) |
| Economic support | | | 0.0191 |
| | | | (0.0160) |
| Higher-trust countries | | | |
| Stringency | | | $0.112^{*}$ |

| | | | |
|---|---|---|---|
| | | | (0.0450) |
| Lagged stringency | | | 0.0787* |
| | | | (0.0360) |
| Economic support | | | 0.000910 |
| | | | (0.0237) |
| **Conceptions in Jan. 2021–Jan. 2022** | | | |
| *Lower-trust countries* | | | |
| Stringency | | | 0.0644 |
| | | | (0.0554) |
| Lagged stringency | | | -0.0430 |
| | | | (0.0583) |
| Economic support | | | 0.00499 |
| | | | (0.0121) |
| *Higher-trust countries* | | | |
| Stringency | | | 0.187· |
| | | | (0.0938) |
| Lagged stringency | | | -0.0674 |
| | | | (0.0963) |
| Economic support | | | 0.00801 |
| | | | (0.0240) |
| **Cum. per. vaccinated** | | | |
| First dose | -0.284*** | -0.292*** | -0.245*** |
| | (0.0308) | (0.0245) | (0.0198) |
| Primary course | 0.235*** | 0.254*** | 0.213*** |
| | (0.0333) | (0.0261) | (0.0227) |
| **Pandemic indicators** | | | |
| Excess mortality | -0.0887*** | -0.0577* | -0.0616· |
| | (0.0233) | (0.0262) | (0.0334) |
| First Covid-19 wave | -7.846*** | -5.852*** | -4.838** |
| | (0.653) | (1.376) | (1.305) |
| Constant | 215.5*** | 228.9*** | 231.9*** |
| | (14.31) | (17.63) | (16.64) |
| Observations | 624 | 624 | 624 |
| Countries | 26 | 26 | 26 |
| Within R-squared | 0.424 | 0.477 | 0.515 |

Standard errors in parentheses
· $p < 0.10$, * $p < 0.05$, ** $p < 0.01$, *** $p < 0.001$

Table S2: Estimated model coefficients of alternative fixed effect model of the monthly seasonally adjusted TFR per 100 women with normalcy index (Model 1A) and vaccination indicators of women of childbearing age (Model 1B).

| | (1)<br>Model 1A | (2)<br>Model 1B |
|---|---|---|
| **Economic indicators** | | |
| Unemployment rate | -0.388$^{\cdot}$ | 0.222 |
| | (0.214) | (0.233) |
| Consumer price index | -0.620$^{***}$ | -1.075$^{***}$ |
| | (0.159) | (0.221) |
| **Non-pharmaceutical policy interventions** | | |
| Stringency | | 0.0213 |
| | | (0.0222) |
| Economic Support | 0.0168 | 0.0266$^{*}$ |
| | (0.0132) | (0.0123) |
| Normalcy | -0.0270 | |
| | (0.0213) | |
| **Cum. Perc. vaccinated** | | |
| *Total population* | | |
| First dose | -0.260$^{***}$ | |
| | (0.0307) | |
| Primary course | 0.213$^{***}$ | |
| | (0.0330) | |
| *Population of childbearing age* | | |
| First dose | | -0.228$^{***}$ |
| | | (0.0236) |
| Primary course | | 0.210$^{***}$ |
| | | (0.0368) |
| **Pandemic indicators** | | |
| Excess mortality | -0.0891$^{***}$ | -0.0789$^{**}$ |
| | (0.0183) | (0.0210) |
| First Covid-19 wave | -8.275$^{***}$ | -7.793$^{***}$ |
| | (0.696) | (0.758) |
| Constant | 224.0$^{***}$ | 264.0$^{***}$ |
| | (15.60) | (22.00) |
| Observations | 528 | 497 |
| Countries | 22 | 22 |
| Within R-squared | 0.393 | 0.445 |

Standard errors in parentheses
$^{\cdot}$ $p < 0.10$, $^{*}$ $p < 0.05$, $^{**}$ $p < 0.01$, $^{***}$ $p < 0.001$

Figure S1: Time trends of monthly total fertility rate from Oct. 2019 to Oct. 2022 (left axis), and time trends of harmonized unemployment rate, consumer price index, stringency index, economic support index, cumulative share of the population that has received at least one dose of the vaccination and that has completed the primary vaccination course, and excess mortality from Jan. 2019 to Jan. 2022 (right axis), by country.

#### Austria

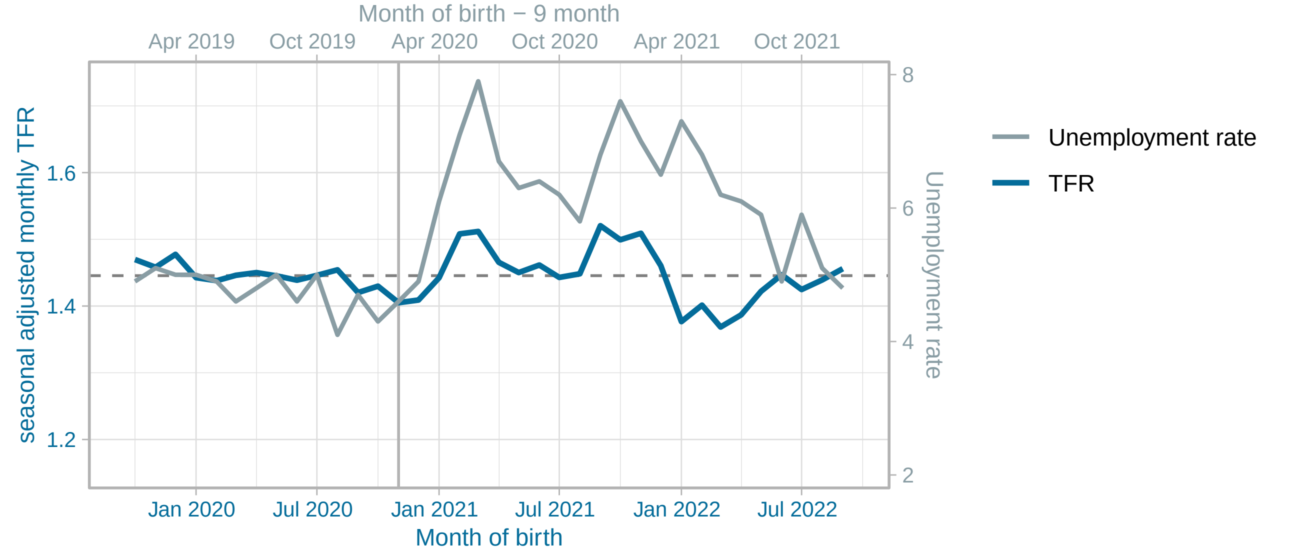


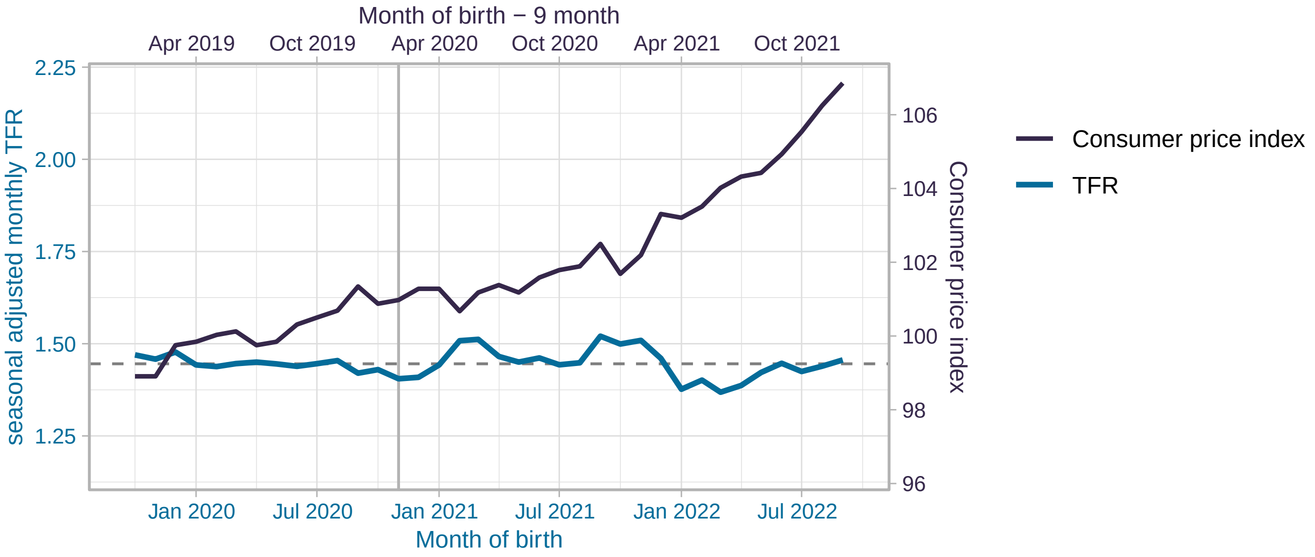


(figure continued on next page)

#### Austria

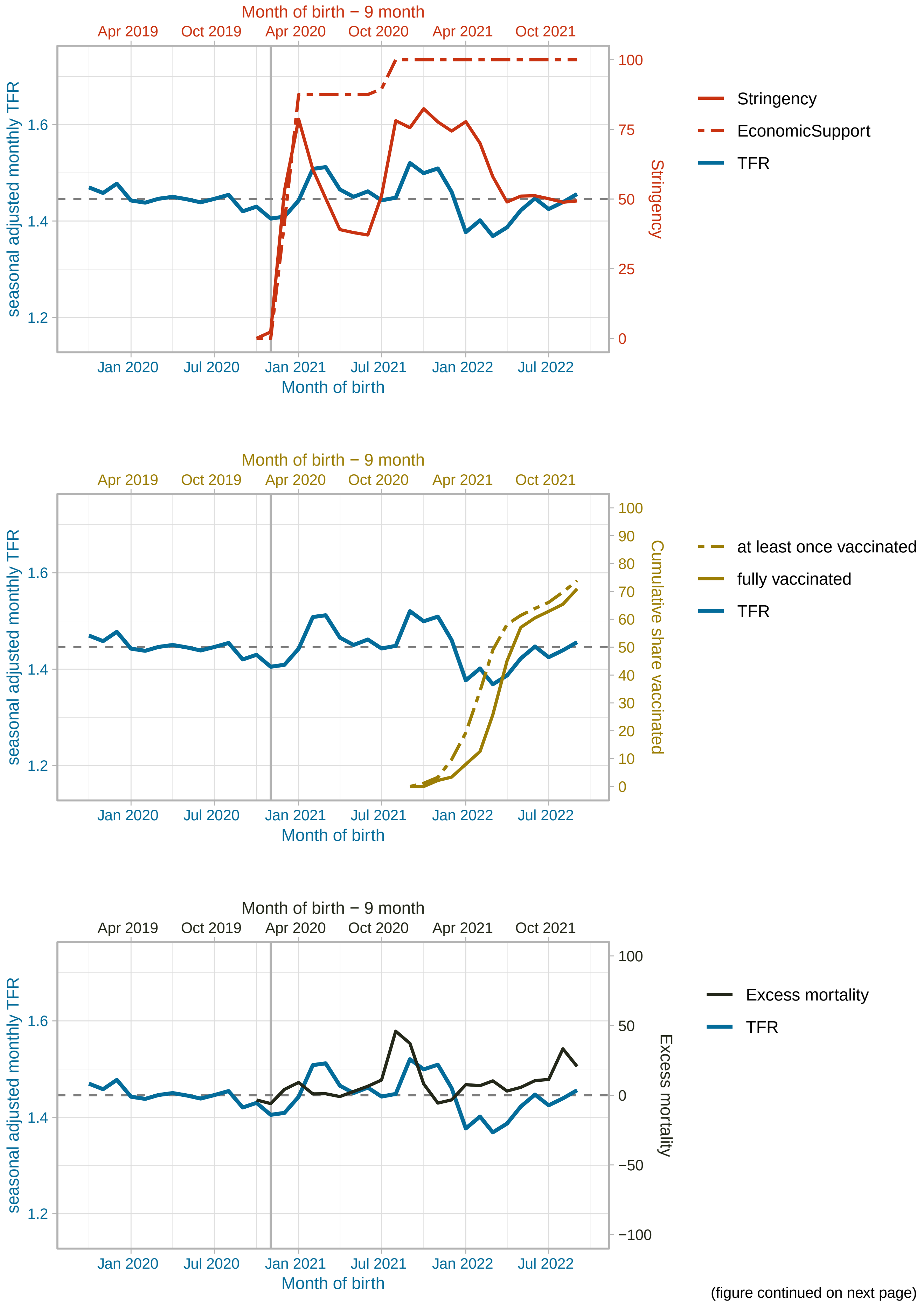


(figure continued on next page)

#### Belgium

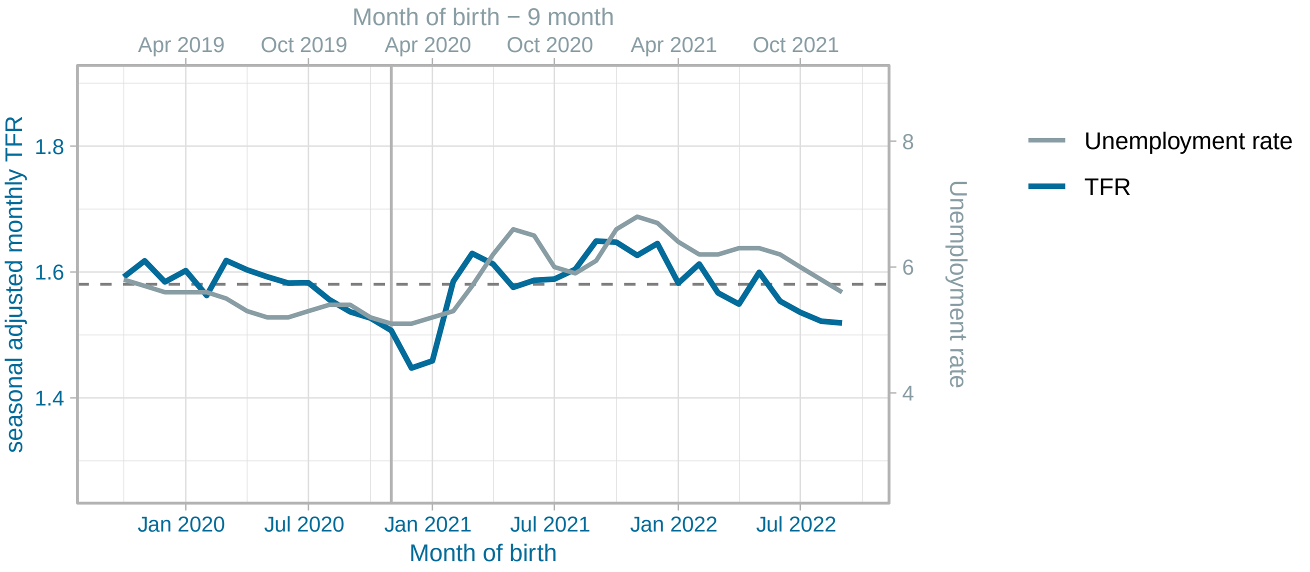


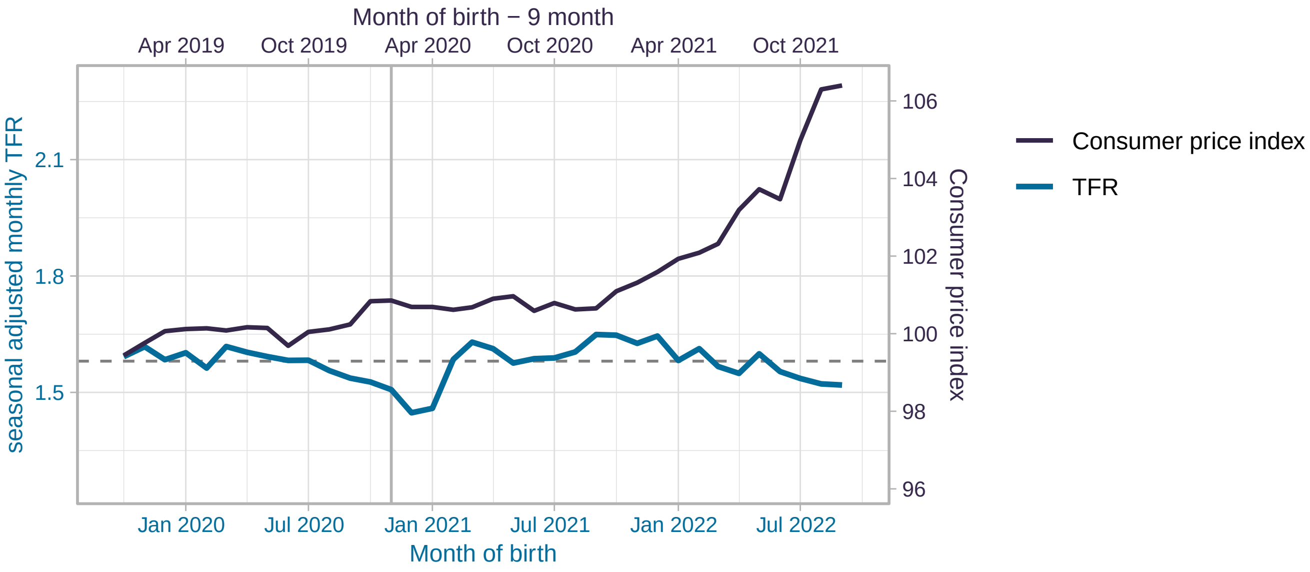


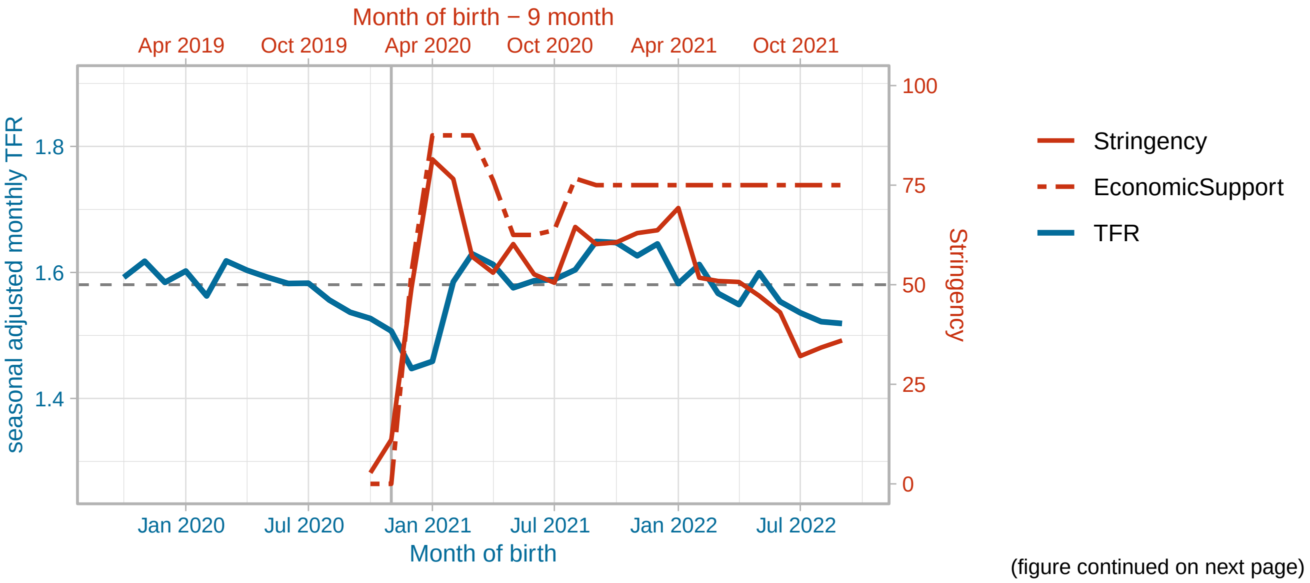


(figure continued on next page)

Belgium

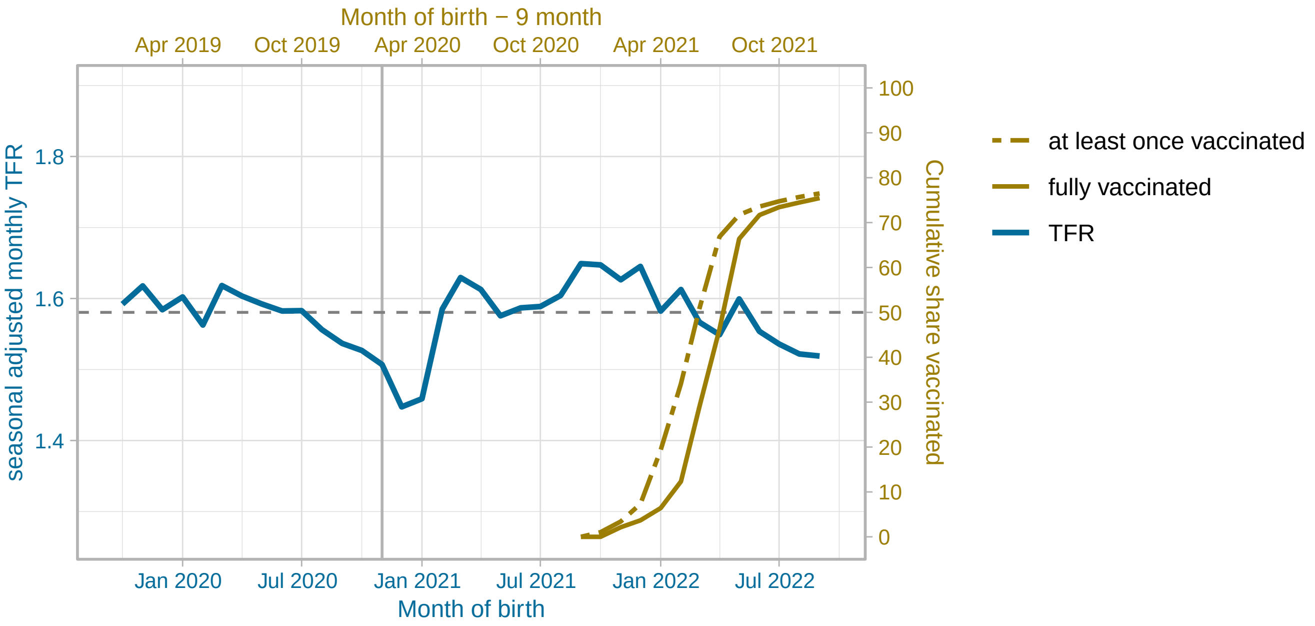


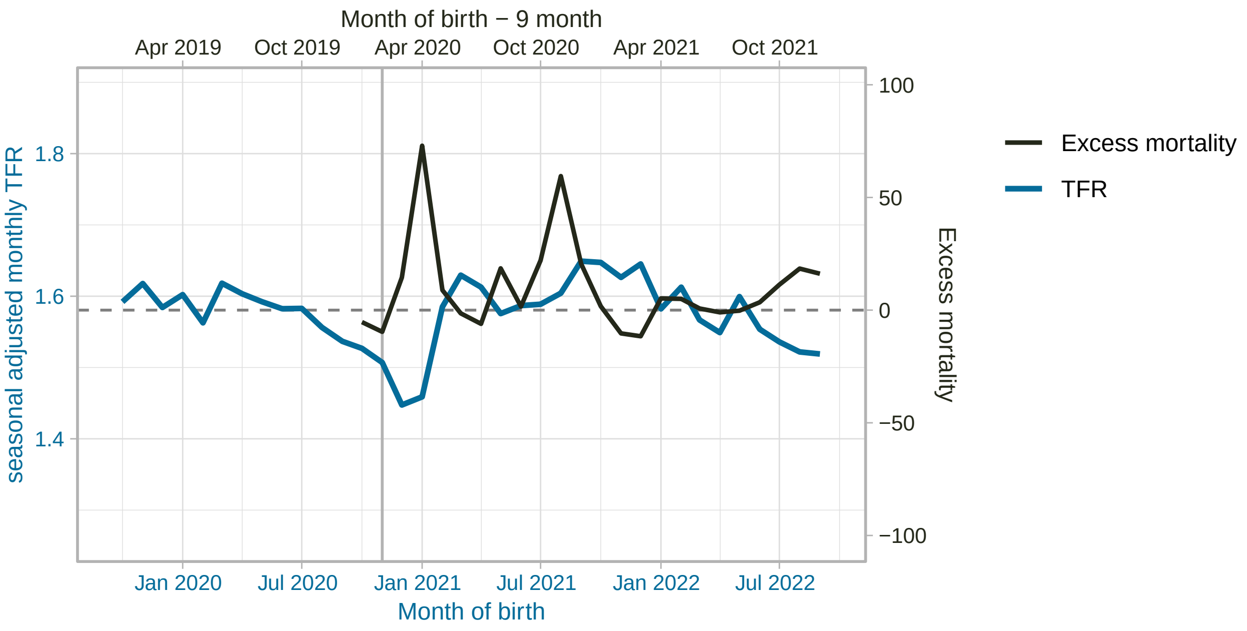


(figure continued on next page)

Canada

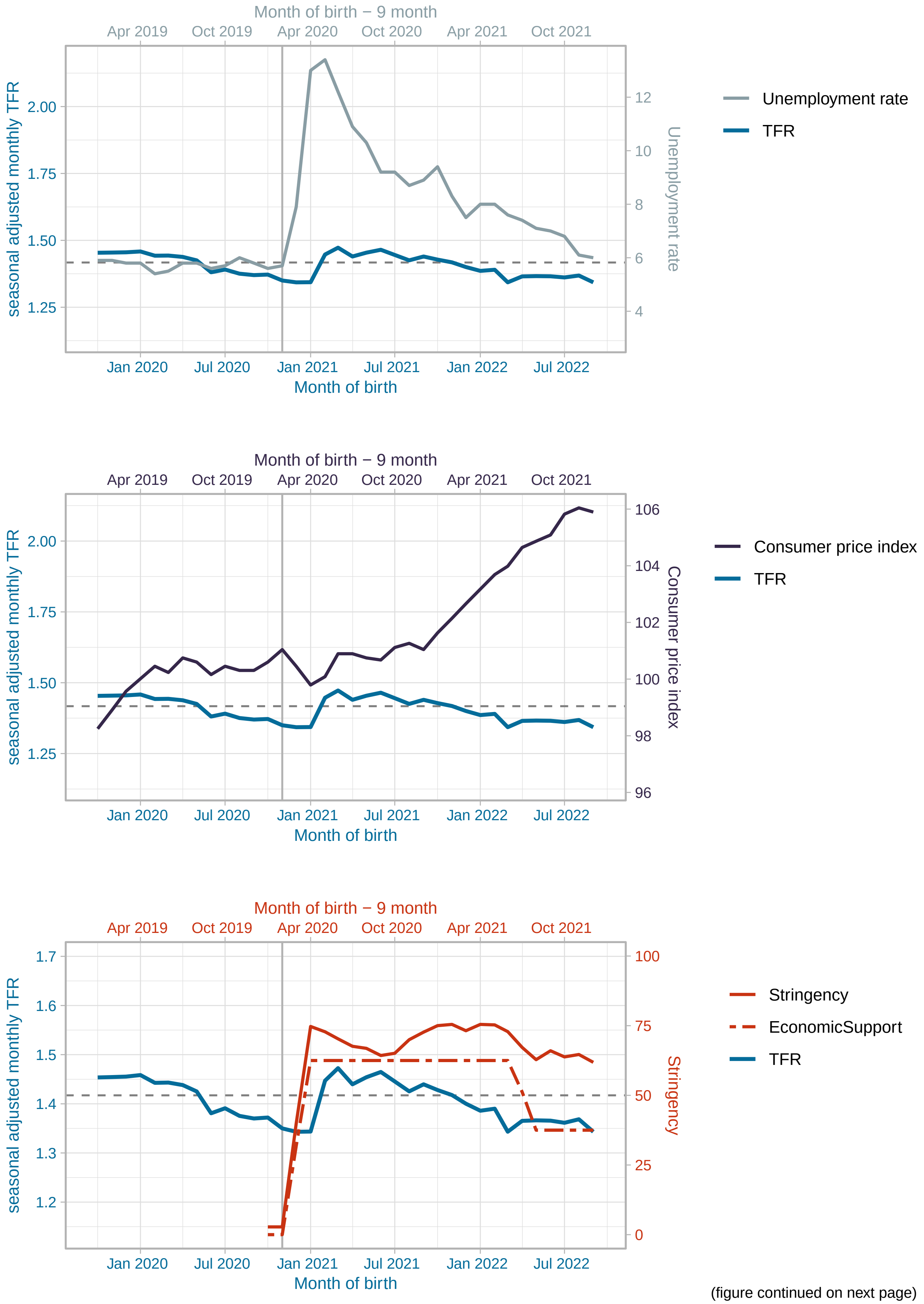


(figure continued on next page)

Canada

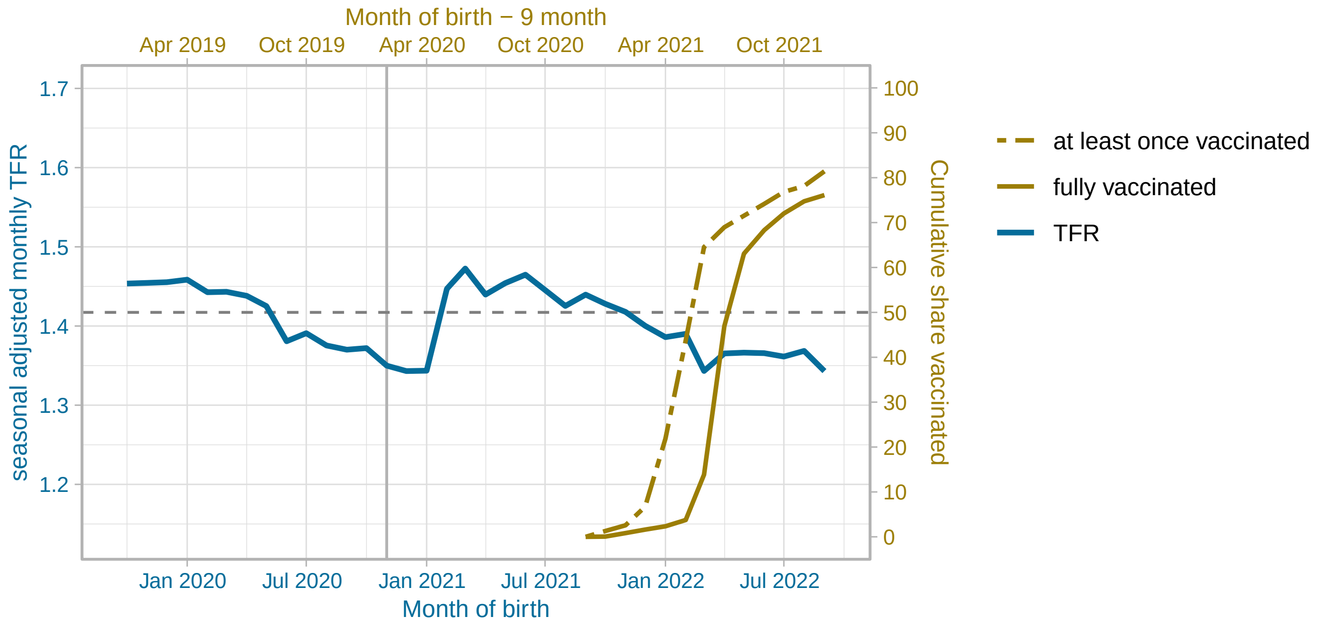


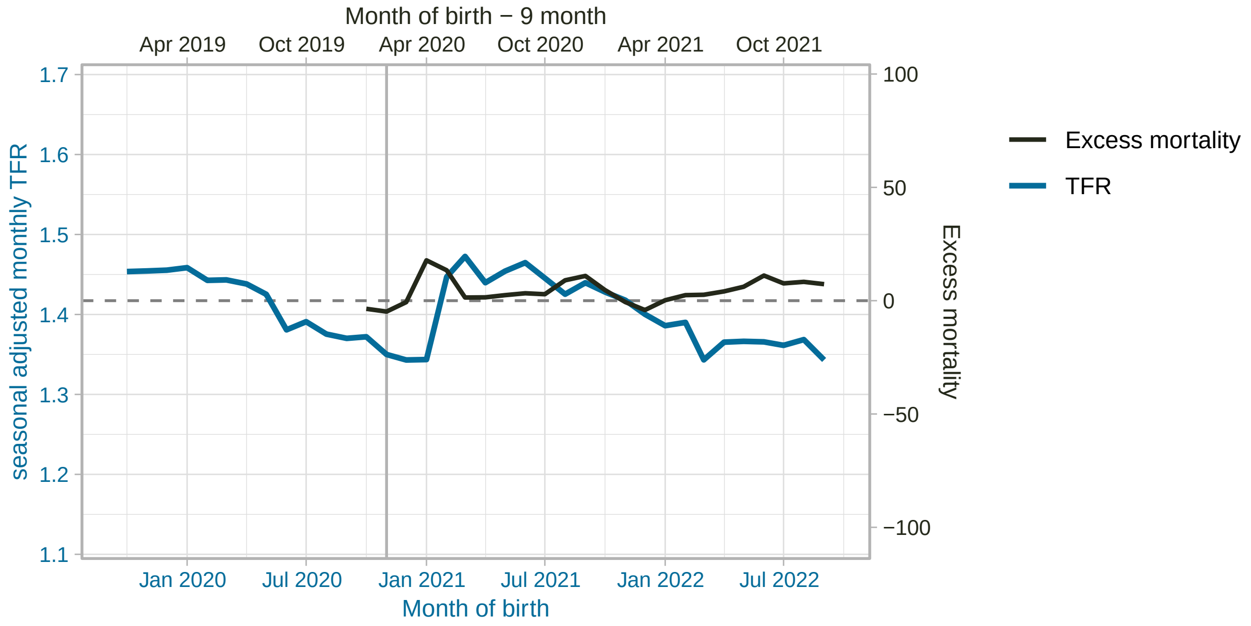


(figure continued on next page)

#### Switzerland

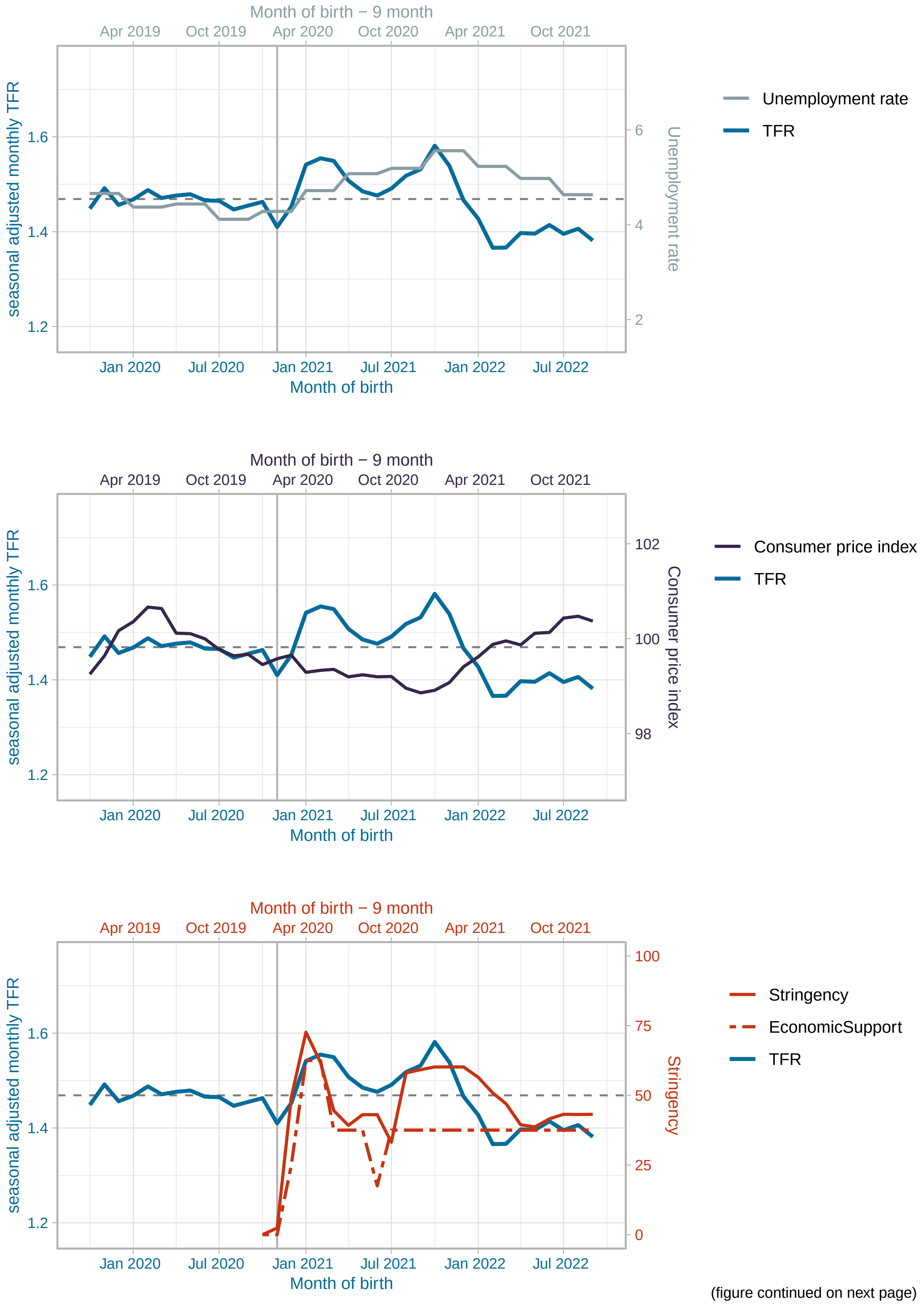


(figure continued on next page)

#### Switzerland

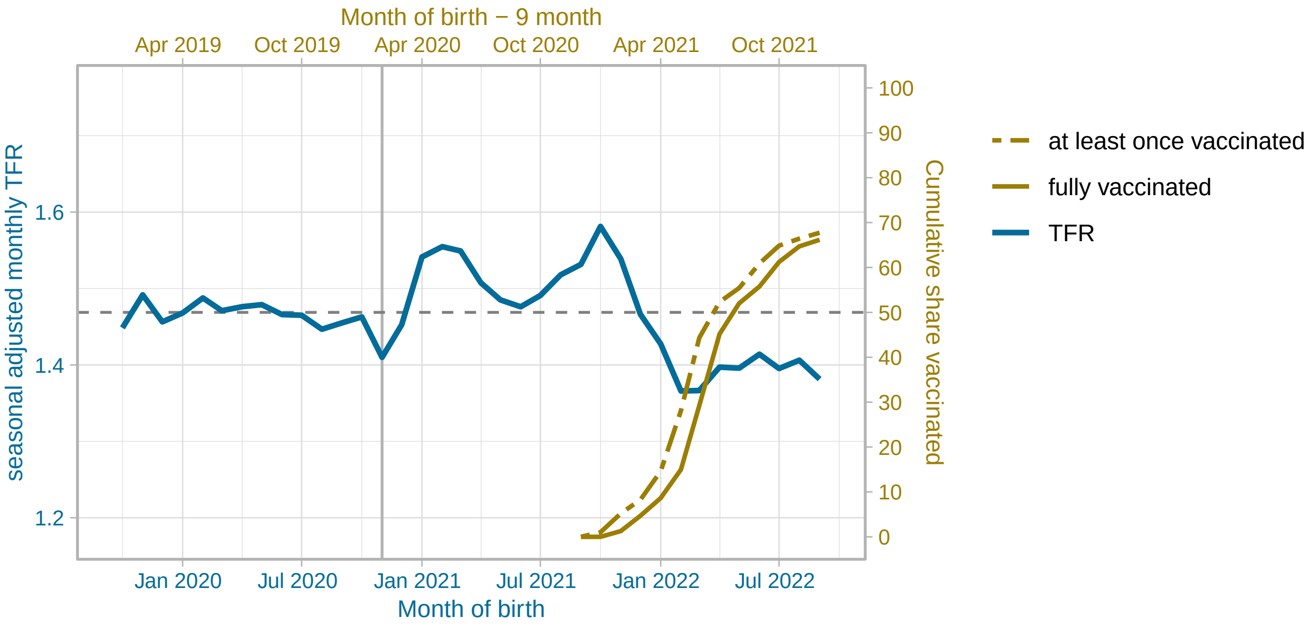


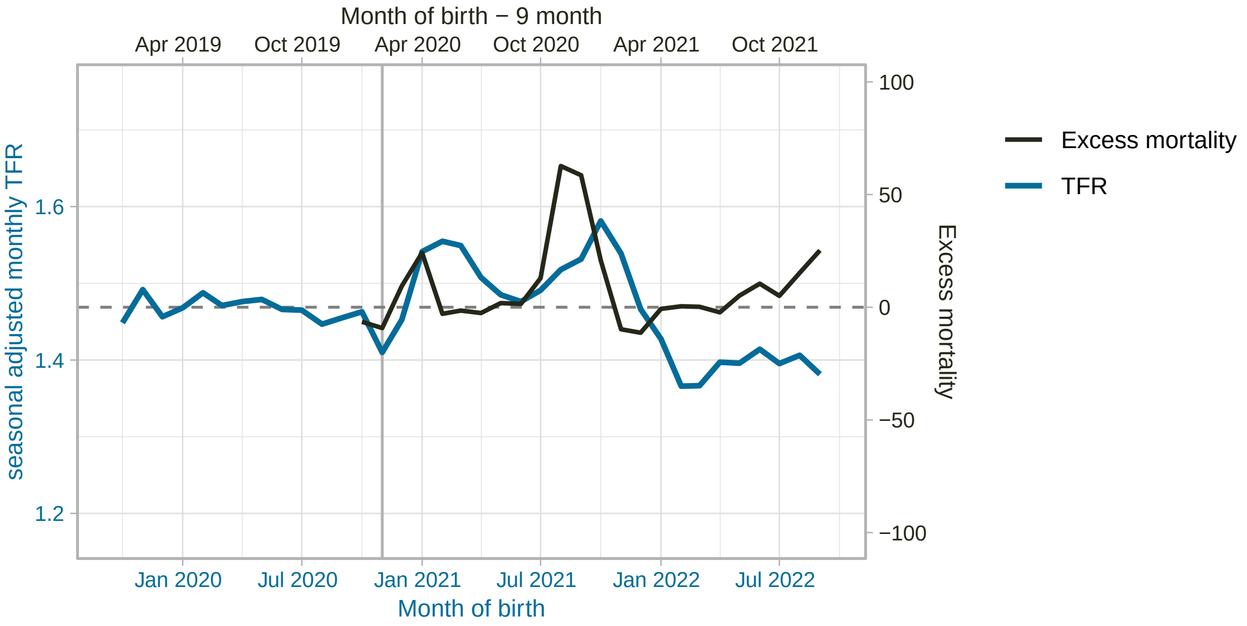


(figure continued on next page)

#### Czechia

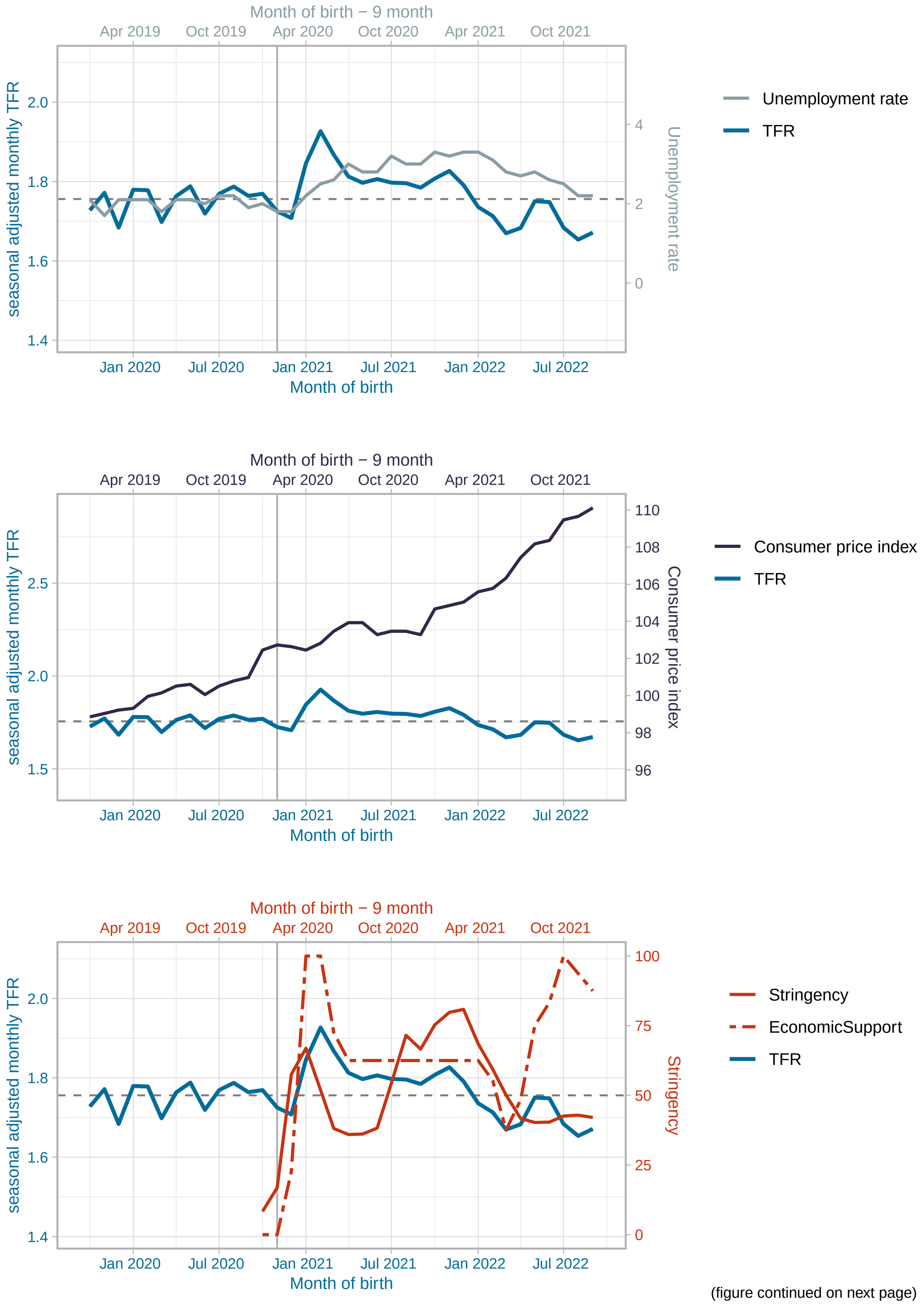


(figure continued on next page)

Czechia

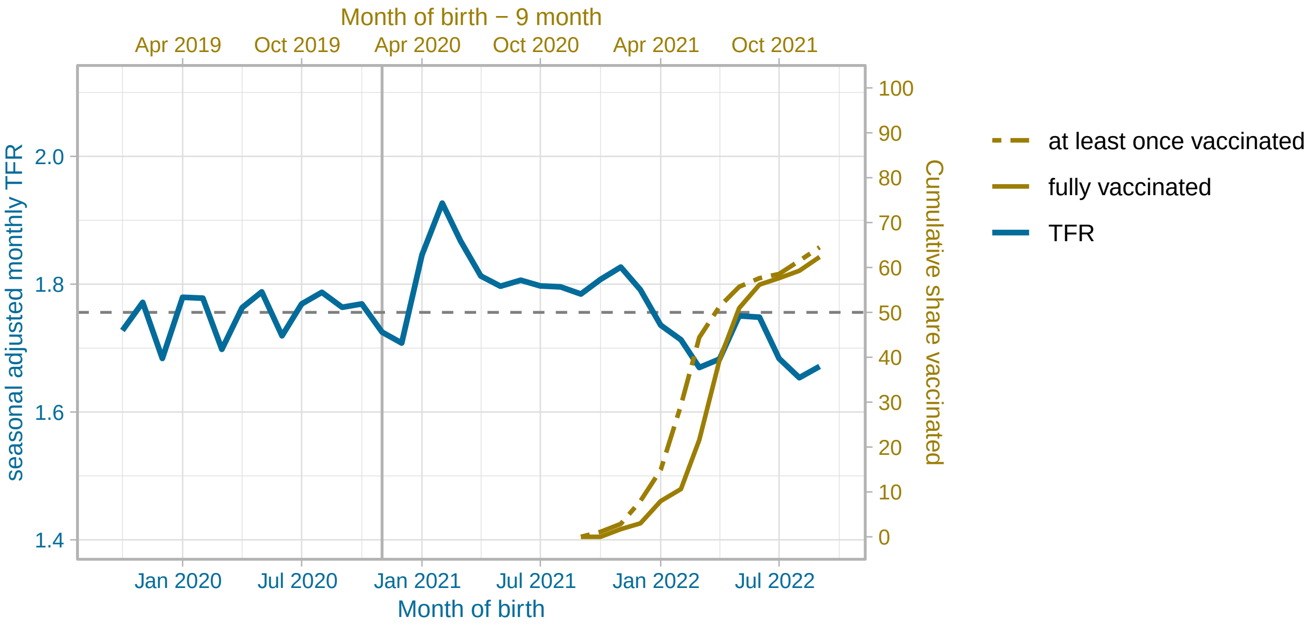


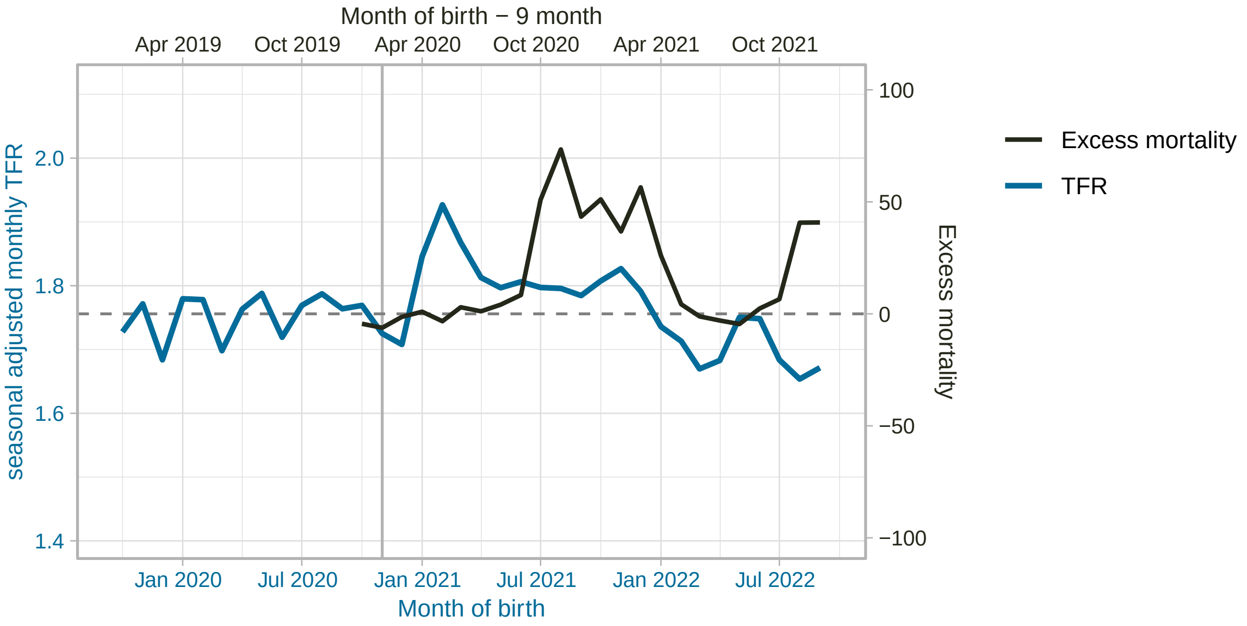


(figure continued on next page)

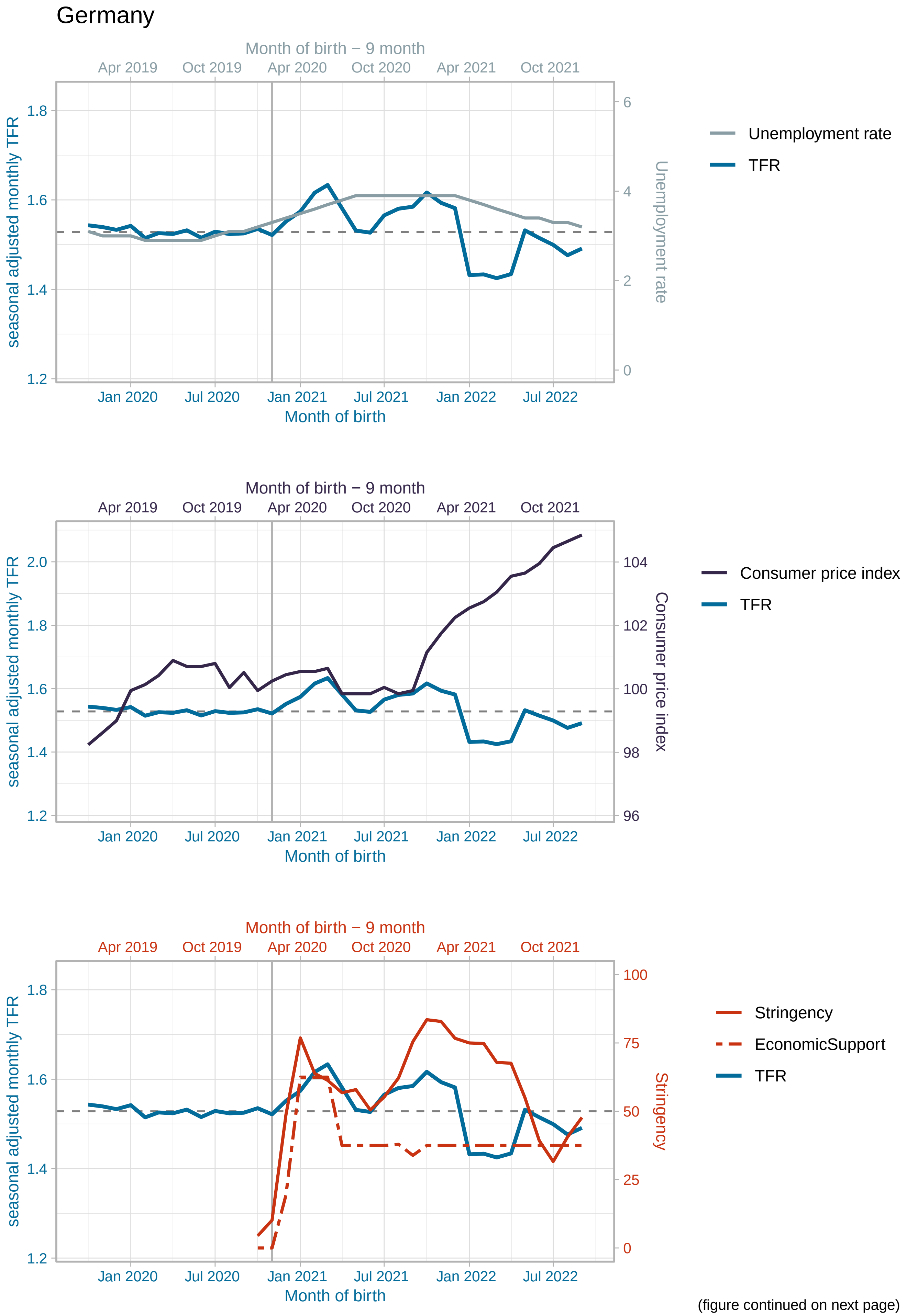


(figure continued on next page)

#### Germany

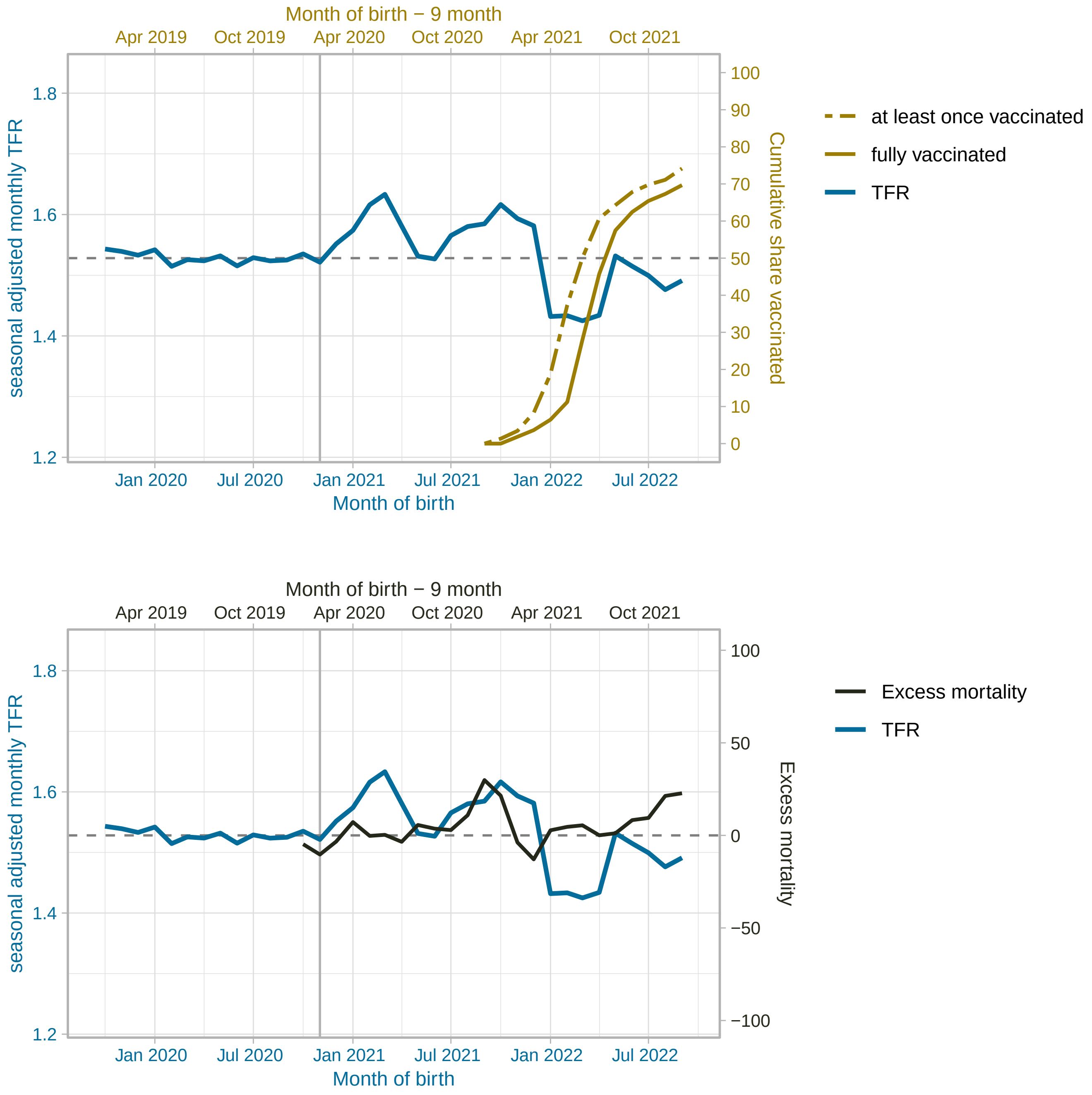


(figure continued on next page)

Denmark

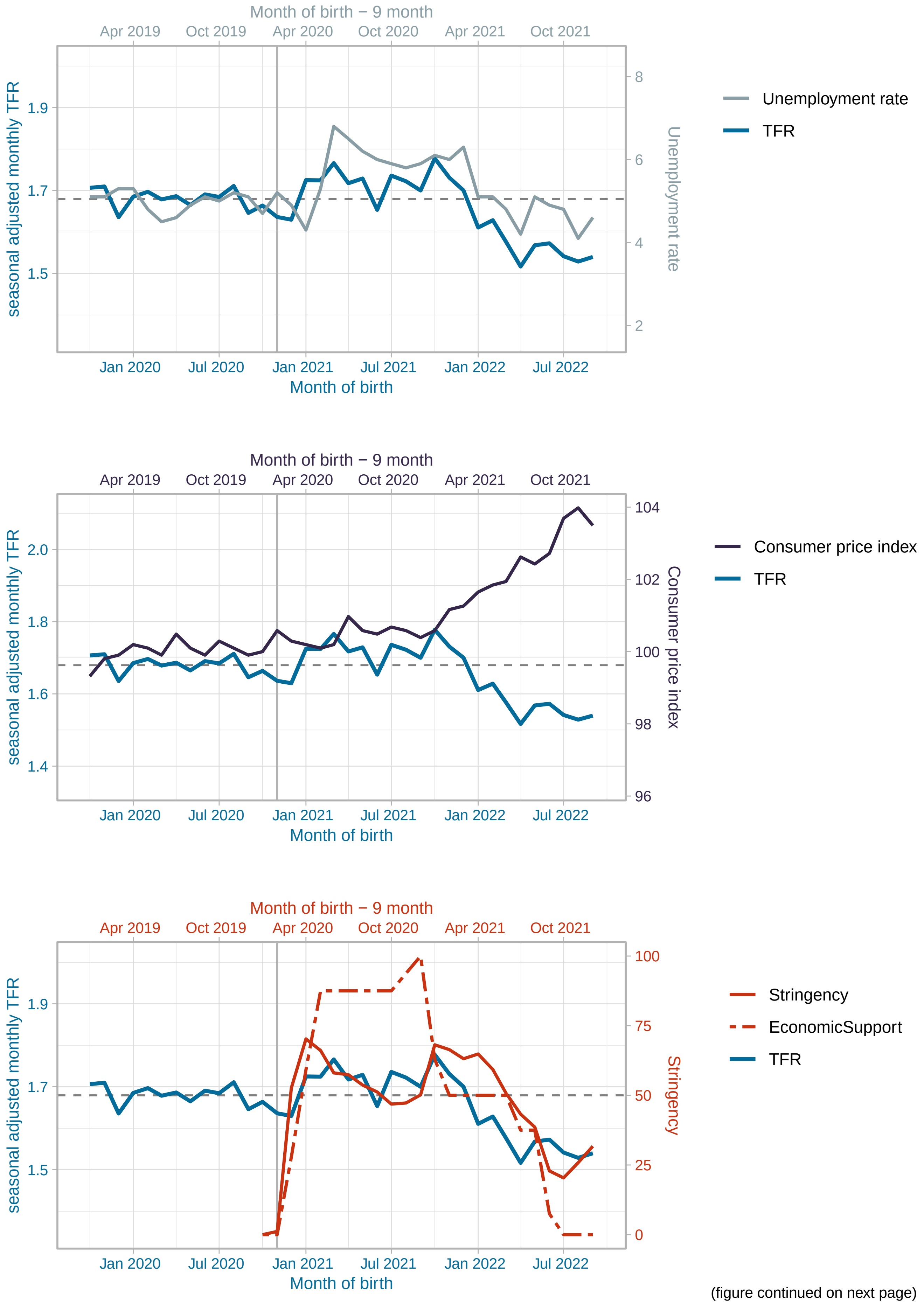


(figure continued on next page)

#### Denmark

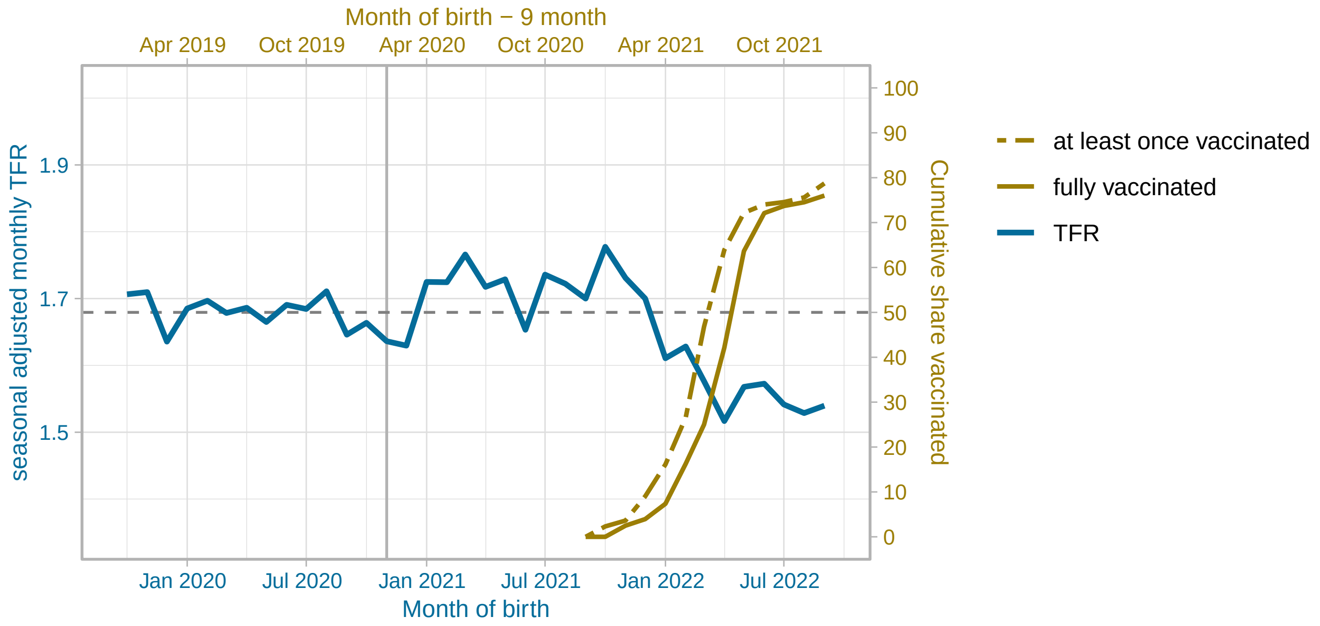


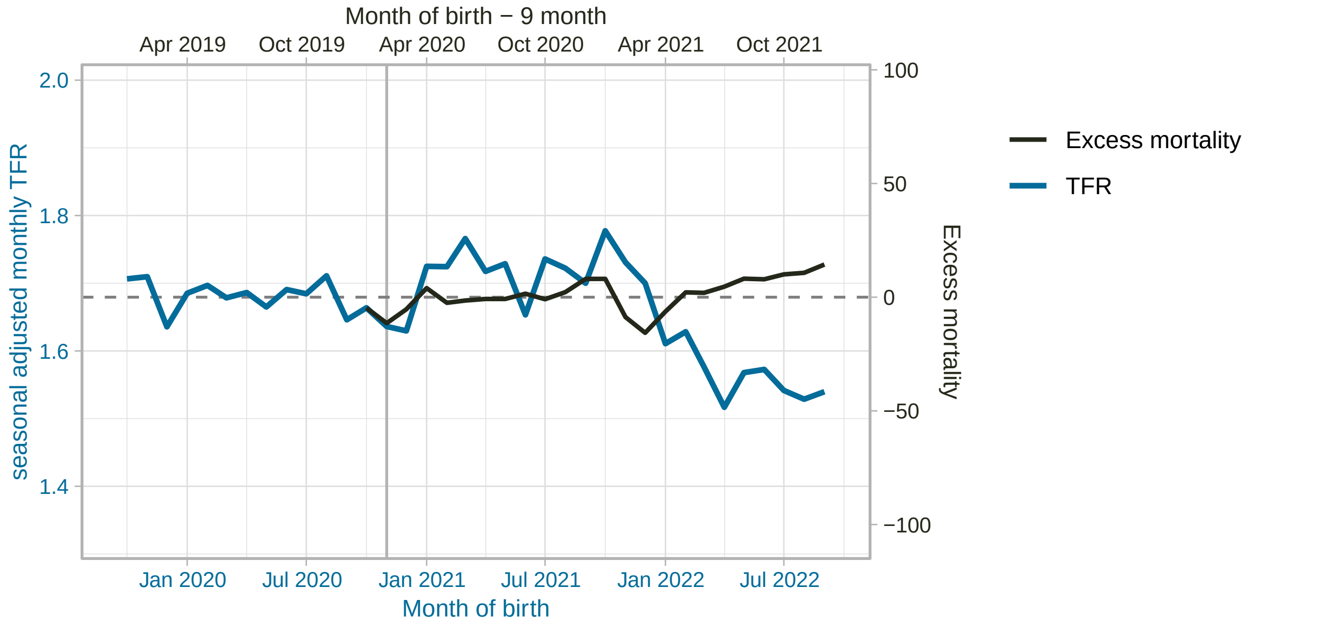


(figure continued on next page)

Spain

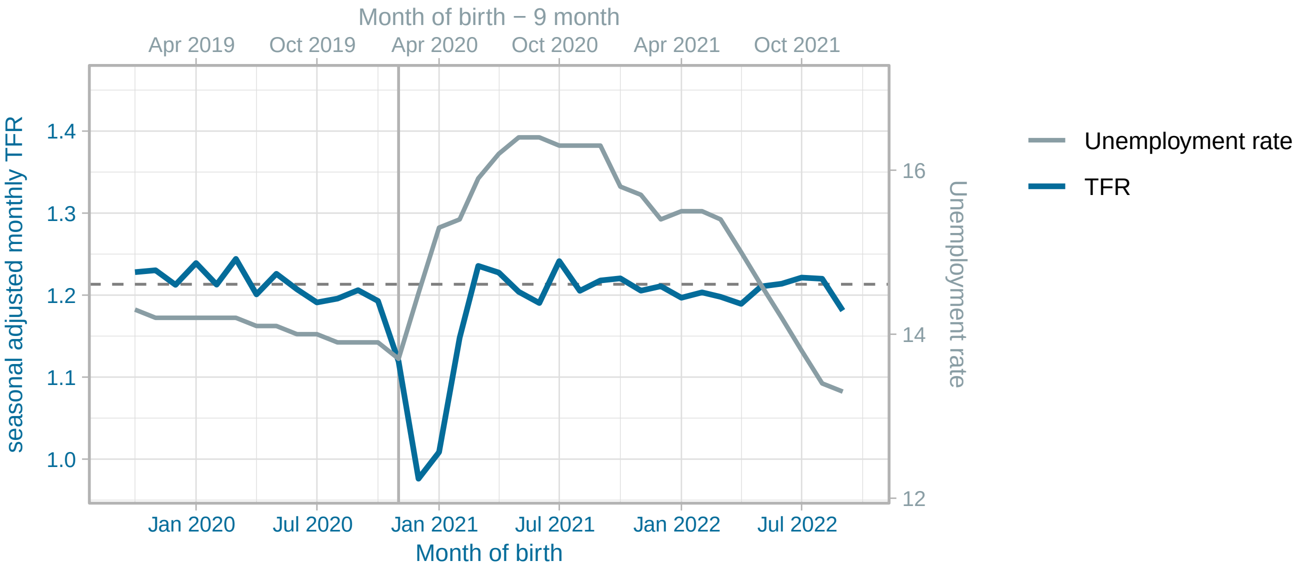


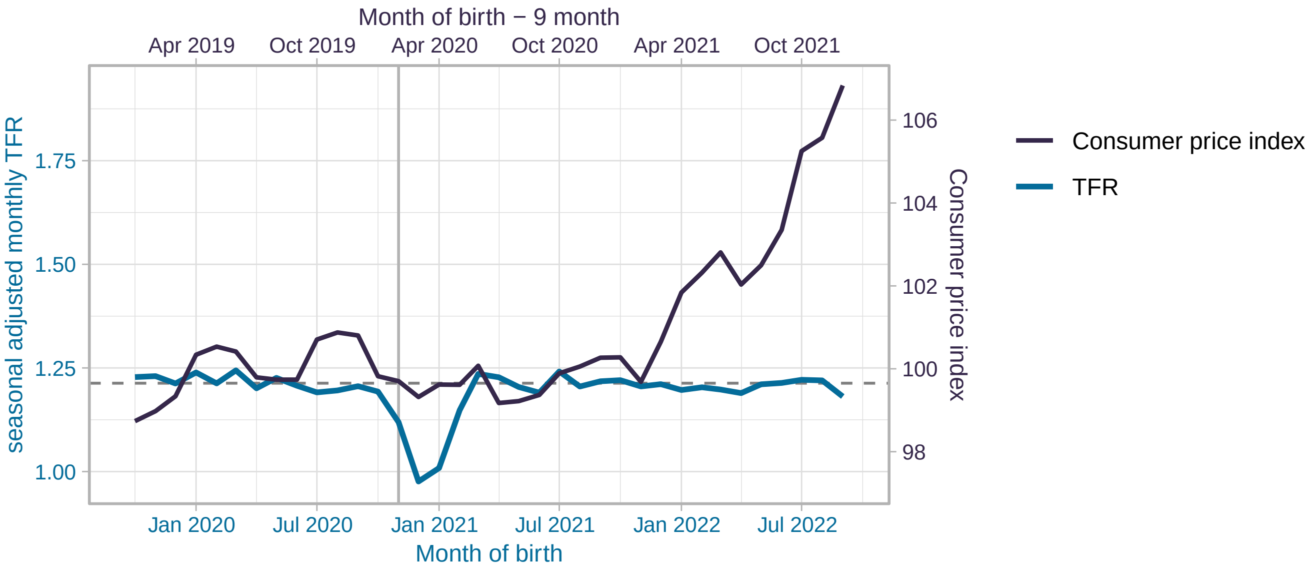


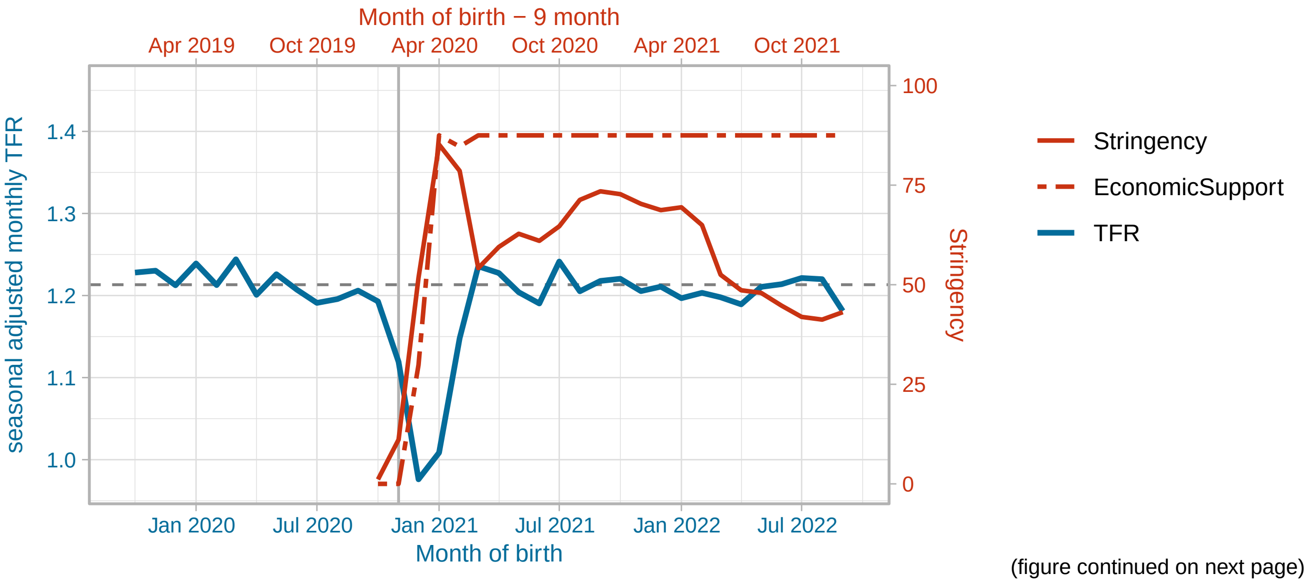


(figure continued on next page)

Spain

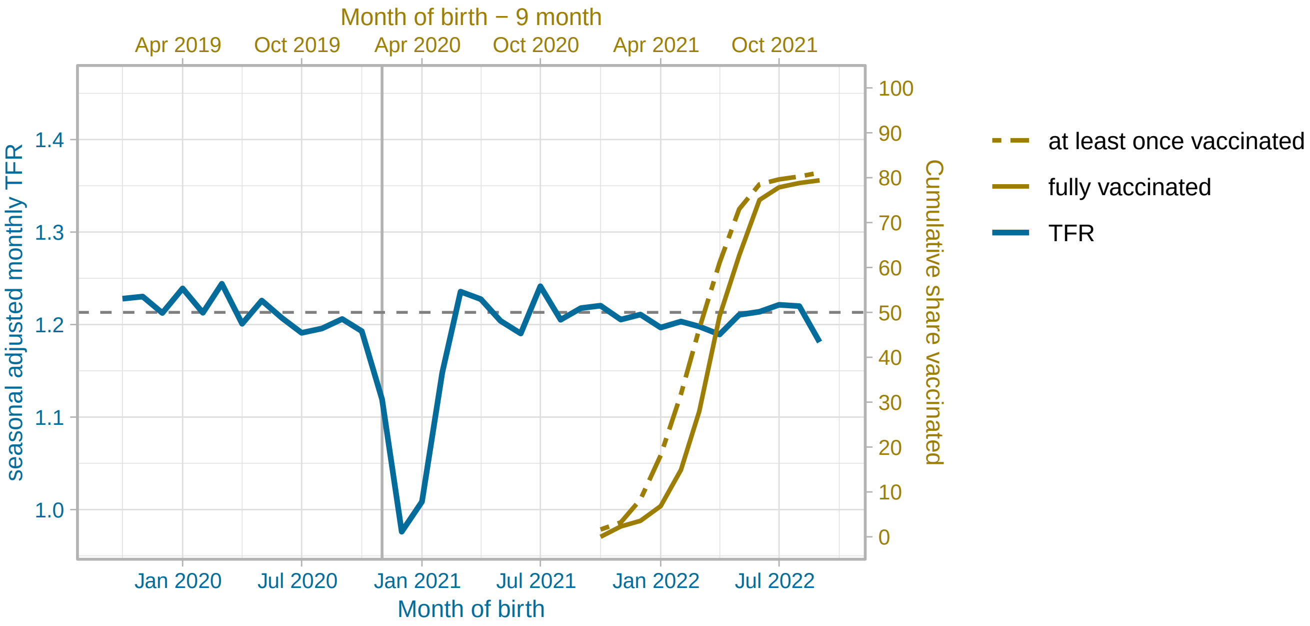


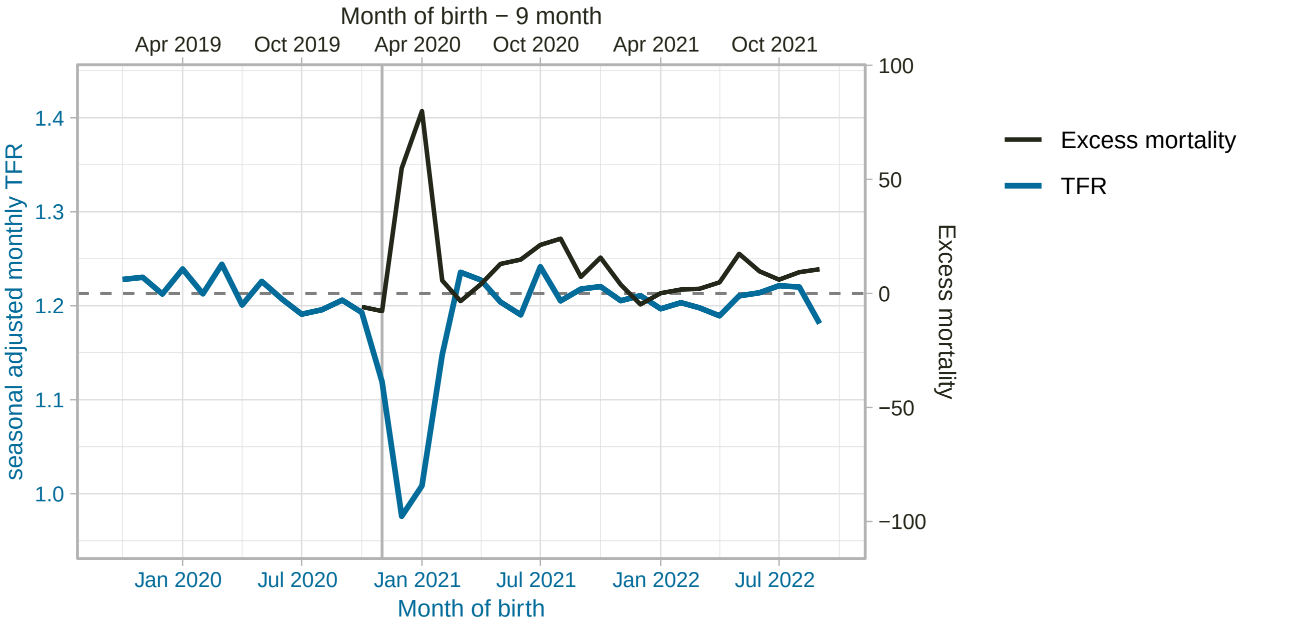


(figure continued on next page)

#### Finland

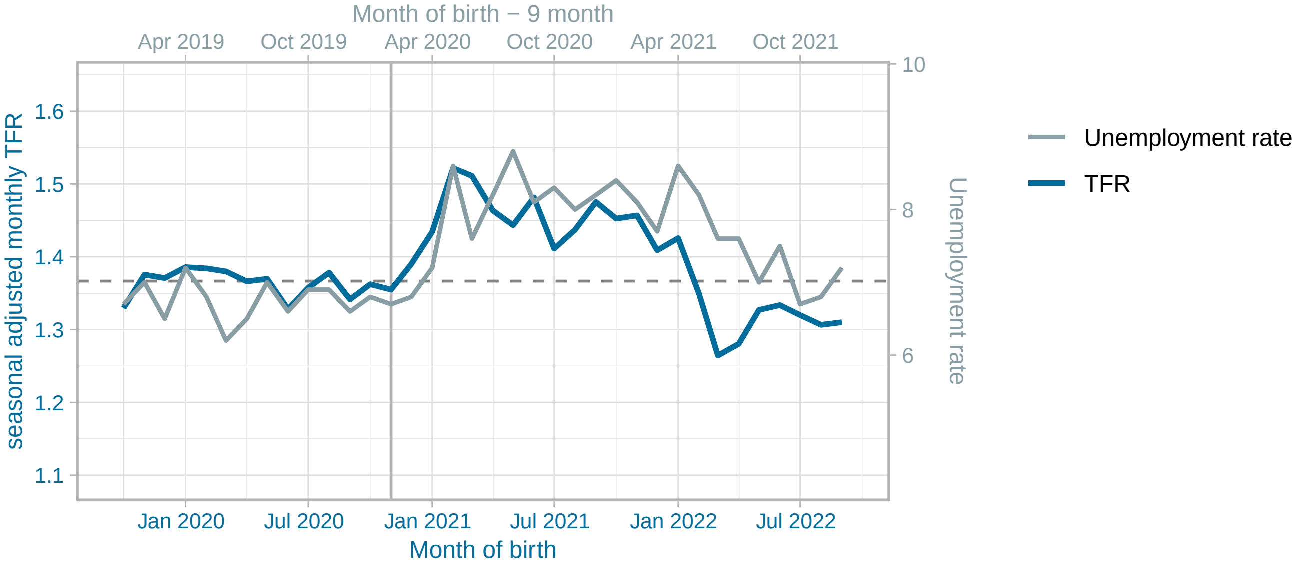


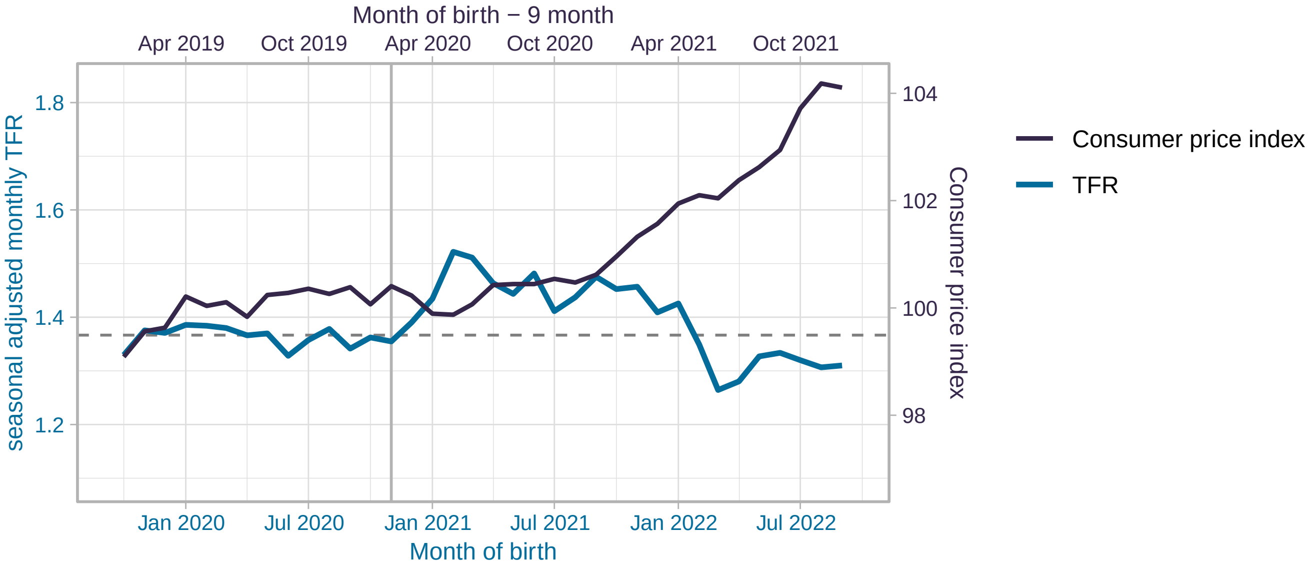


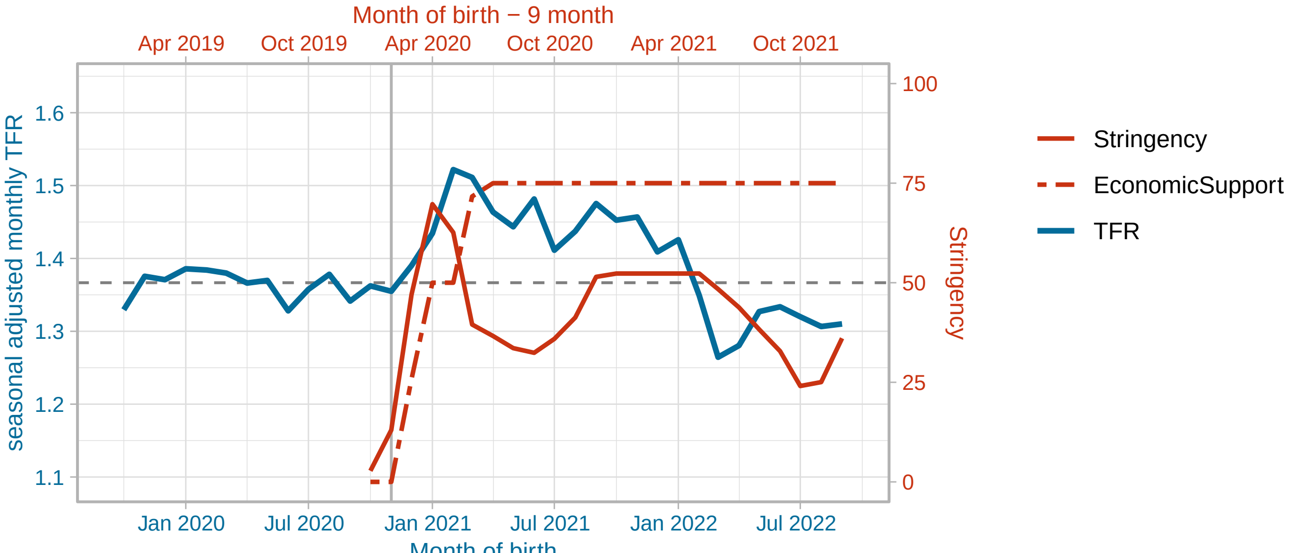


(figure continued on next page)

Finland

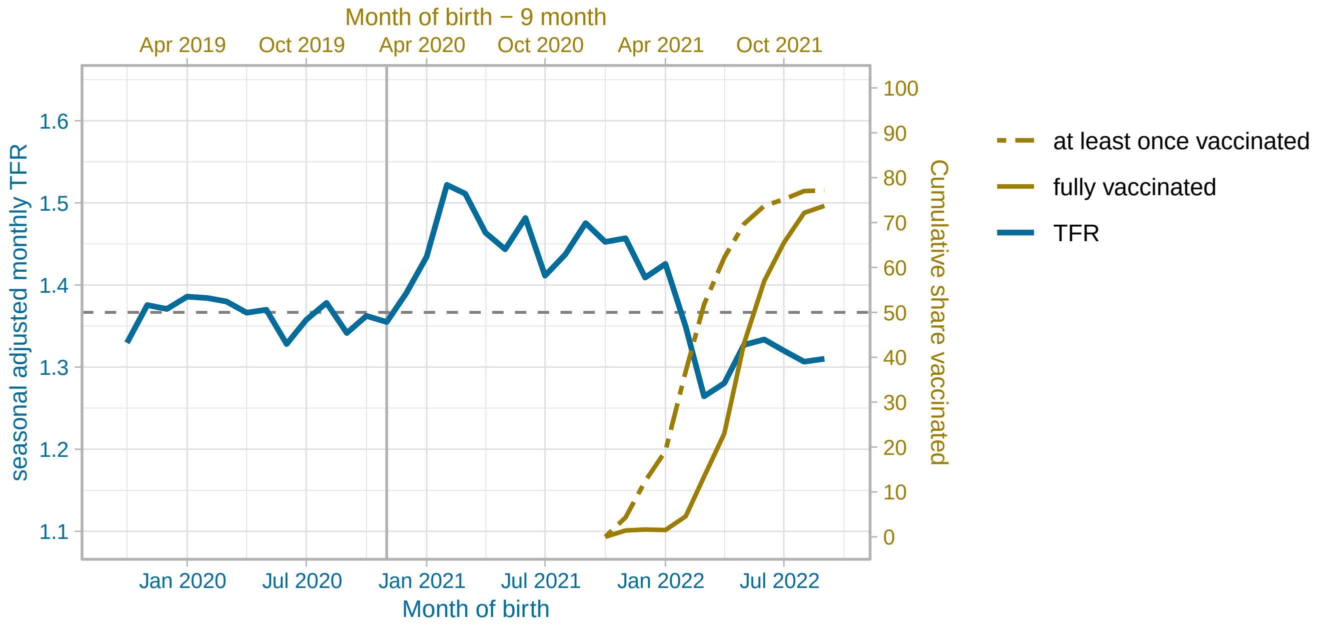


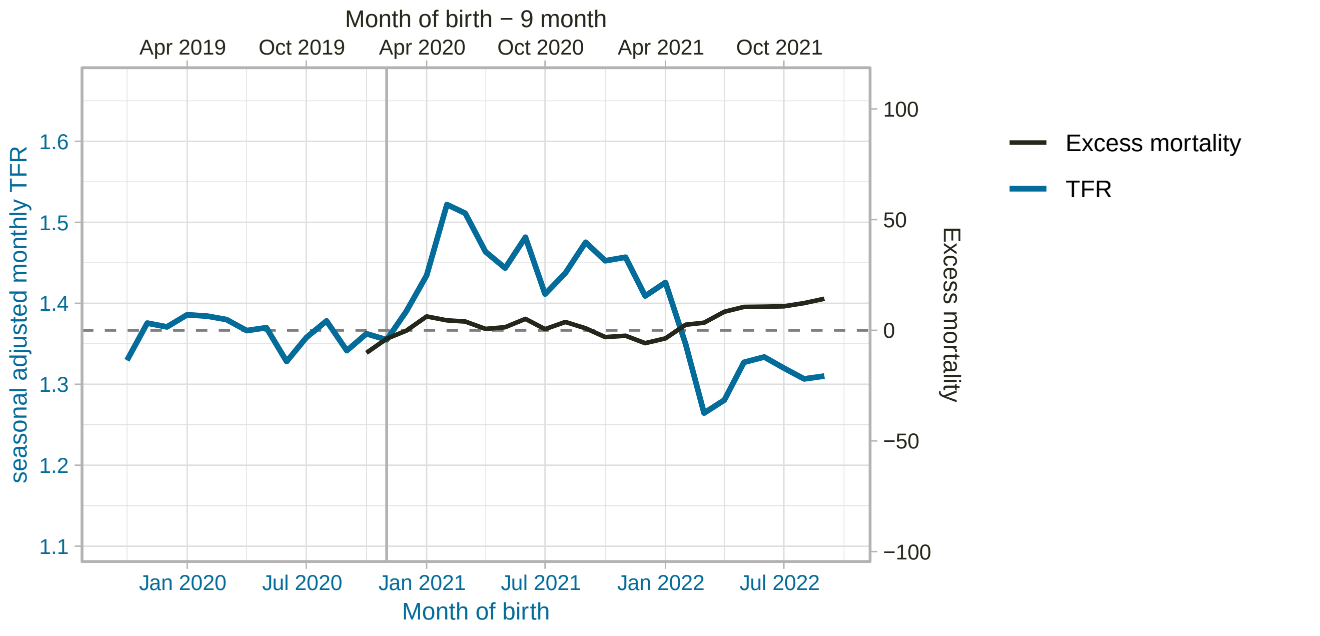


(figure continued on next page)

#### France

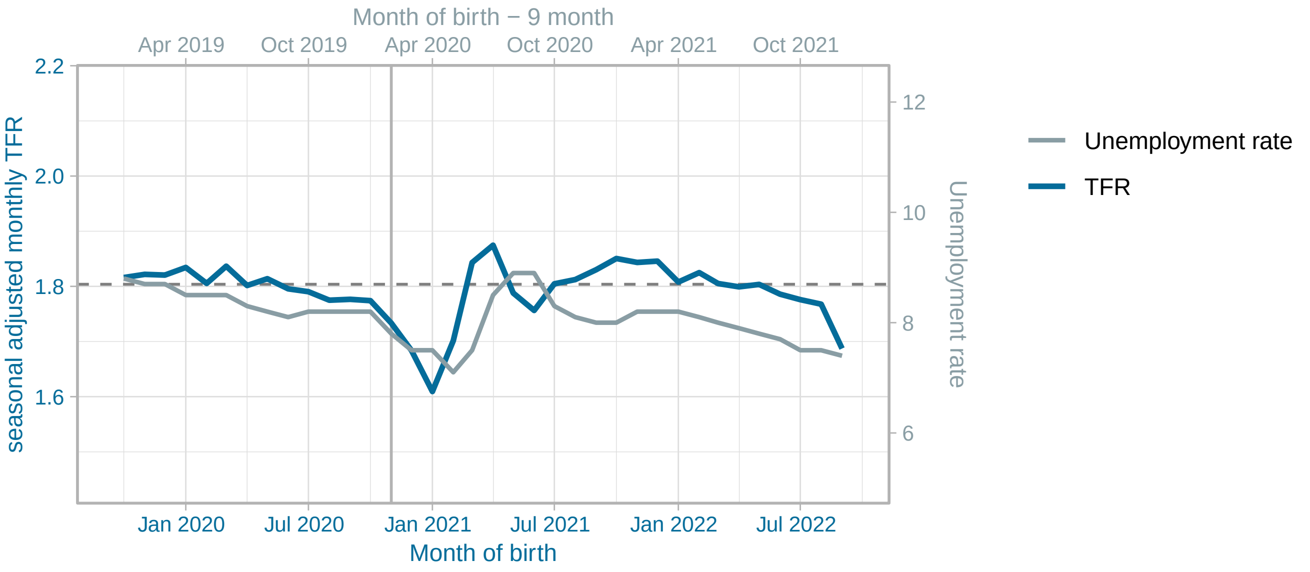


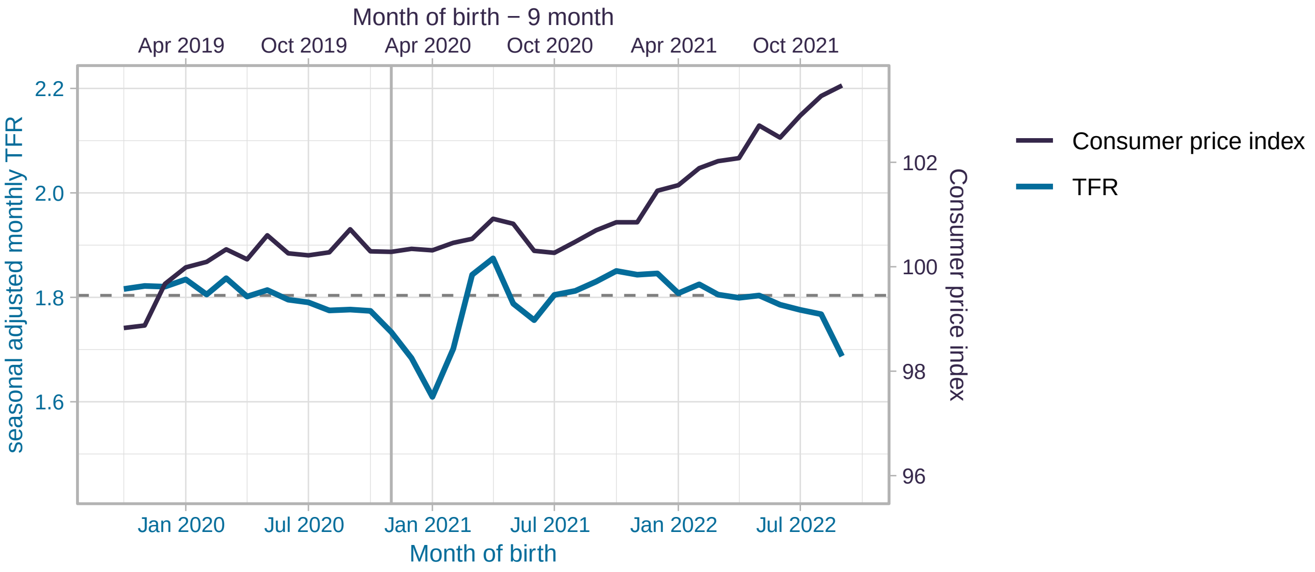


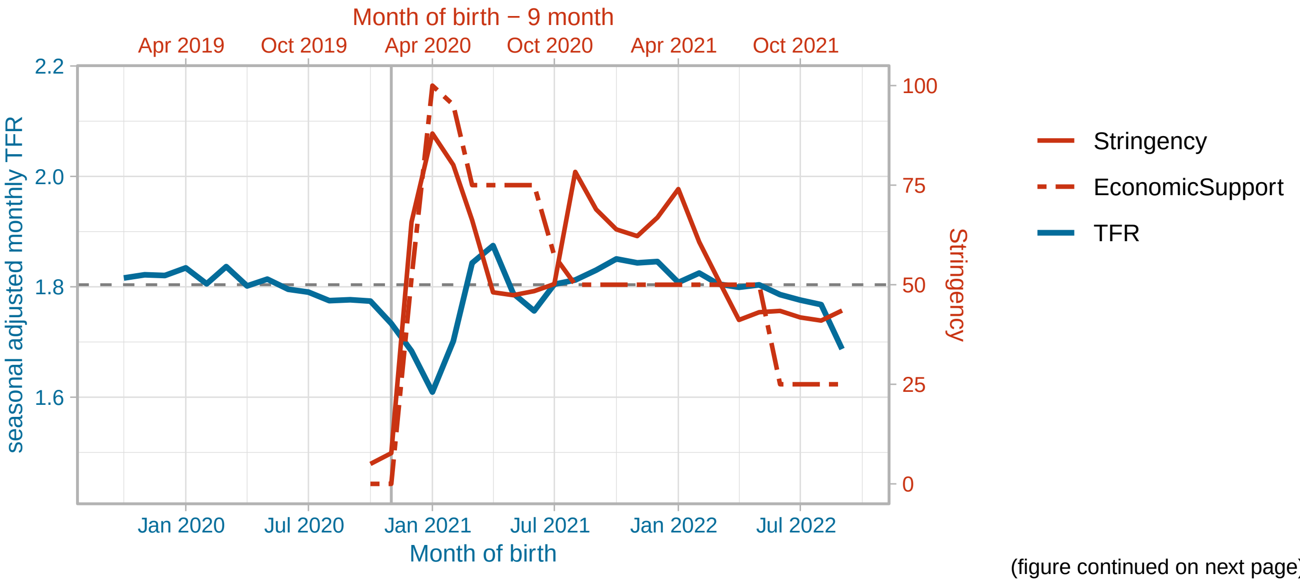


(figure continued on next page)

France

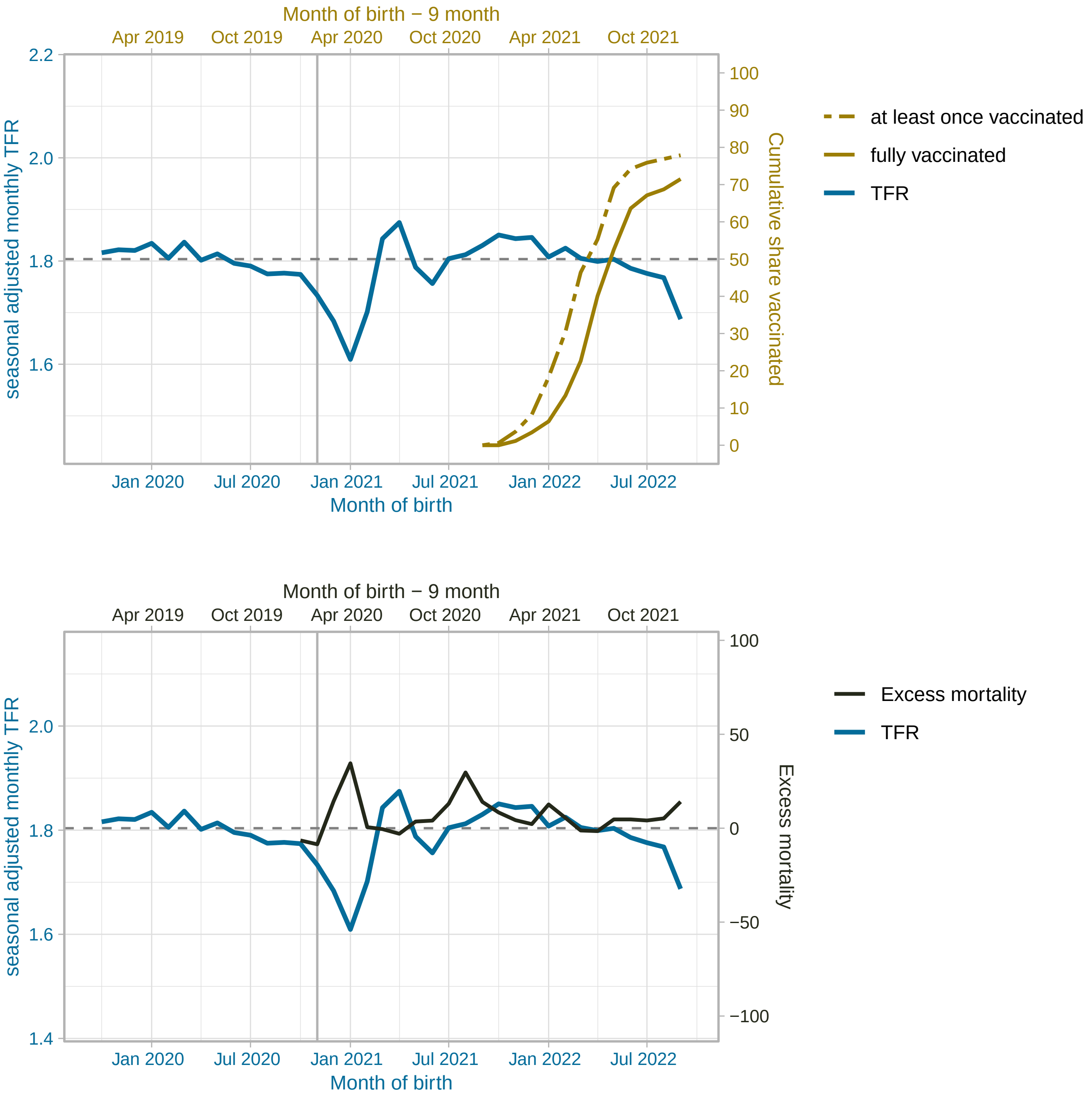


(figure continued on next page)

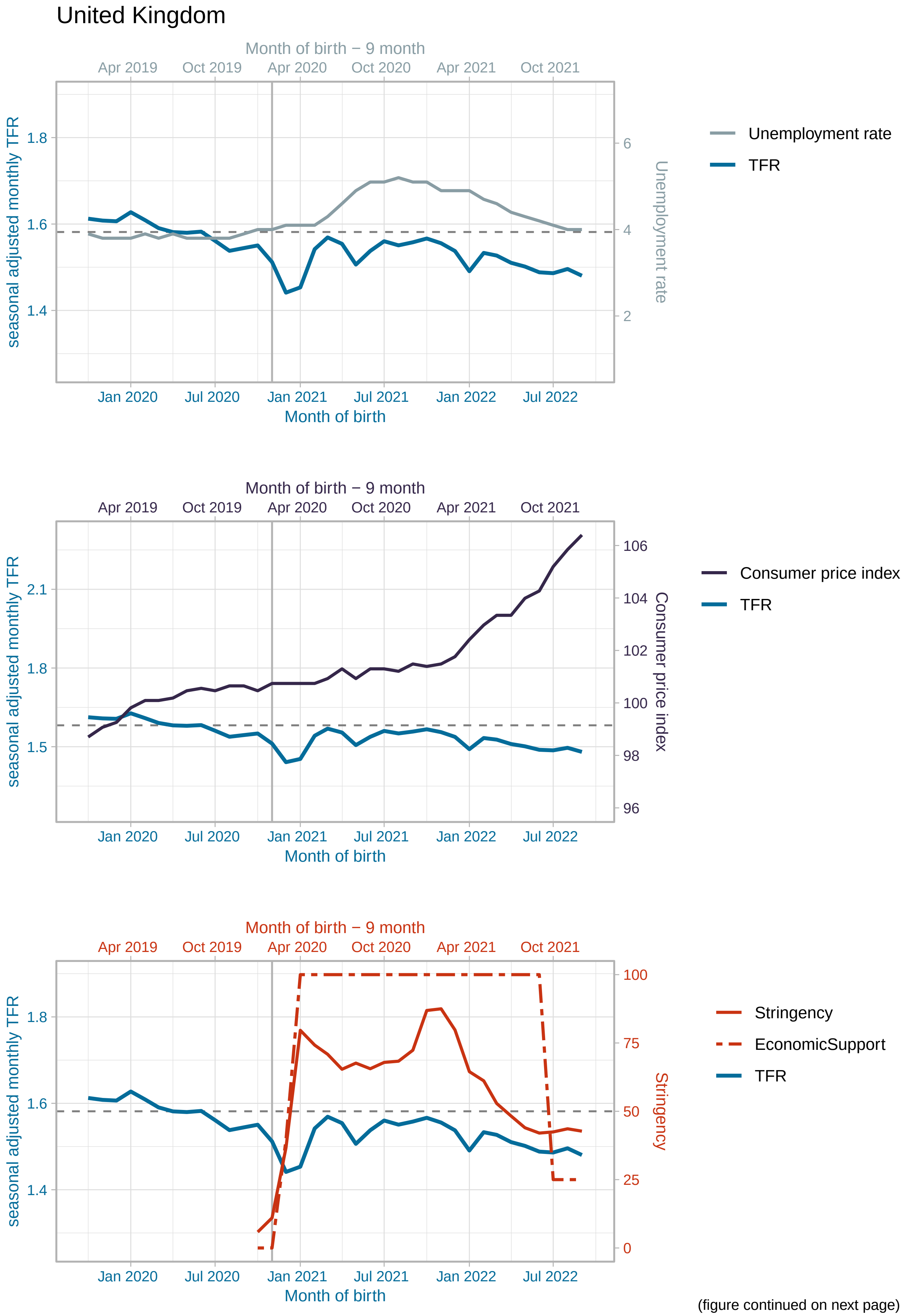


(figure continued on next page)

#### United Kingdom

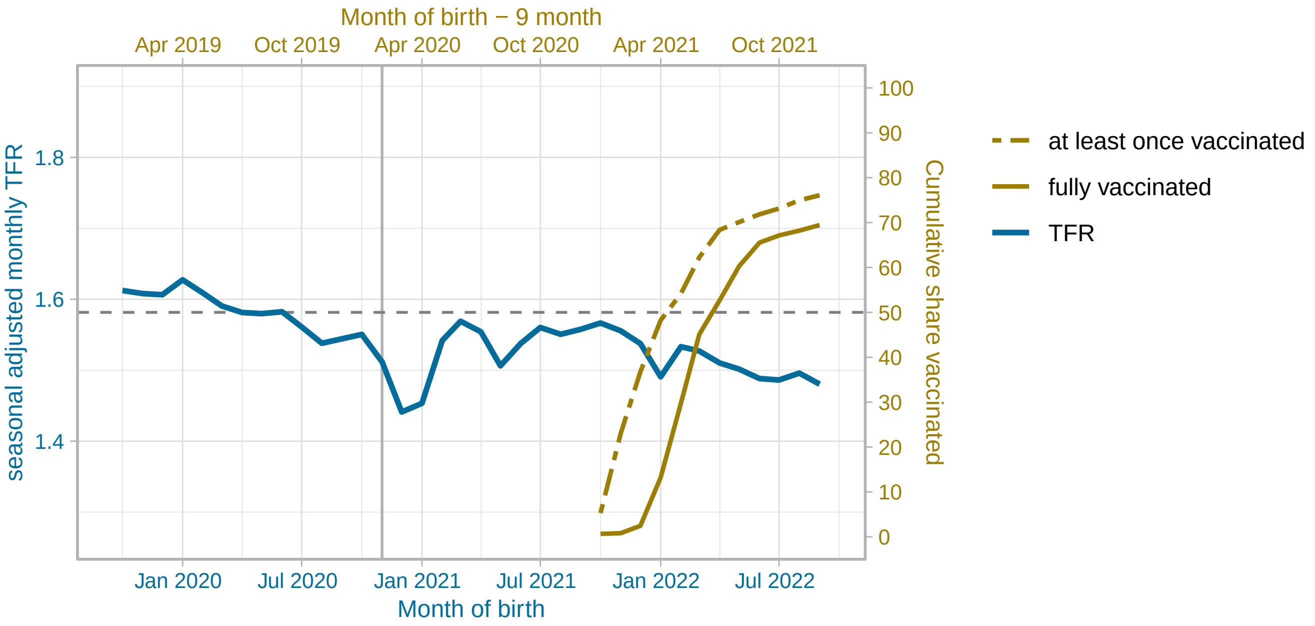


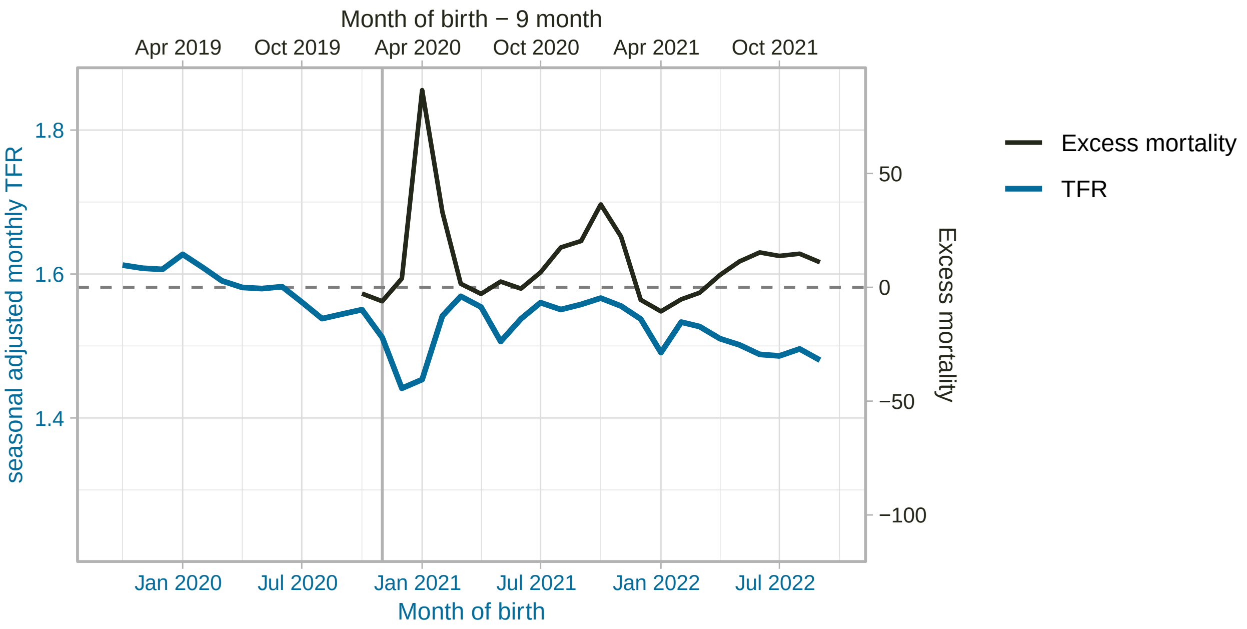


(figure continued on next page)

#### Greece

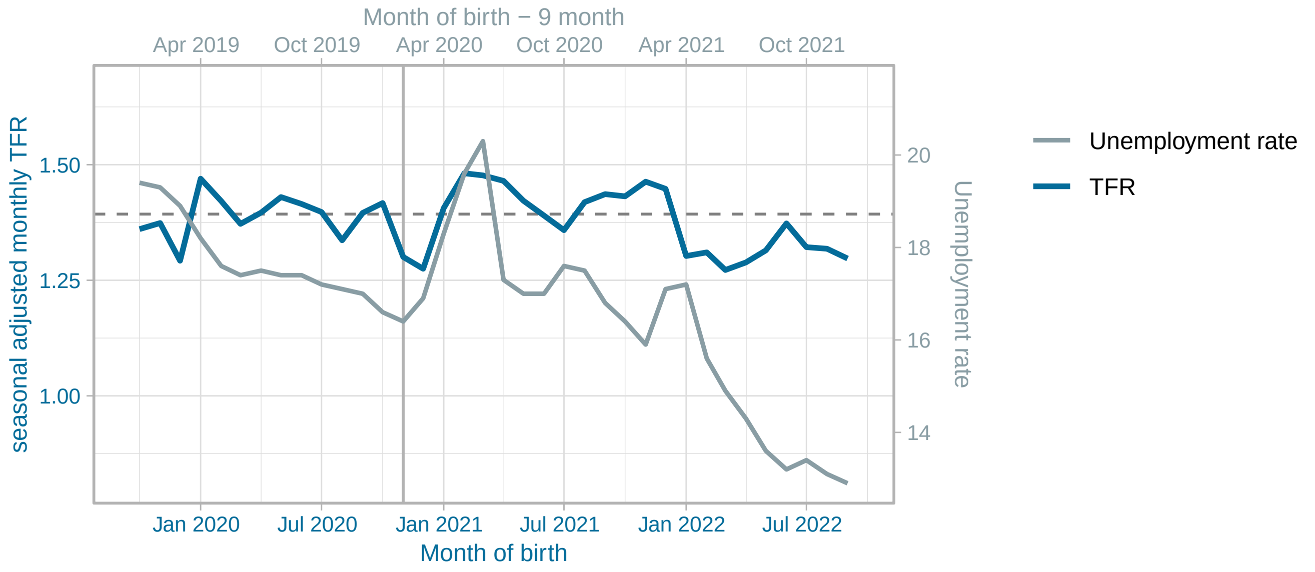


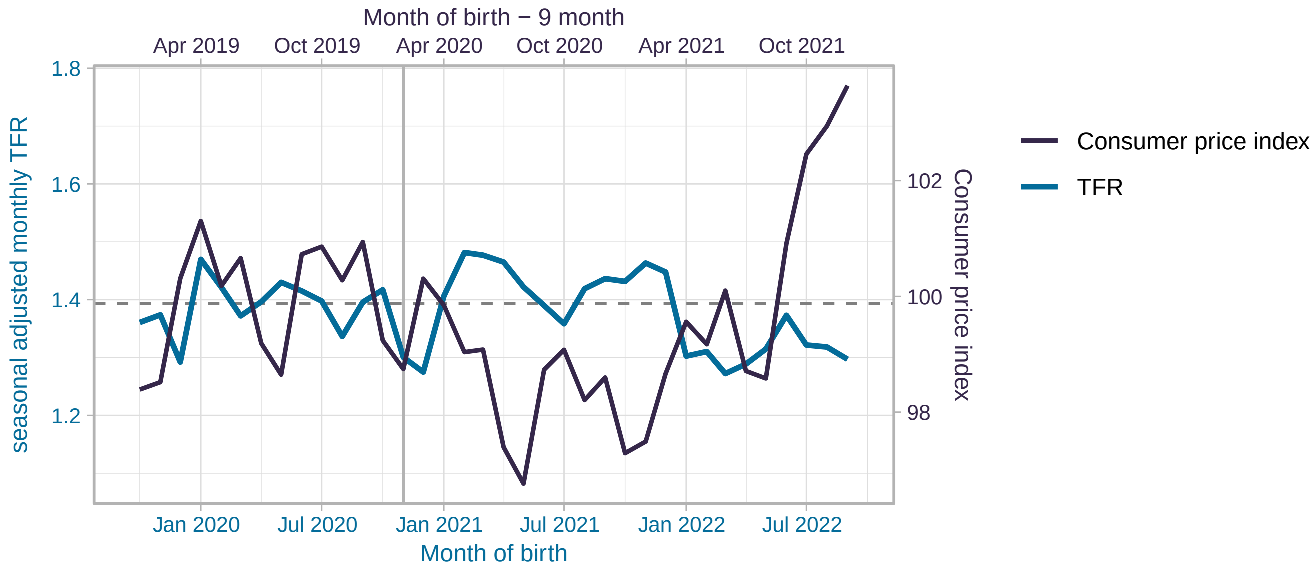


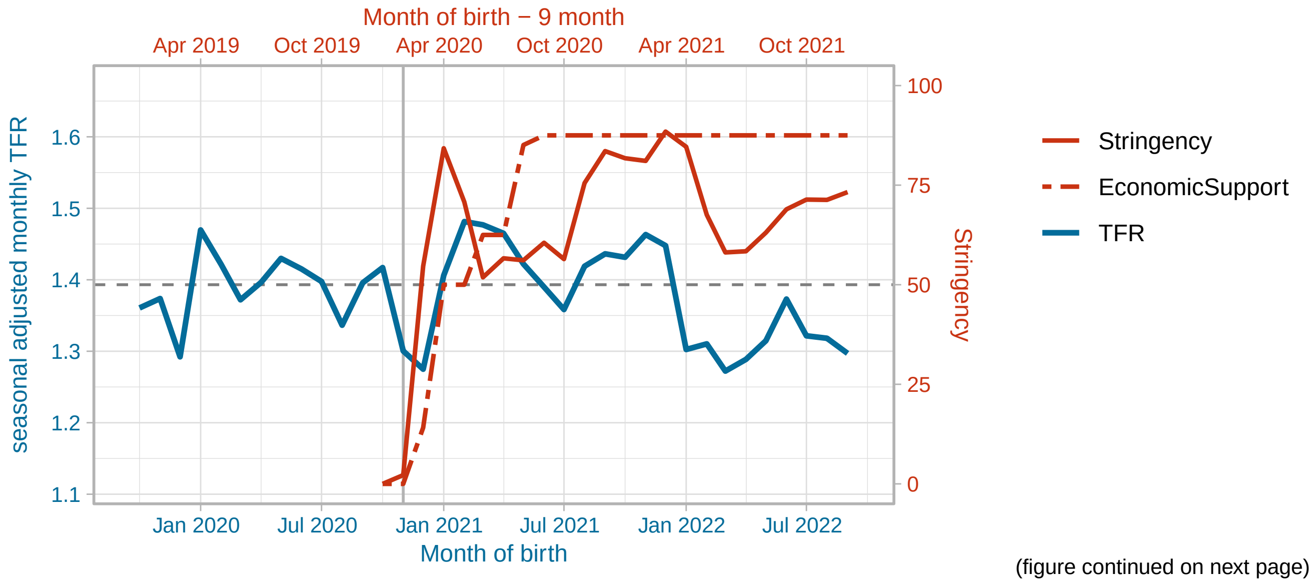


(figure continued on next page)

Greece

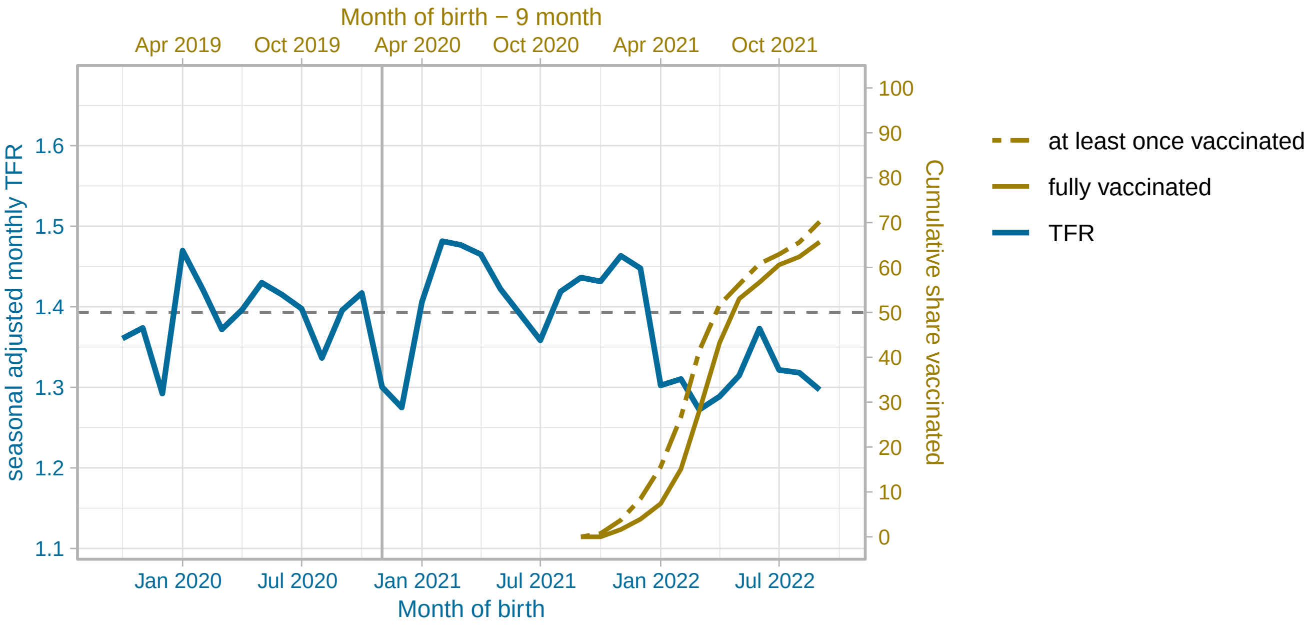


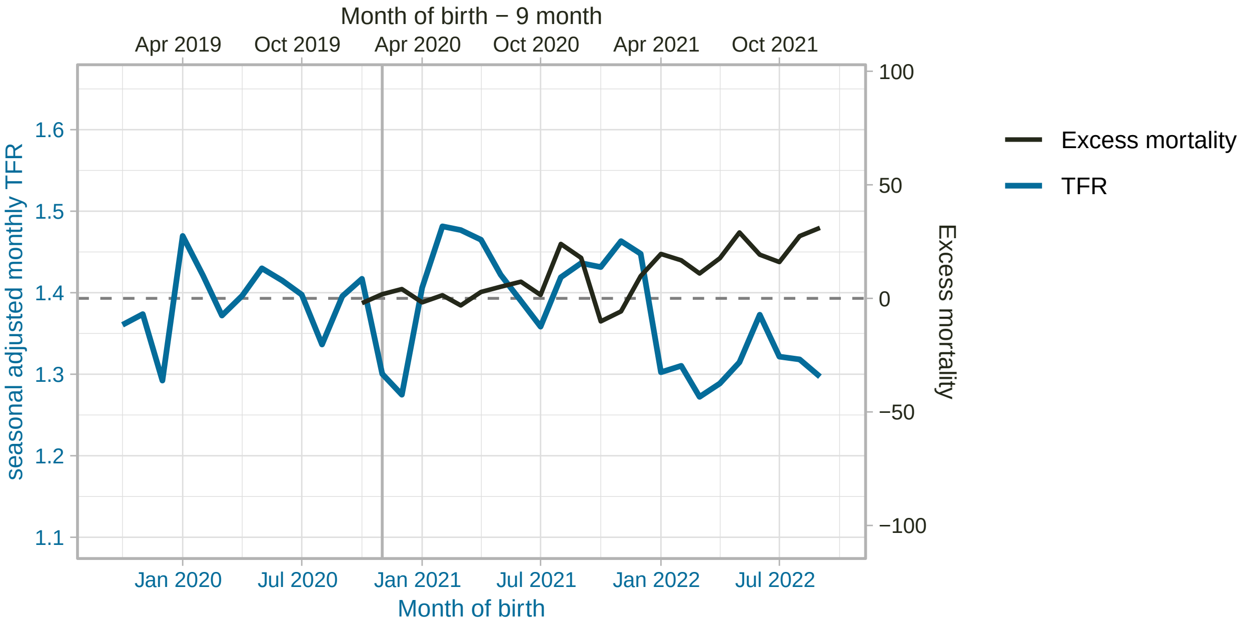


(figure continued on next page)

#### Hungary

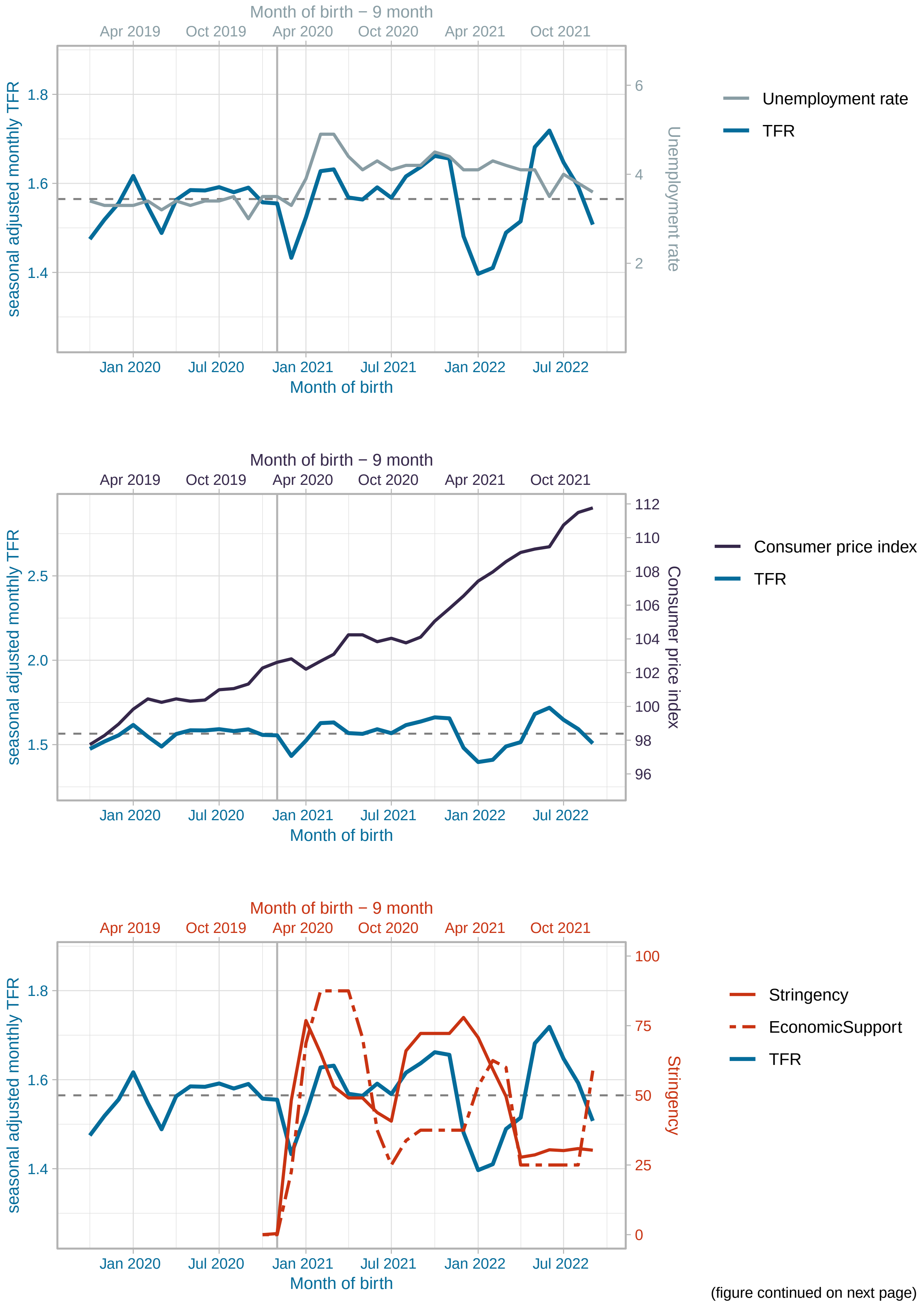


(figure continued on next page)

Hungary

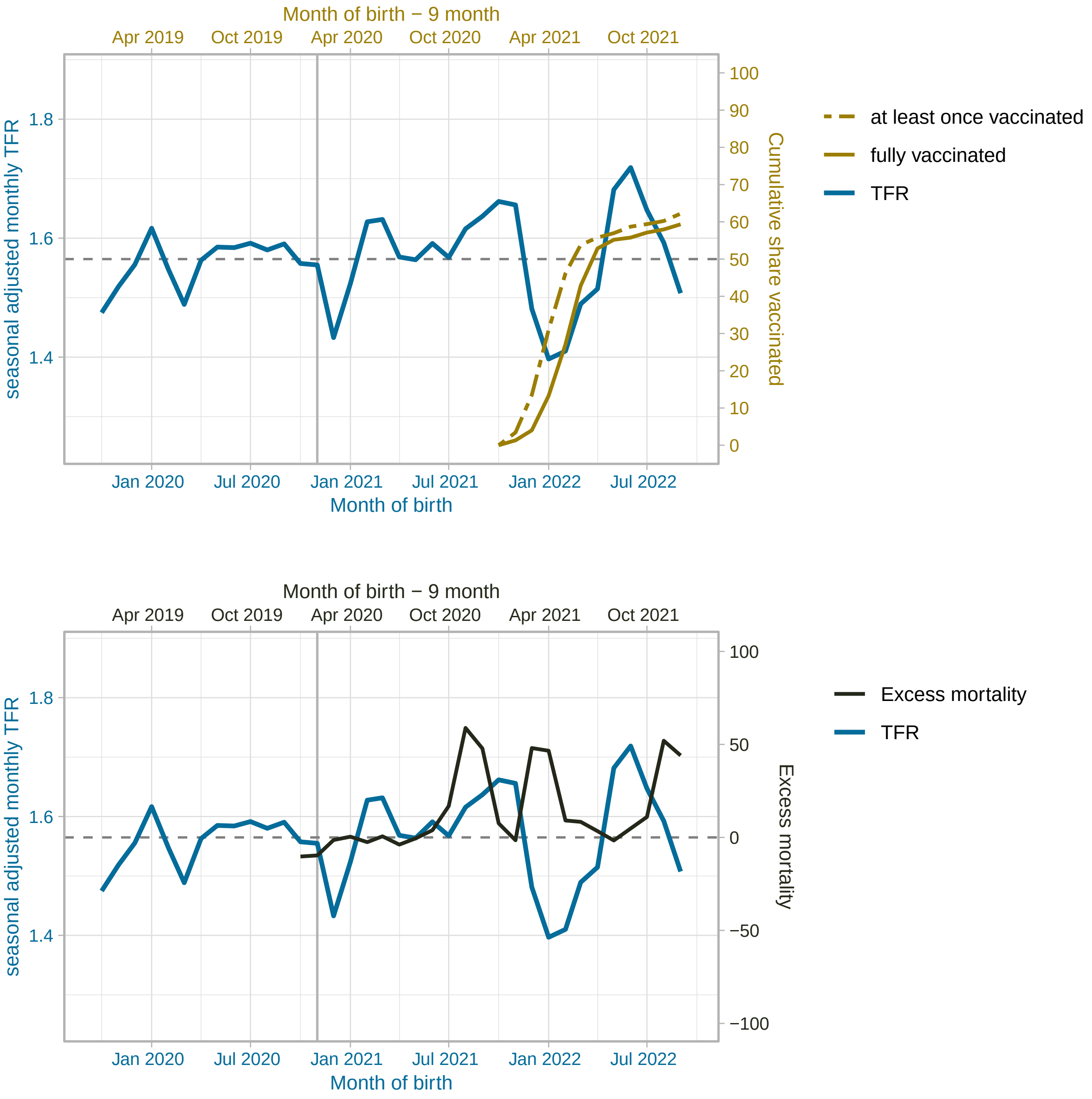


(figure continued on next page)

#### Ireland

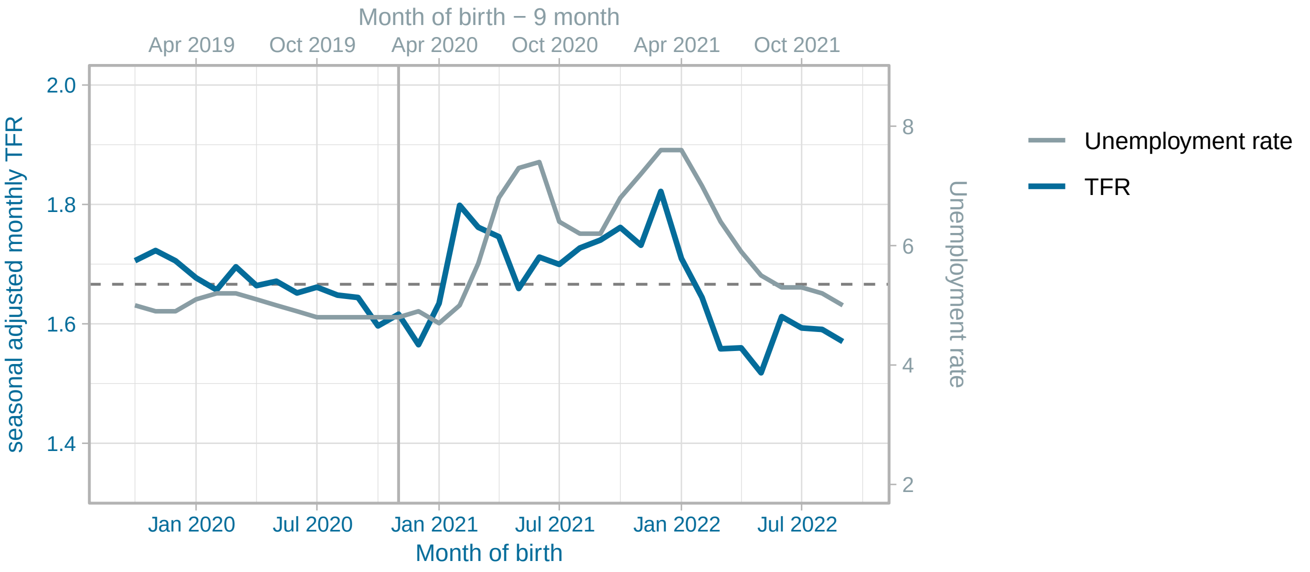


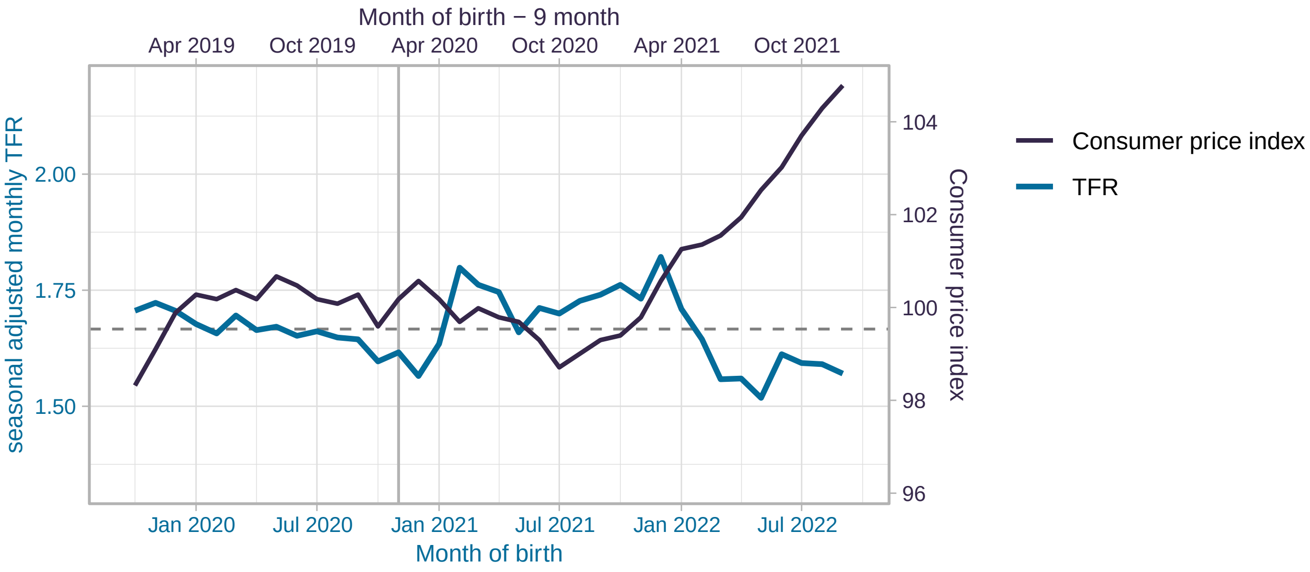


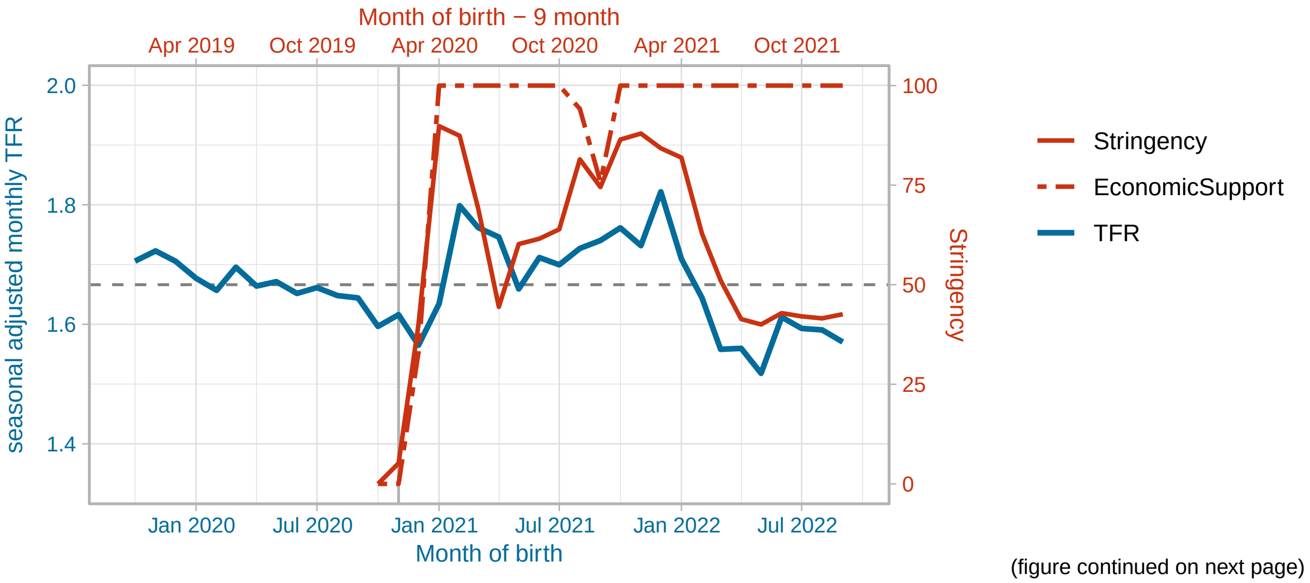


(figure continued on next page)

#### Ireland

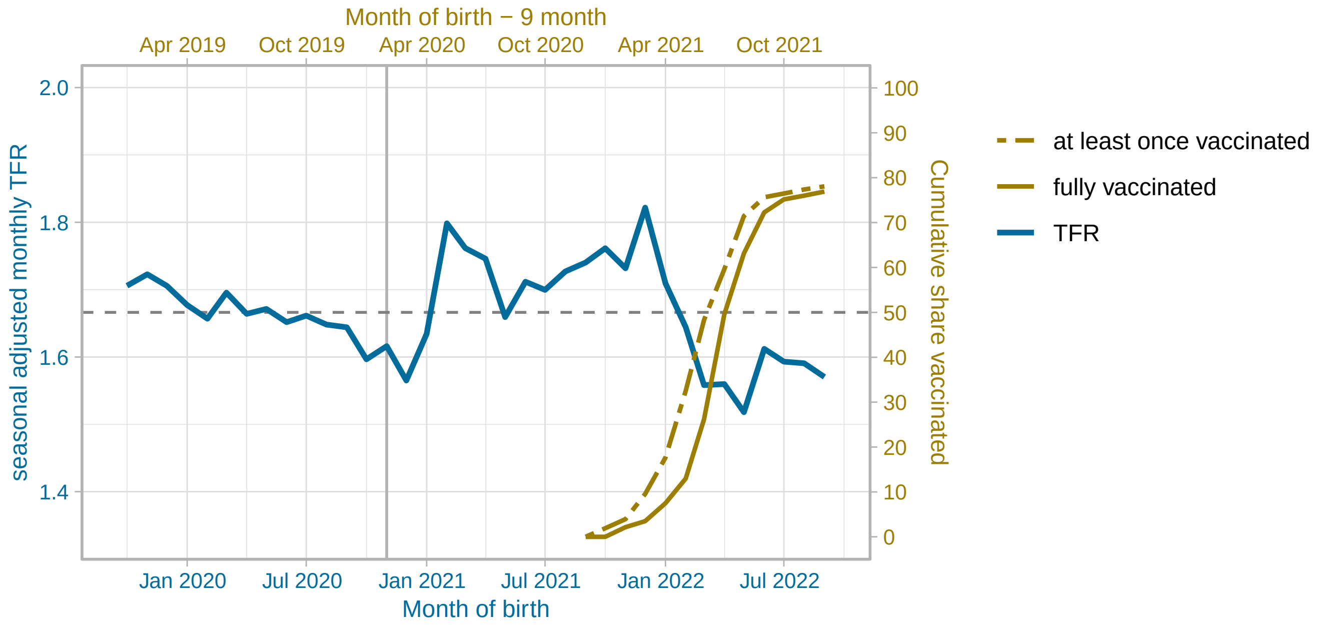


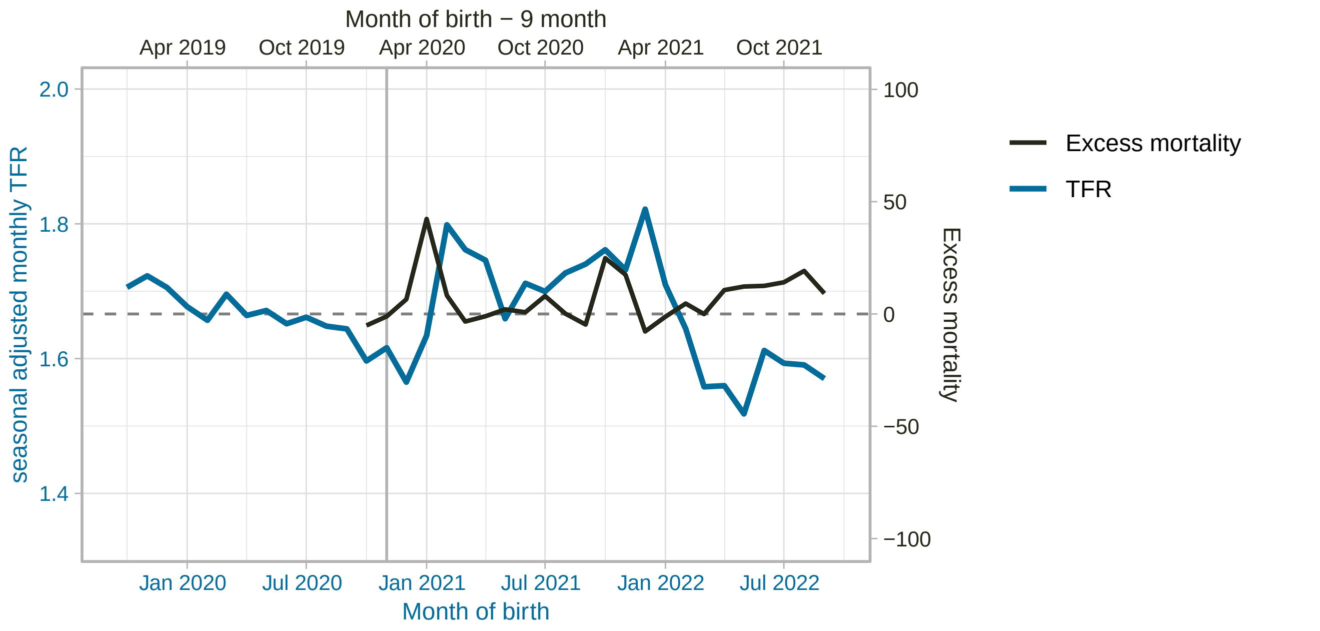


(figure continued on next page)

Israel

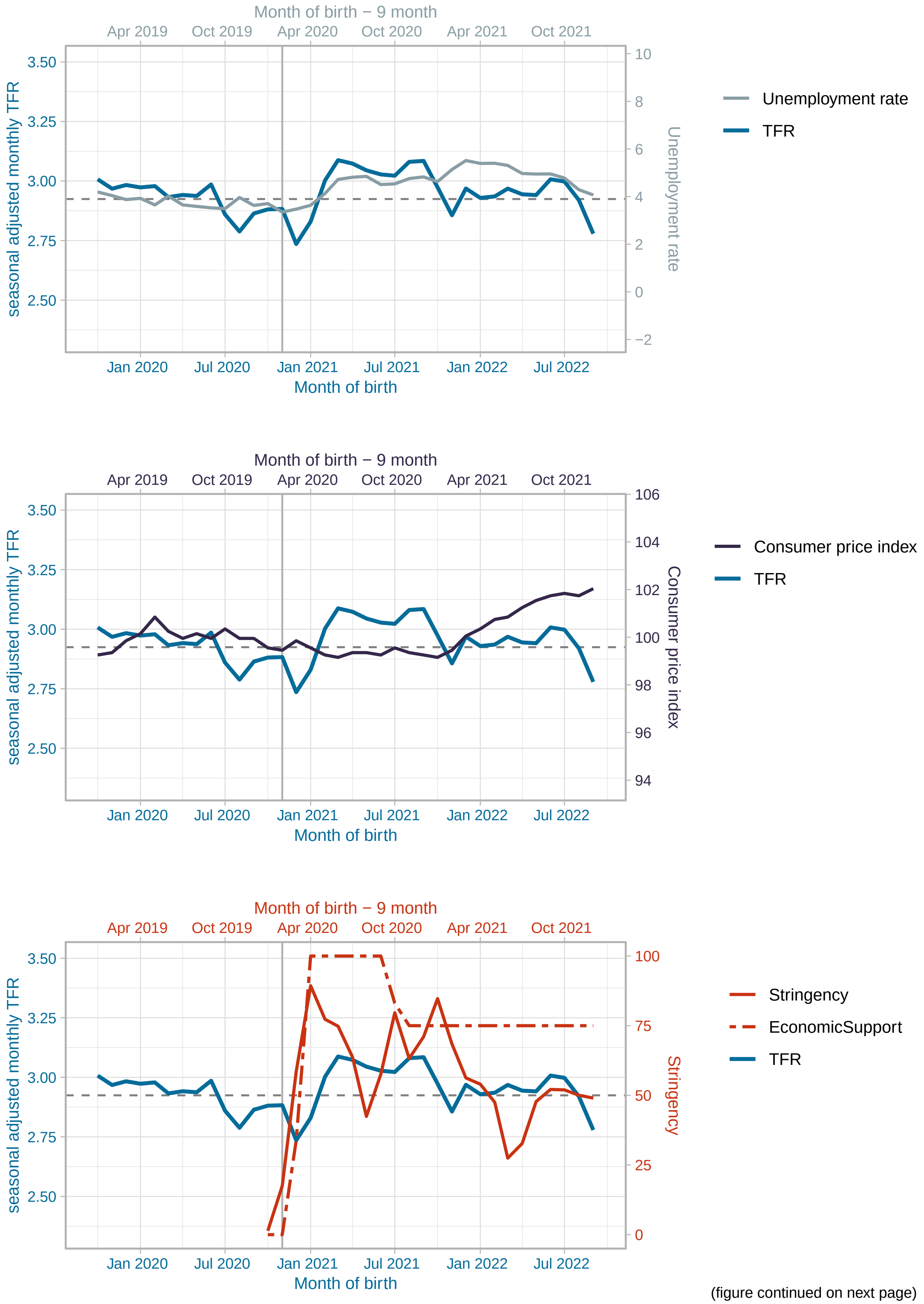


(figure continued on next page)

Israel

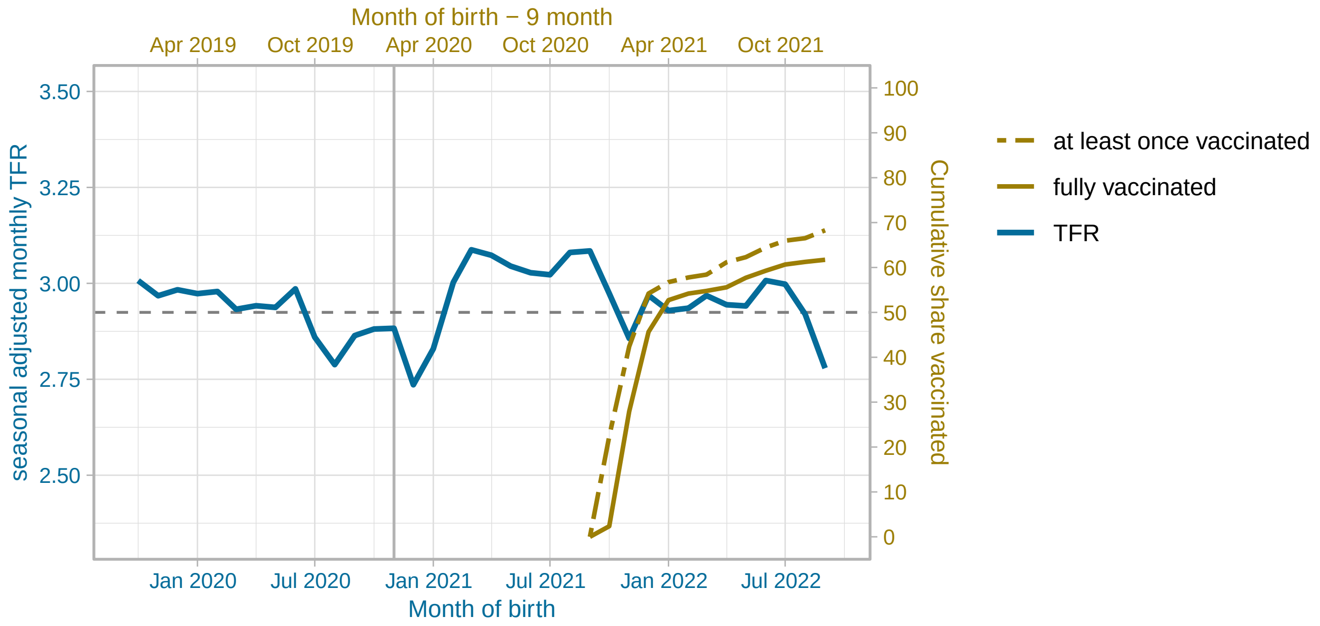


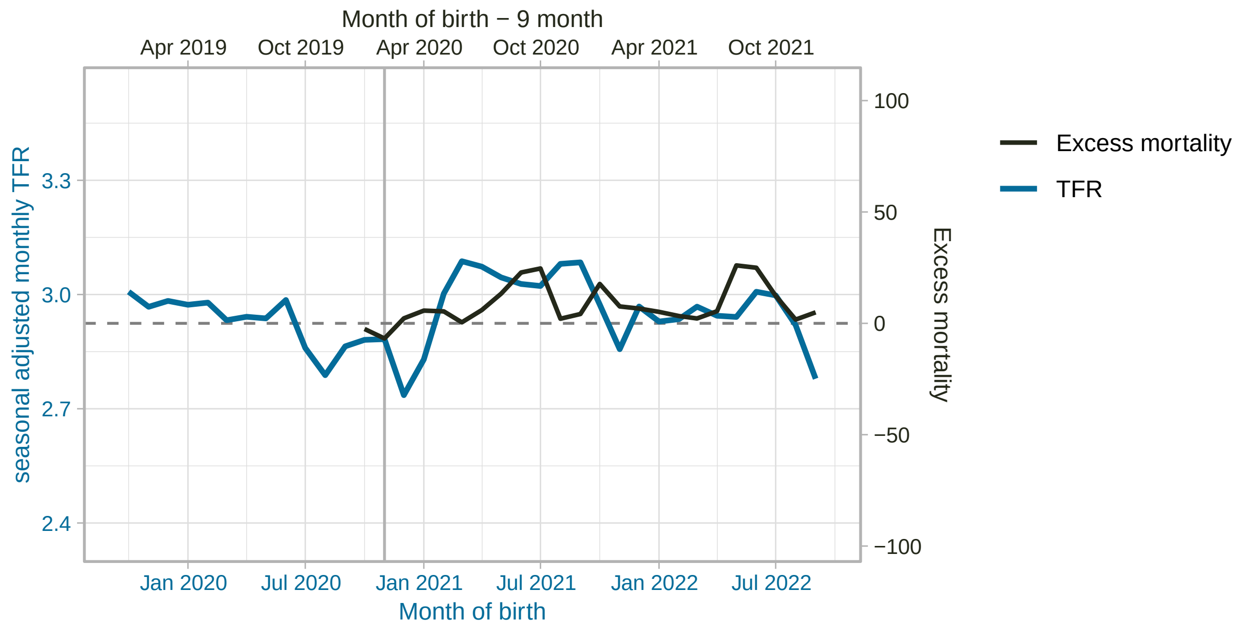


(figure continued on next page)

#### Italy

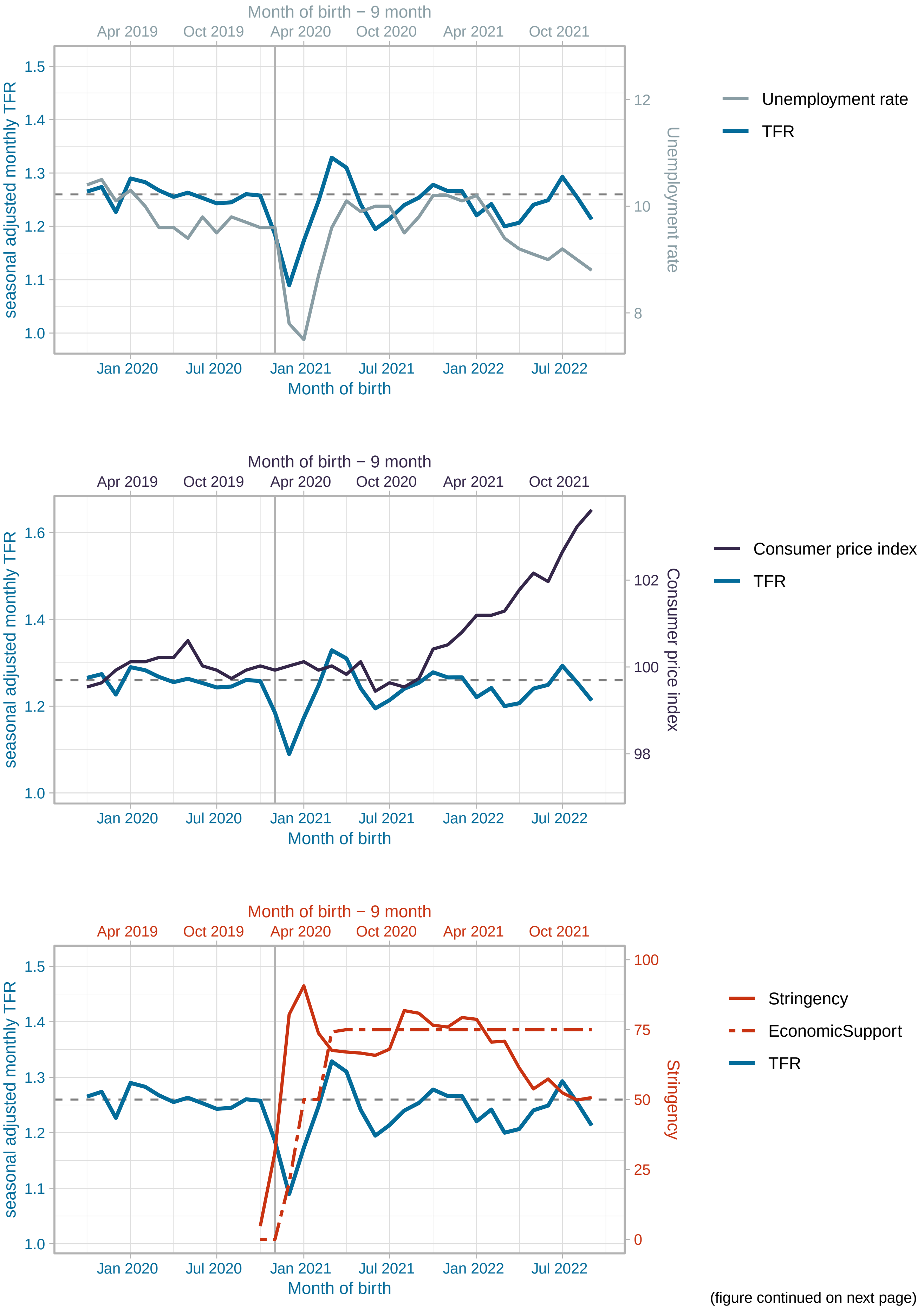


(figure continued on next page)

Italy

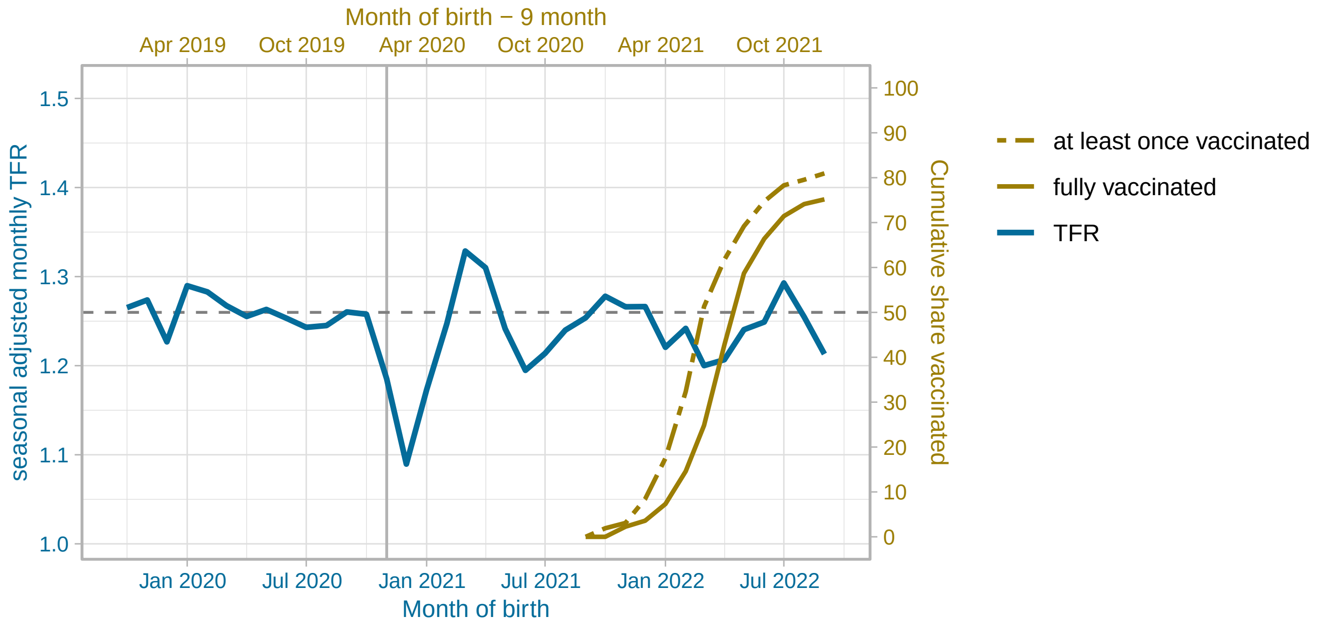


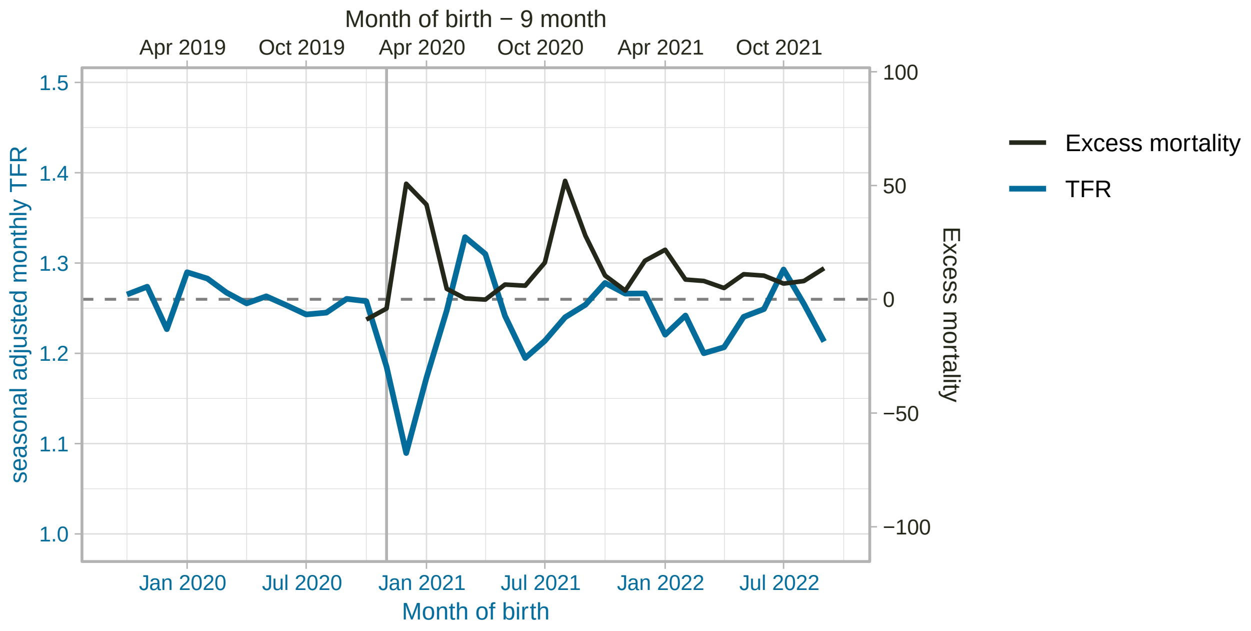


(figure continued on next page)

Japan

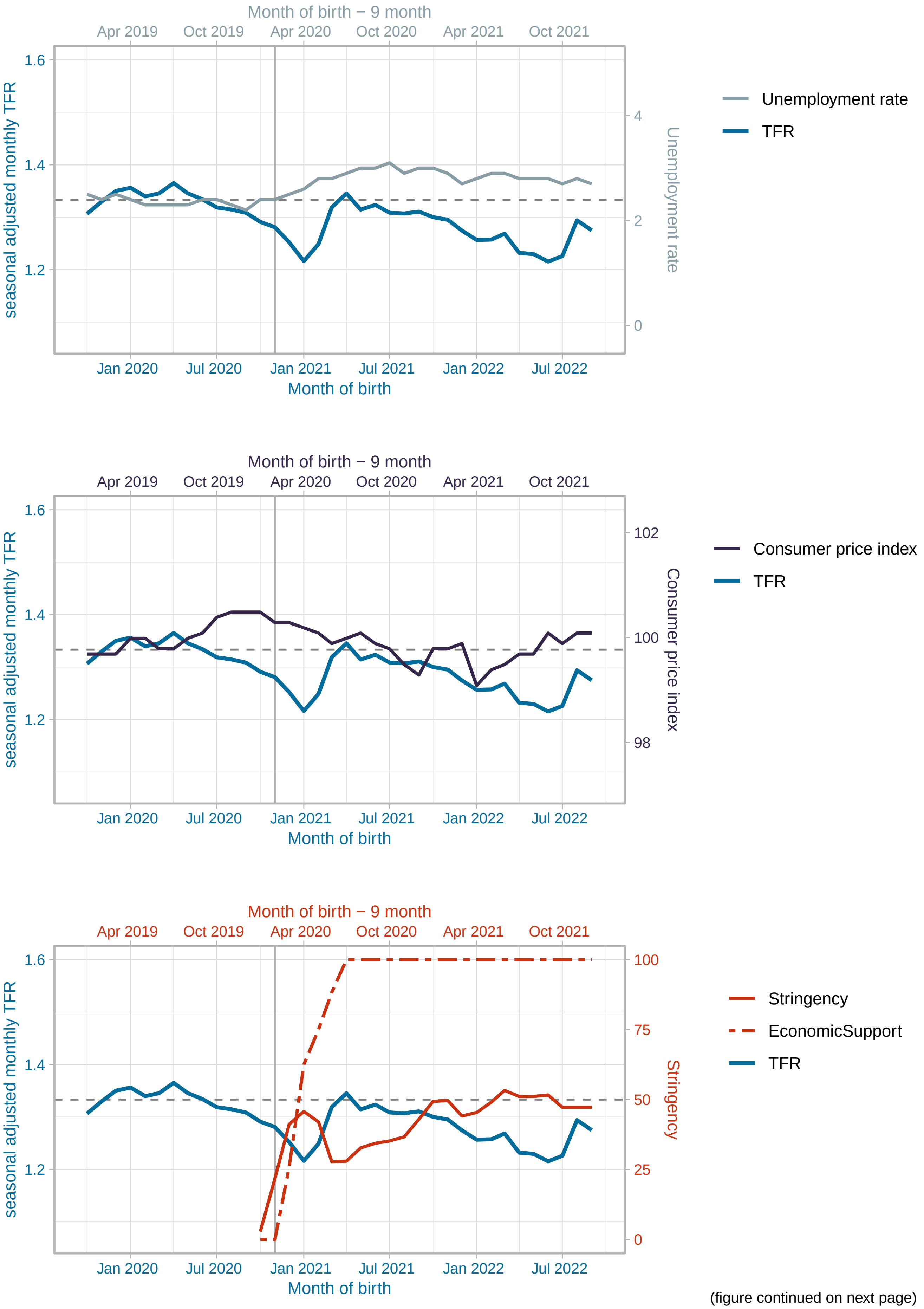


(figure continued on next page)

### Japan

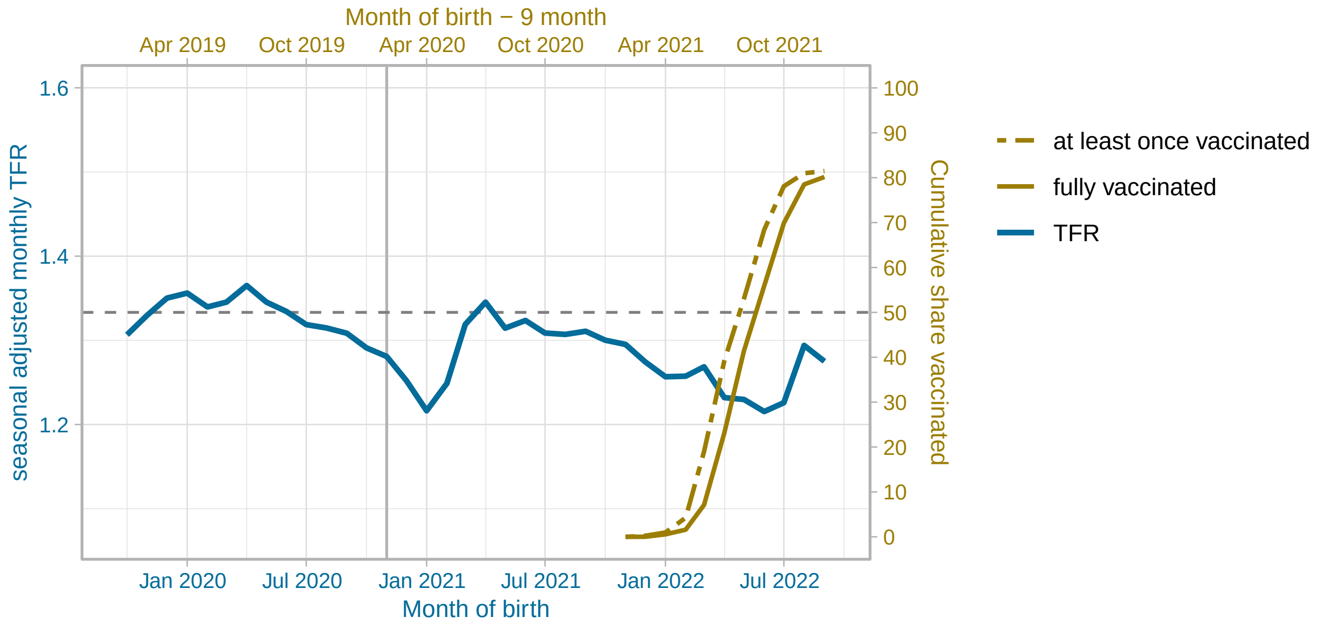


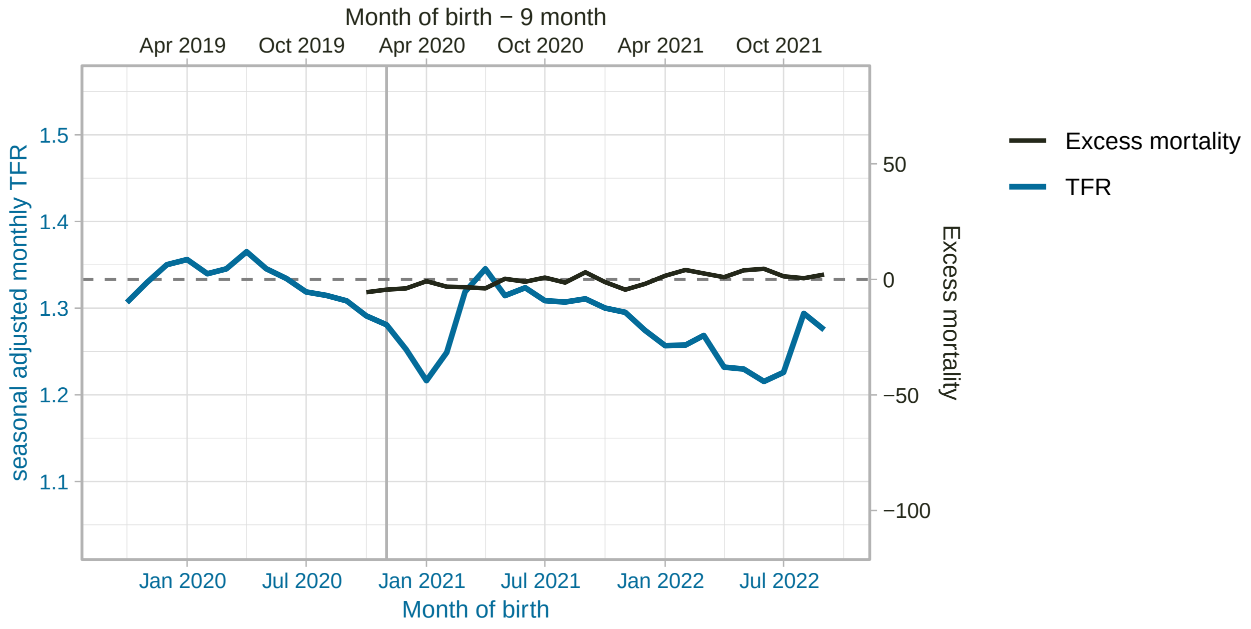


(figure continued on next page)

#### South Korea

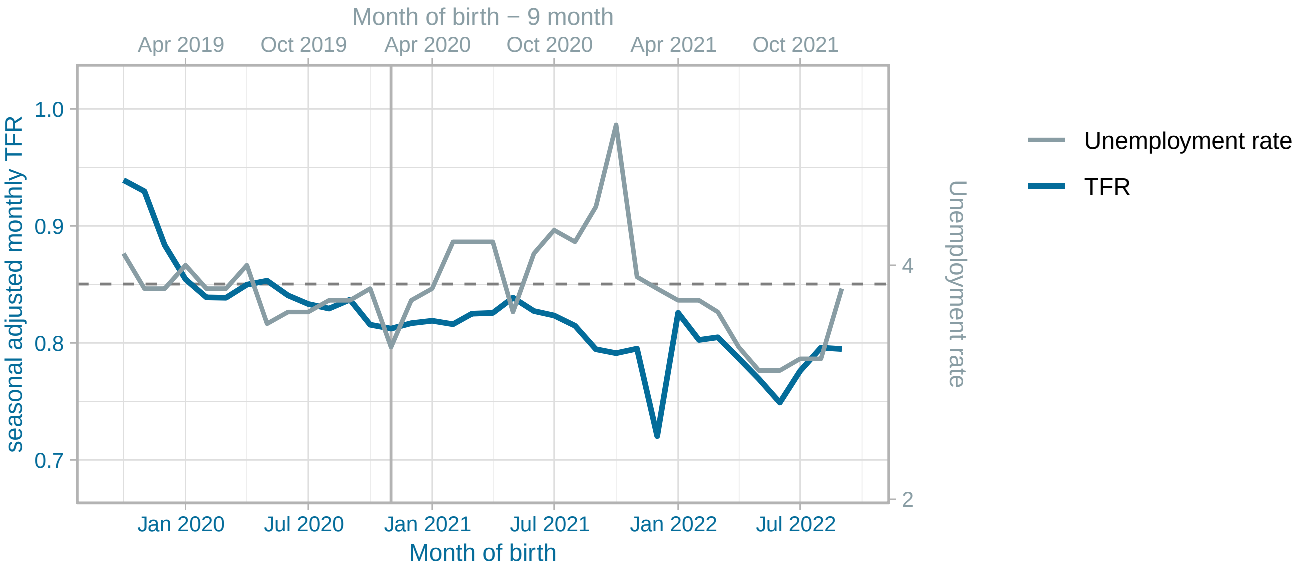


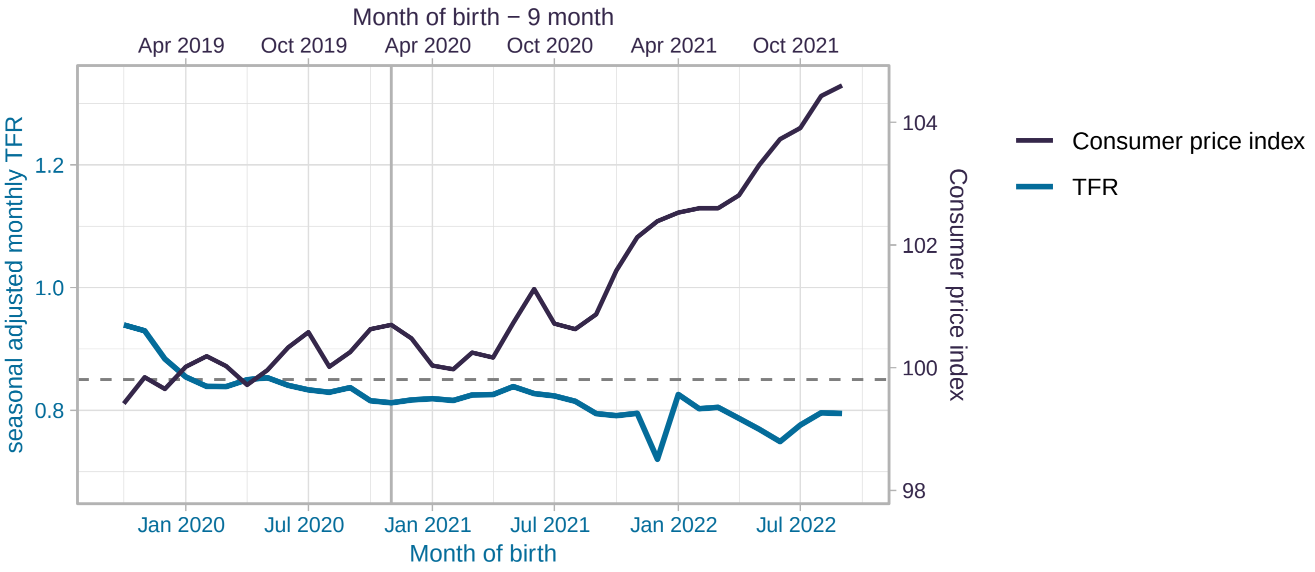


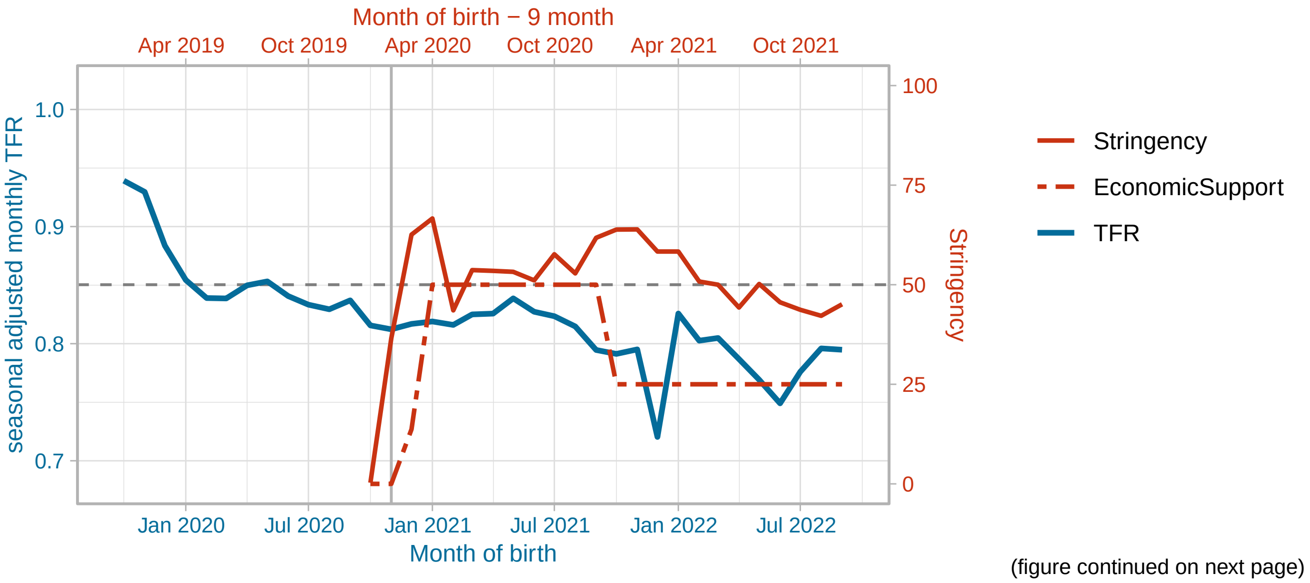


(figure continued on next page)

#### South Korea

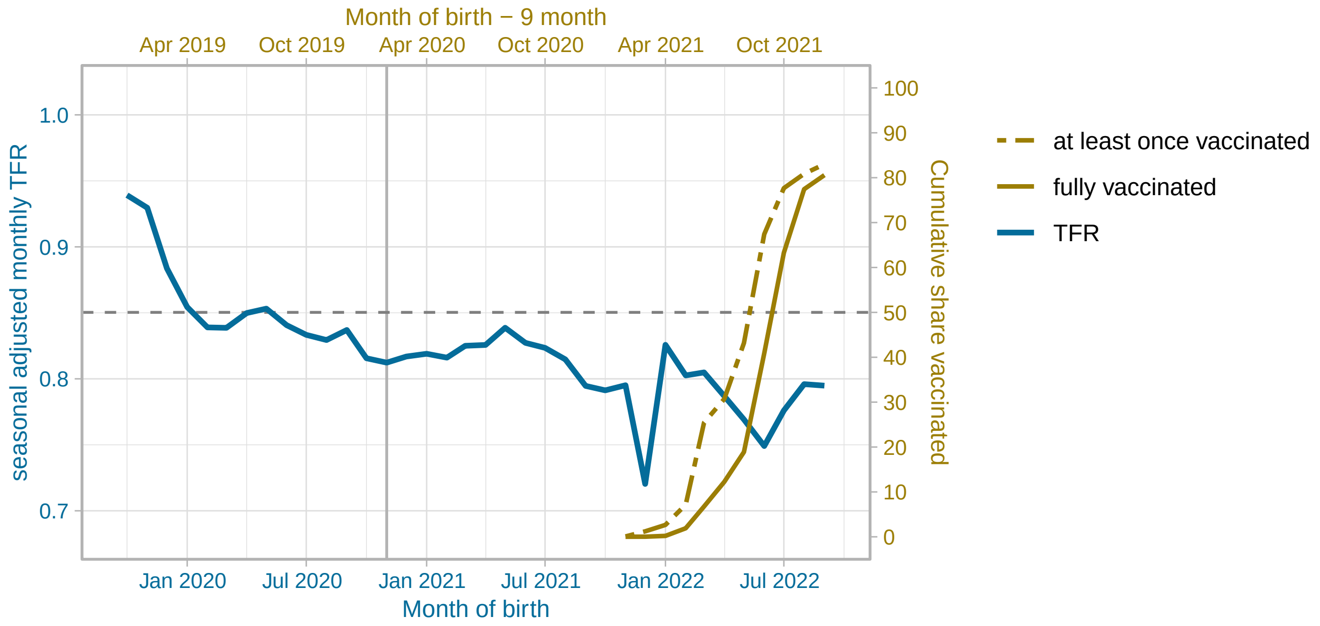


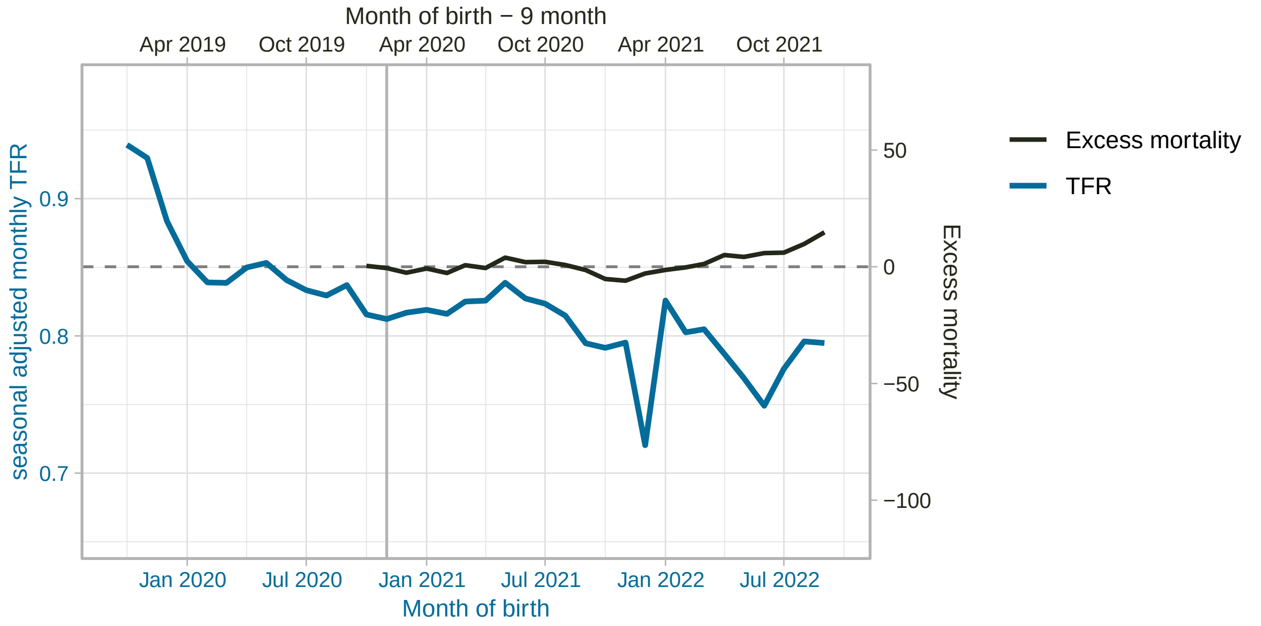


(figure continued on next page)

#### Lithuania

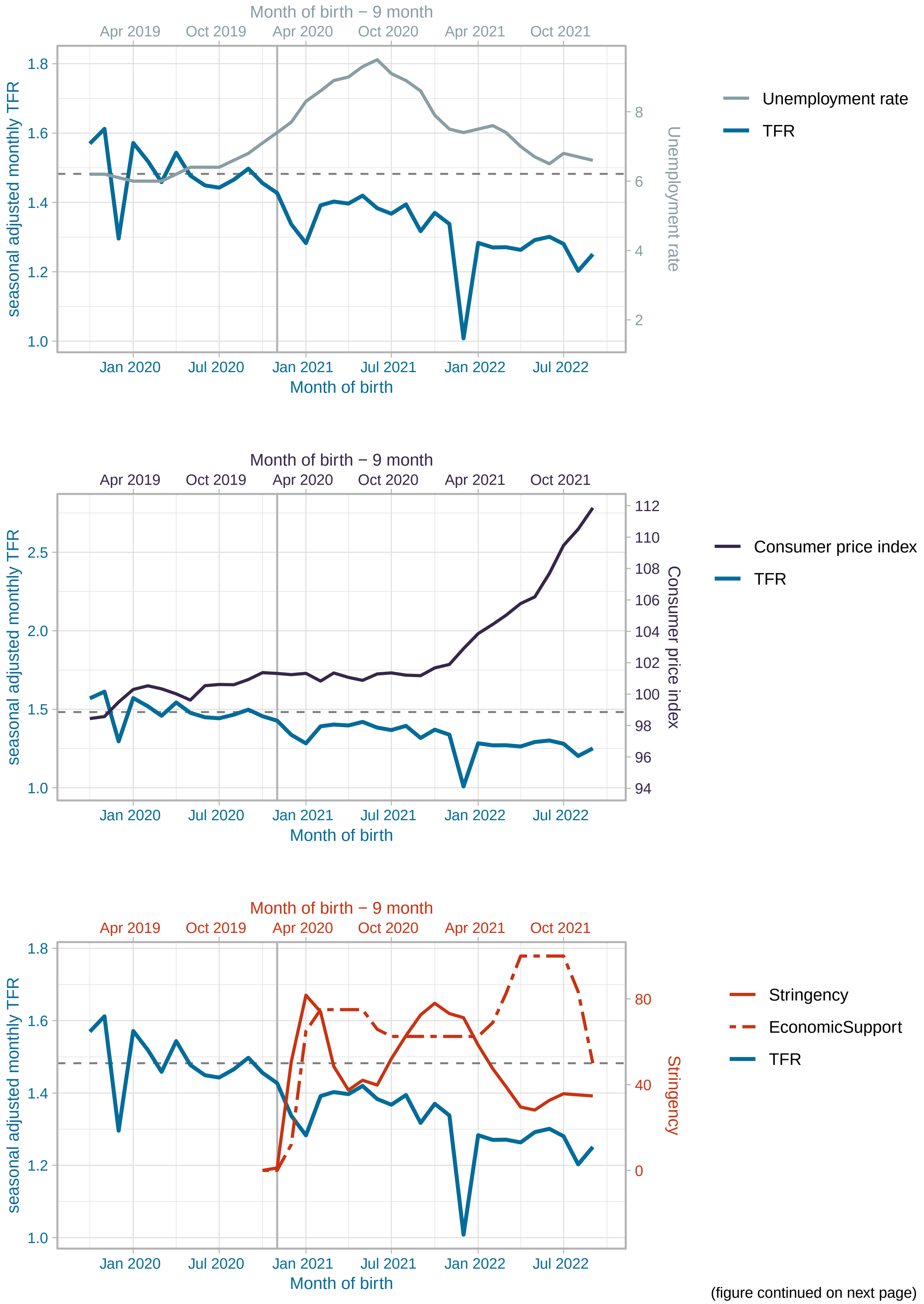


(figure continued on next page)

#### Lithuania

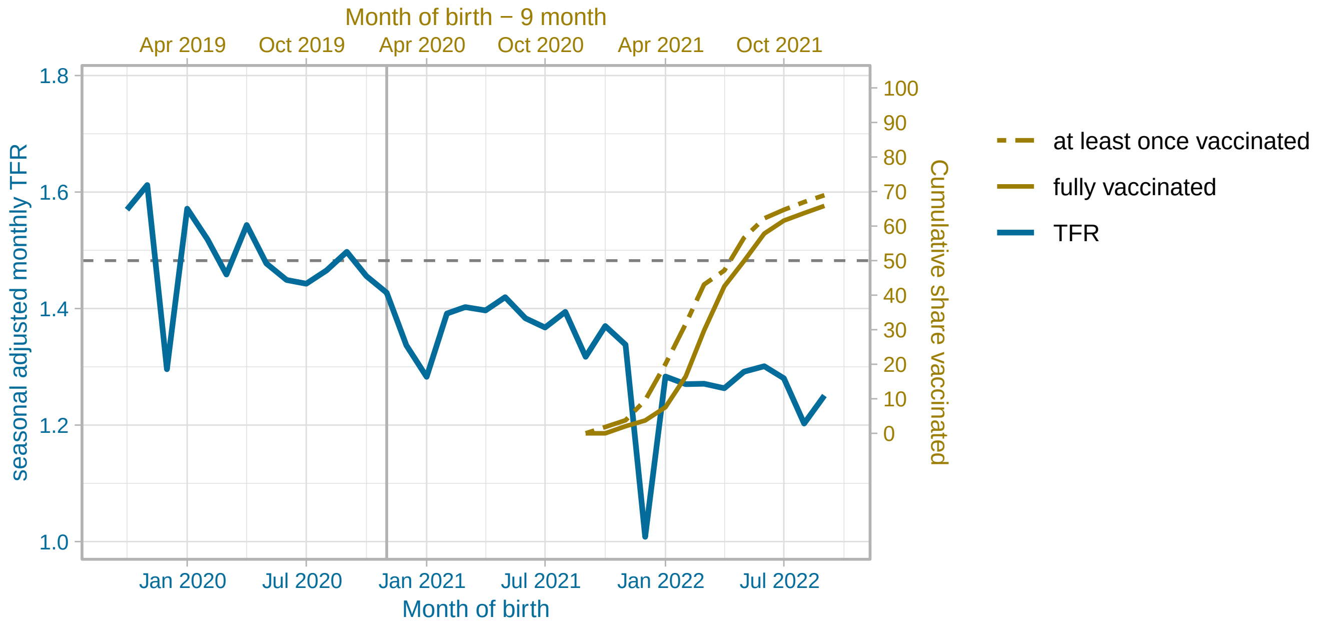


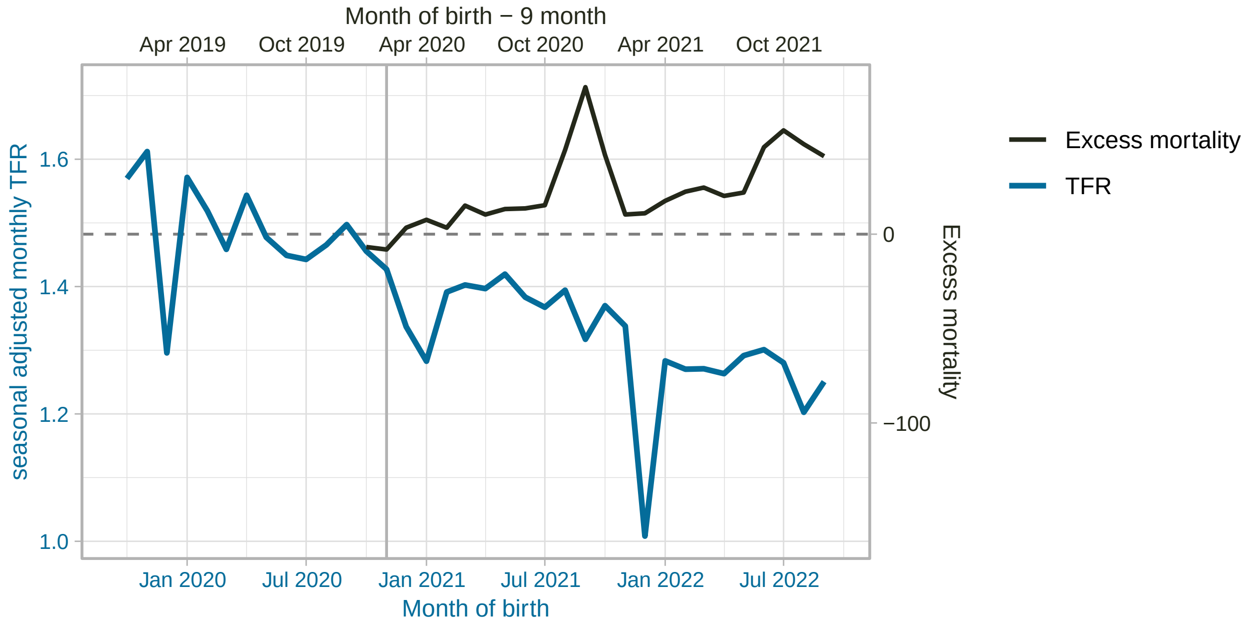


(figure continued on next page)

Latvia

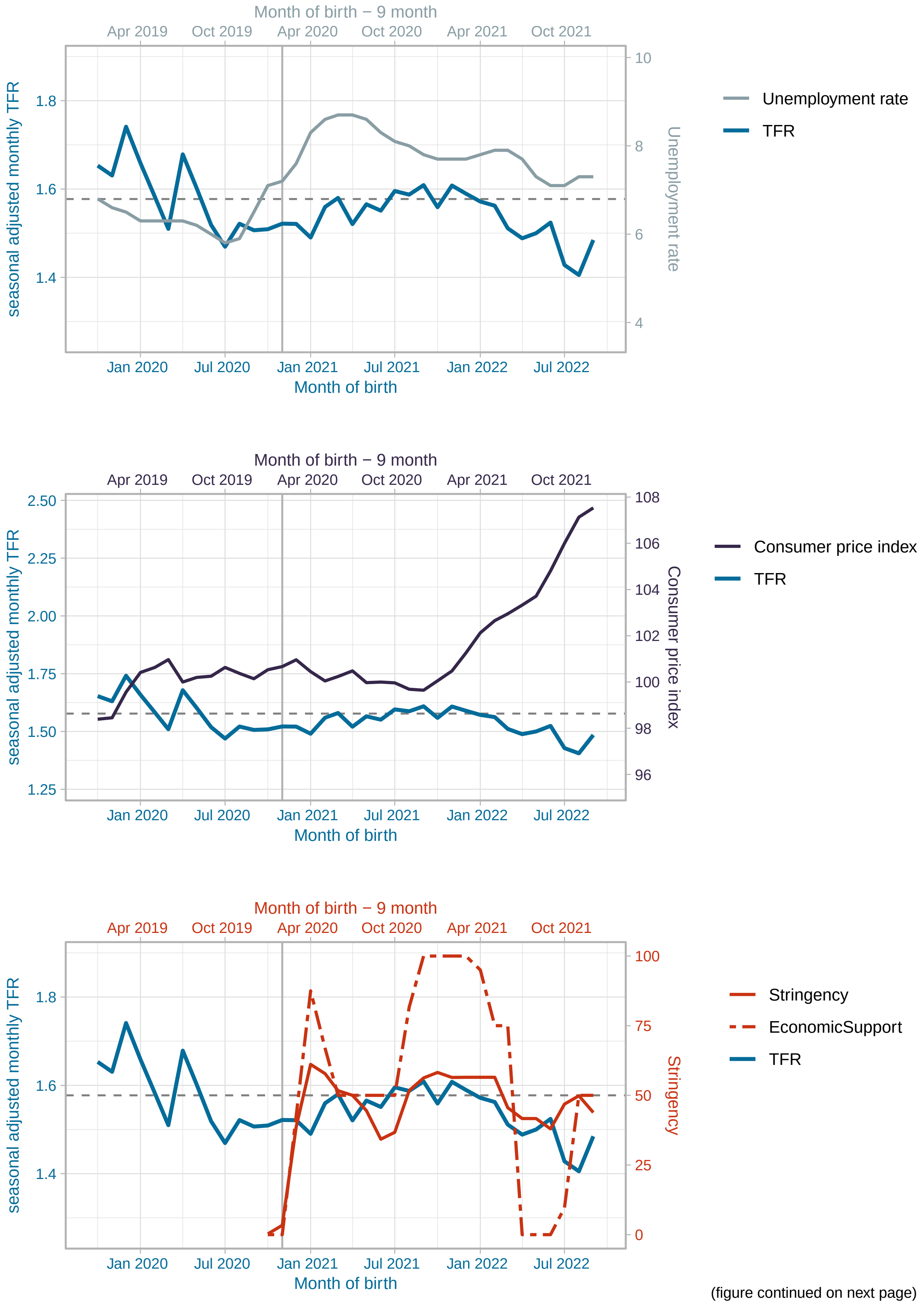


(figure continued on next page)

#### Latvia

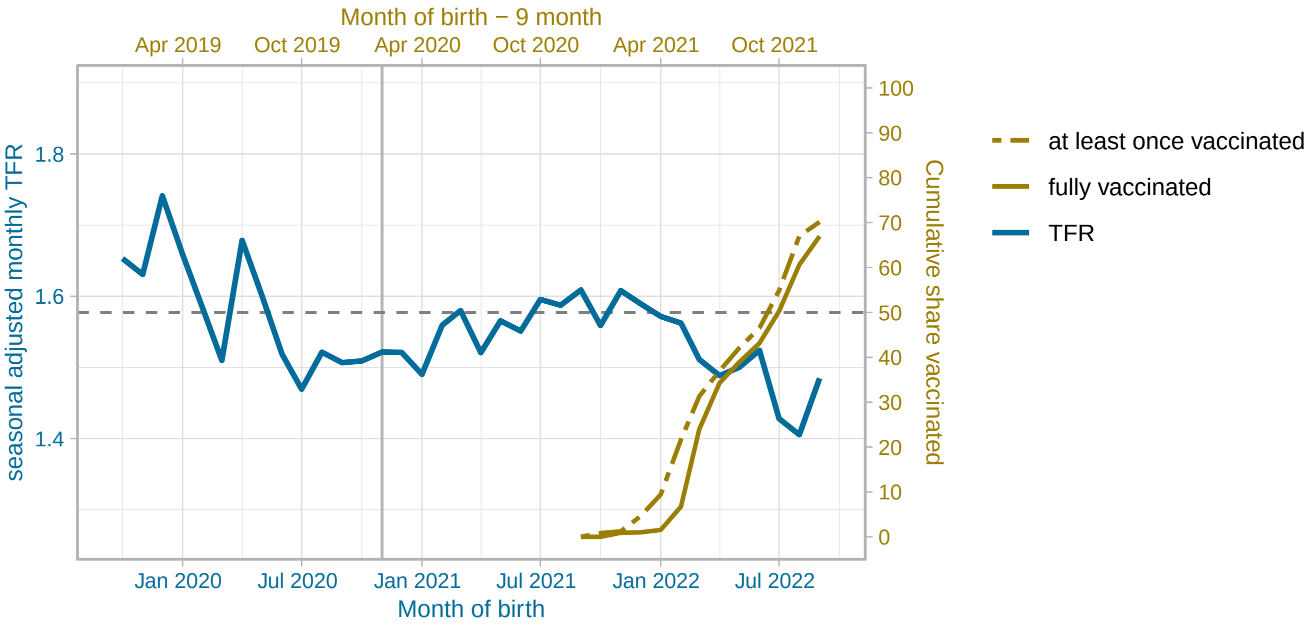


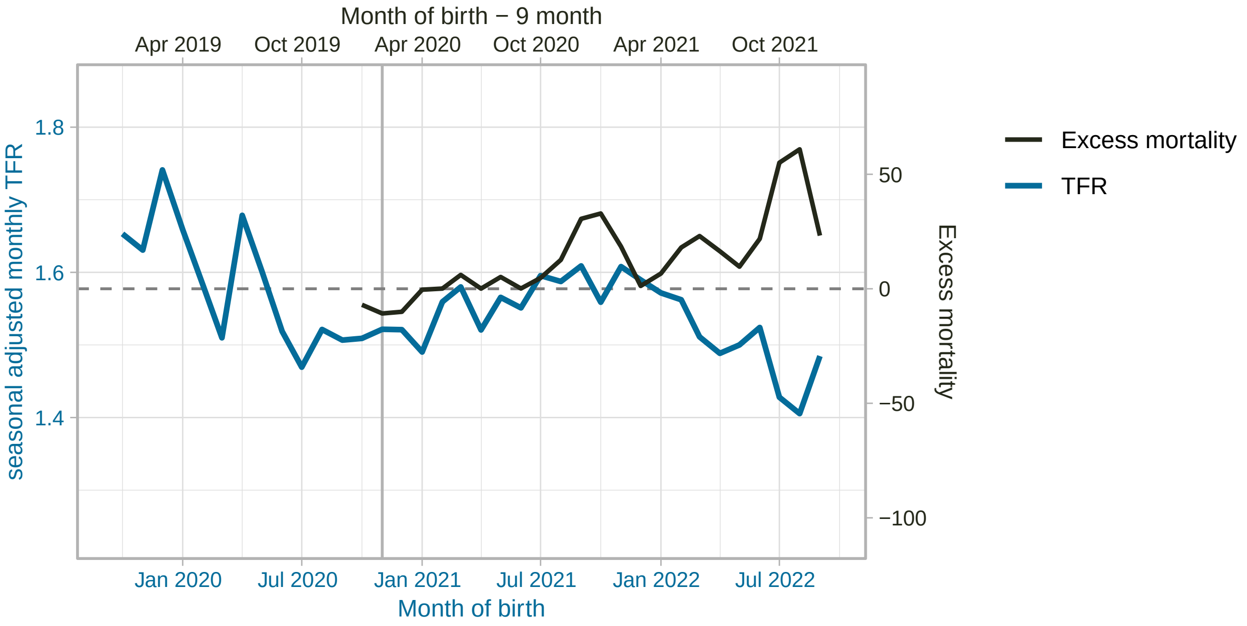


(figure continued on next page)

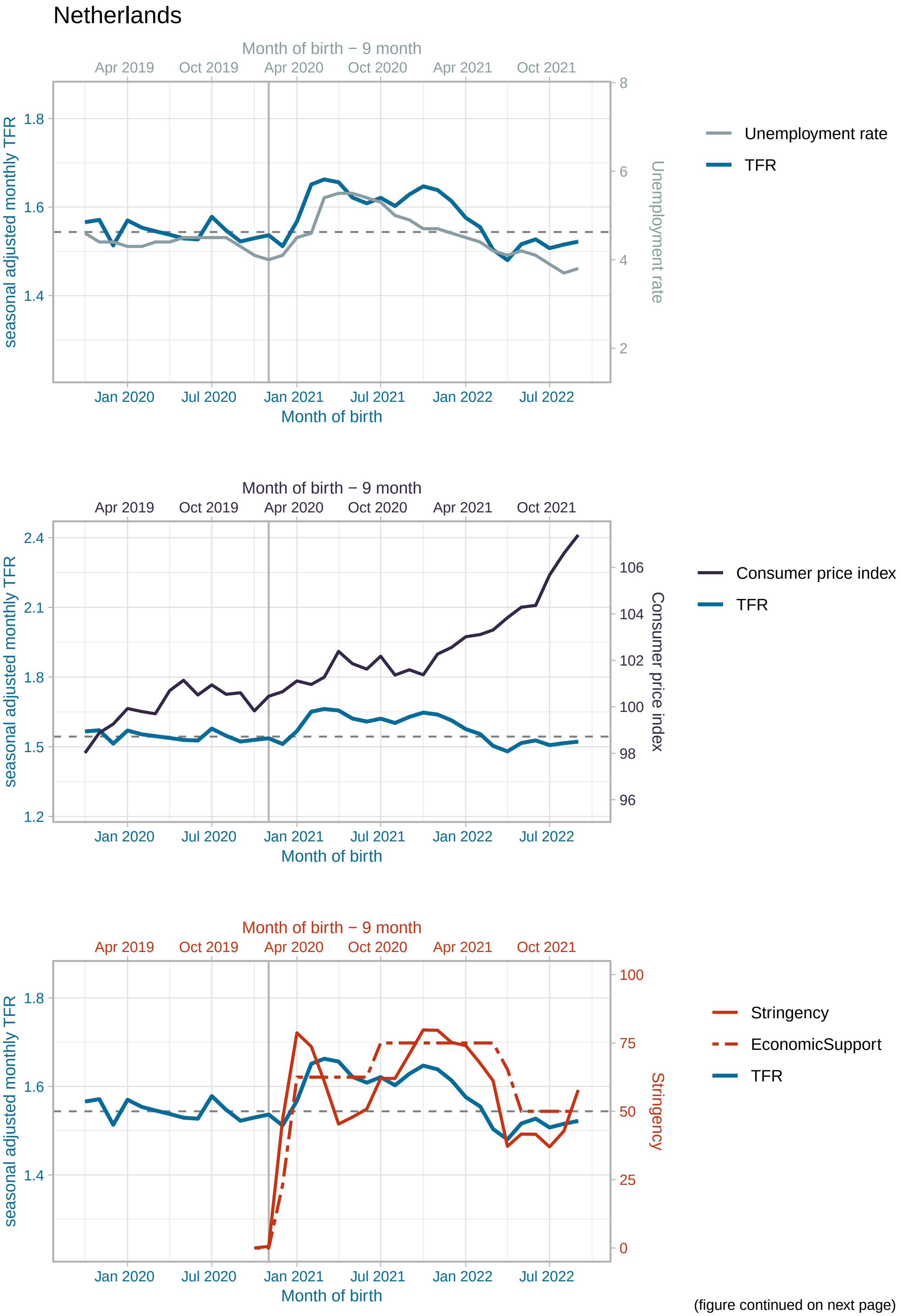


(figure continued on next page)

#### Netherlands

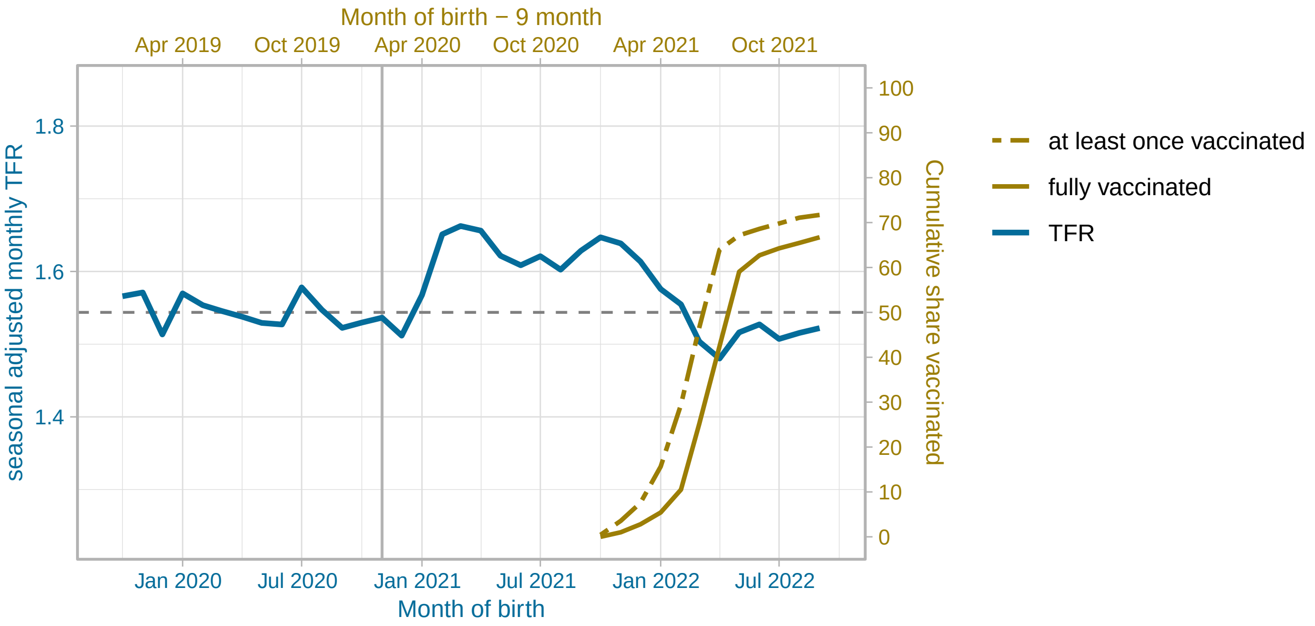


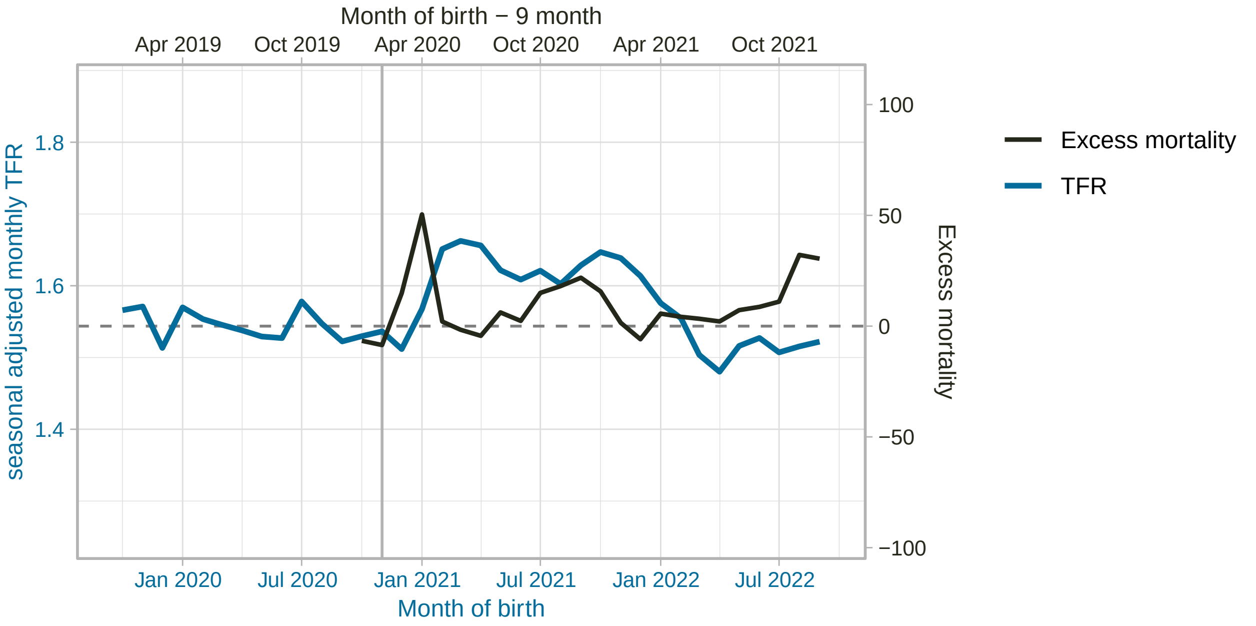


(figure continued on next page)

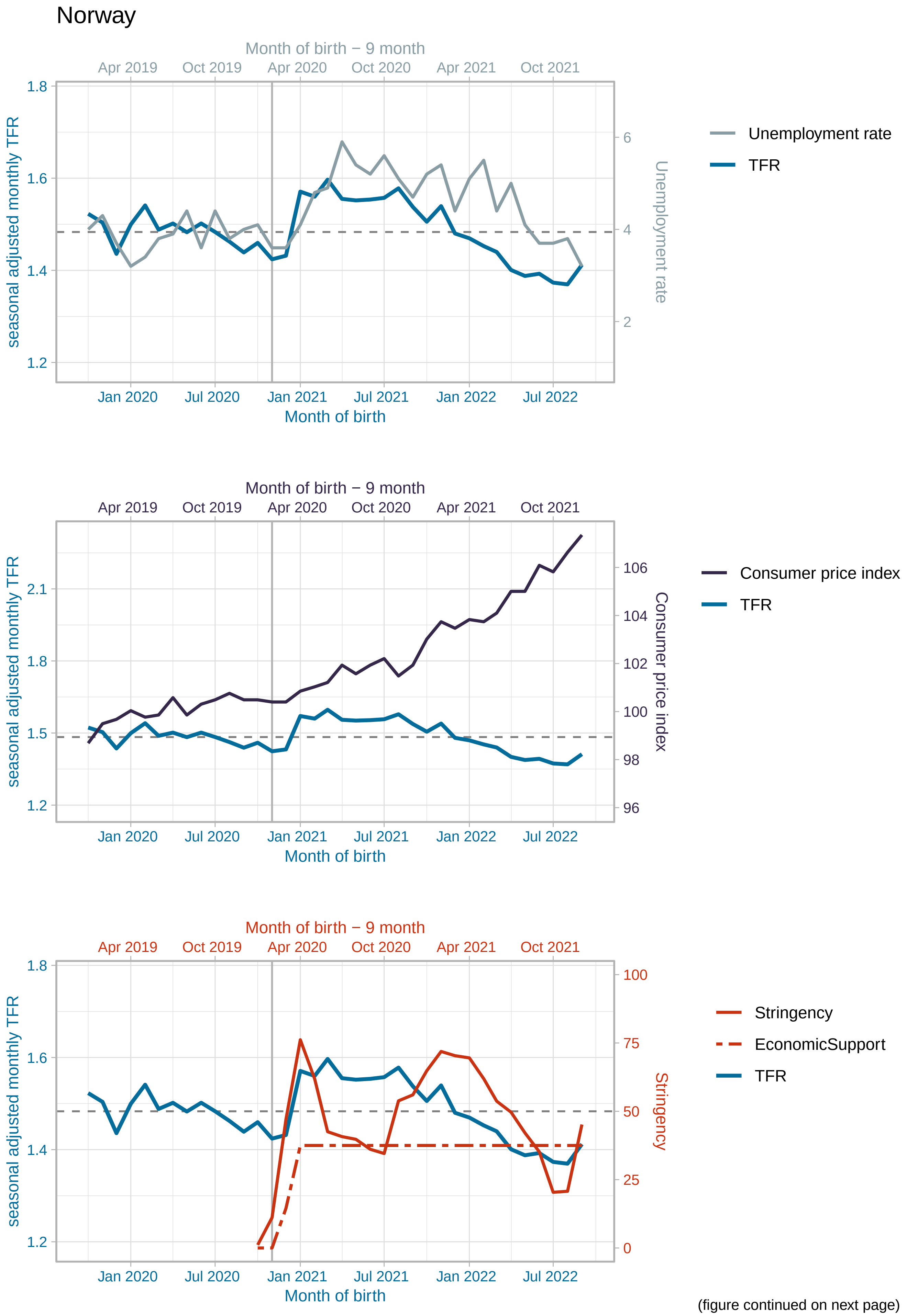


(figure continued on next page)

Norway

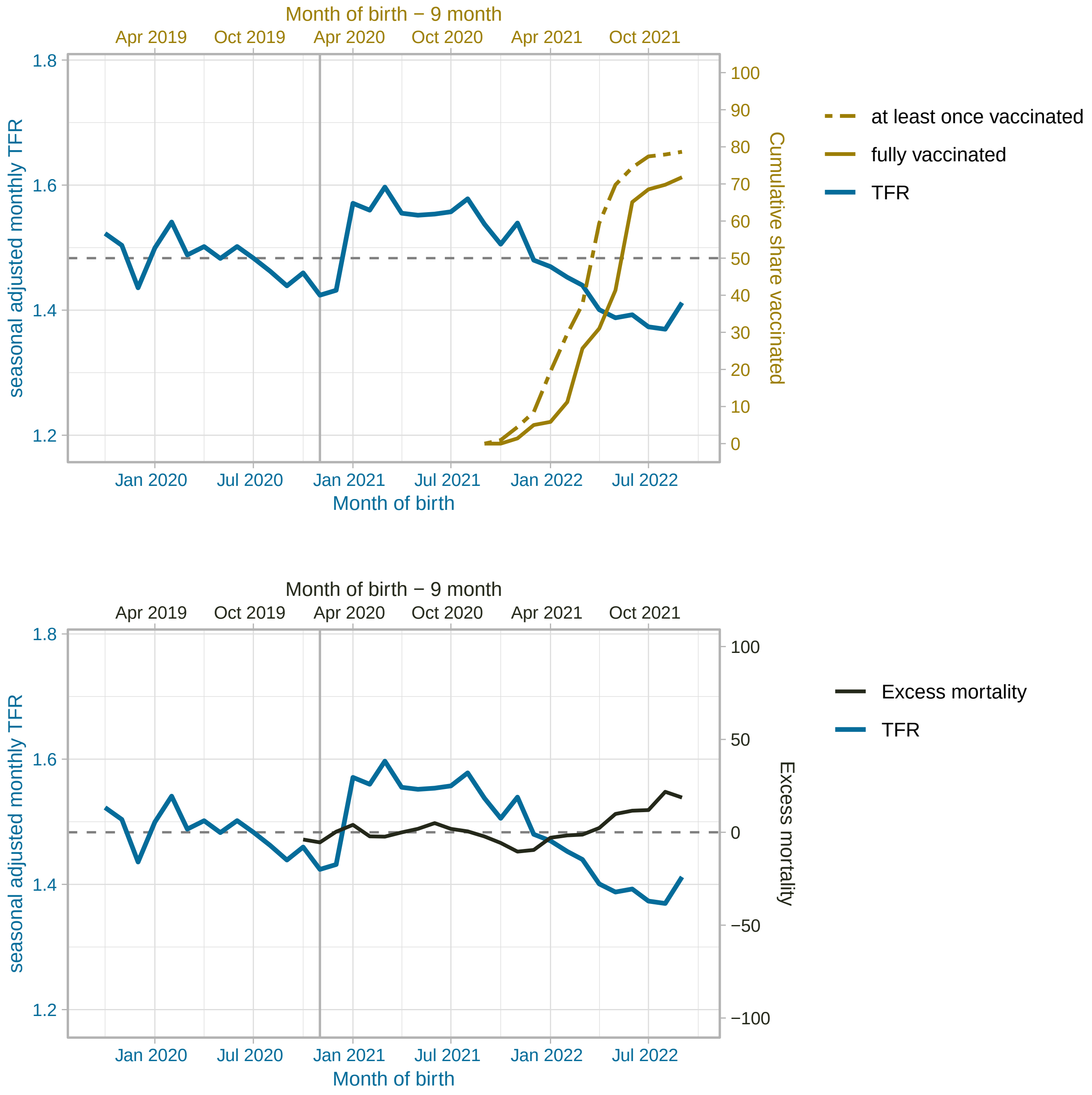


(figure continued on next page)

Poland

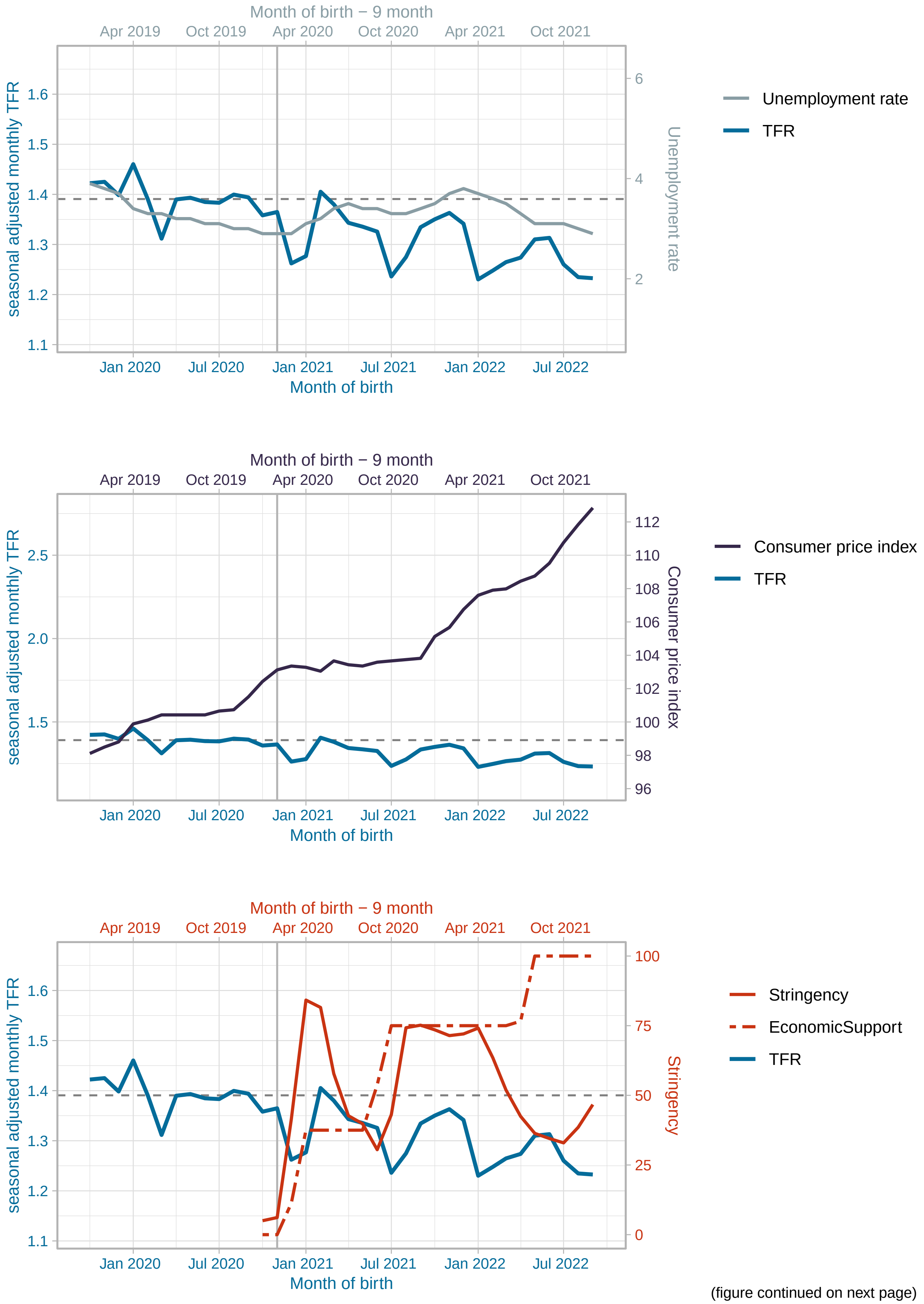


(figure continued on next page)

#### Poland

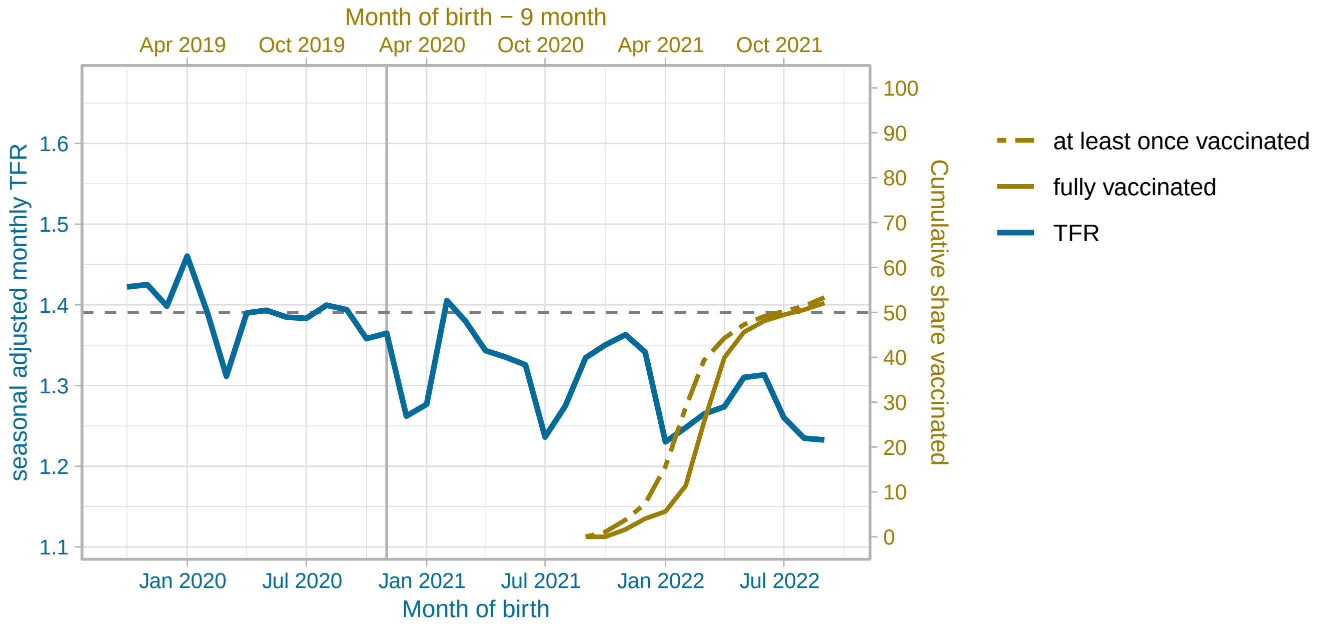


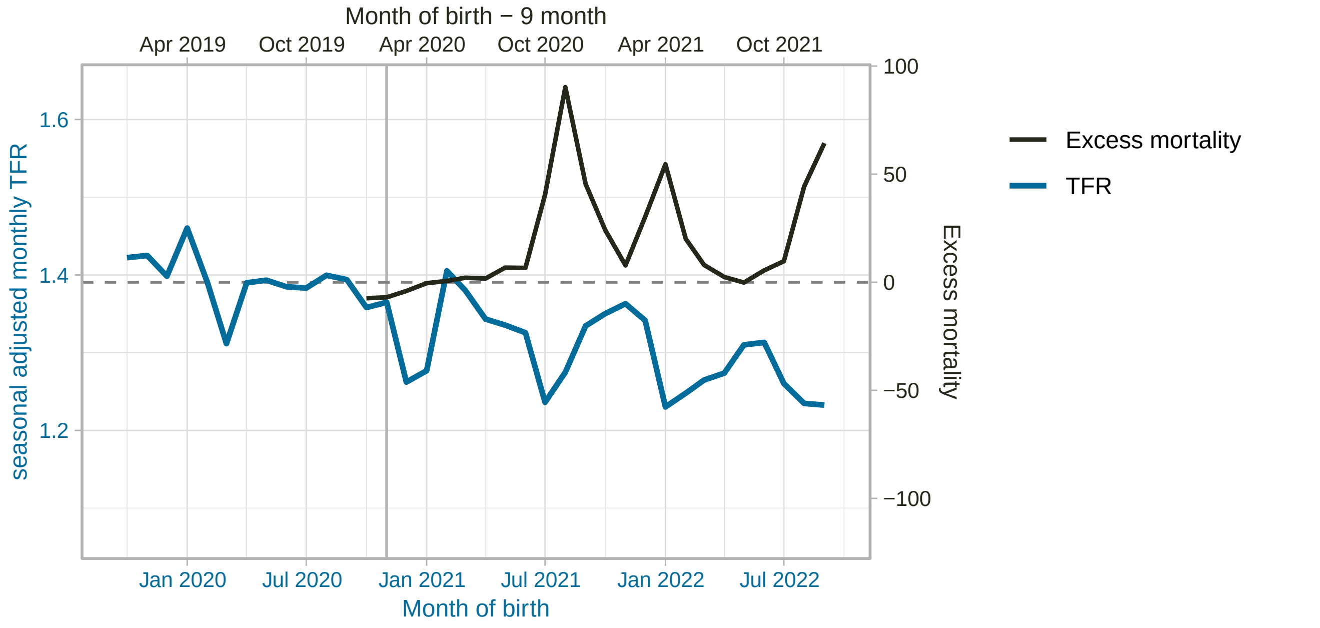


(figure continued on next page)

#### Portugal

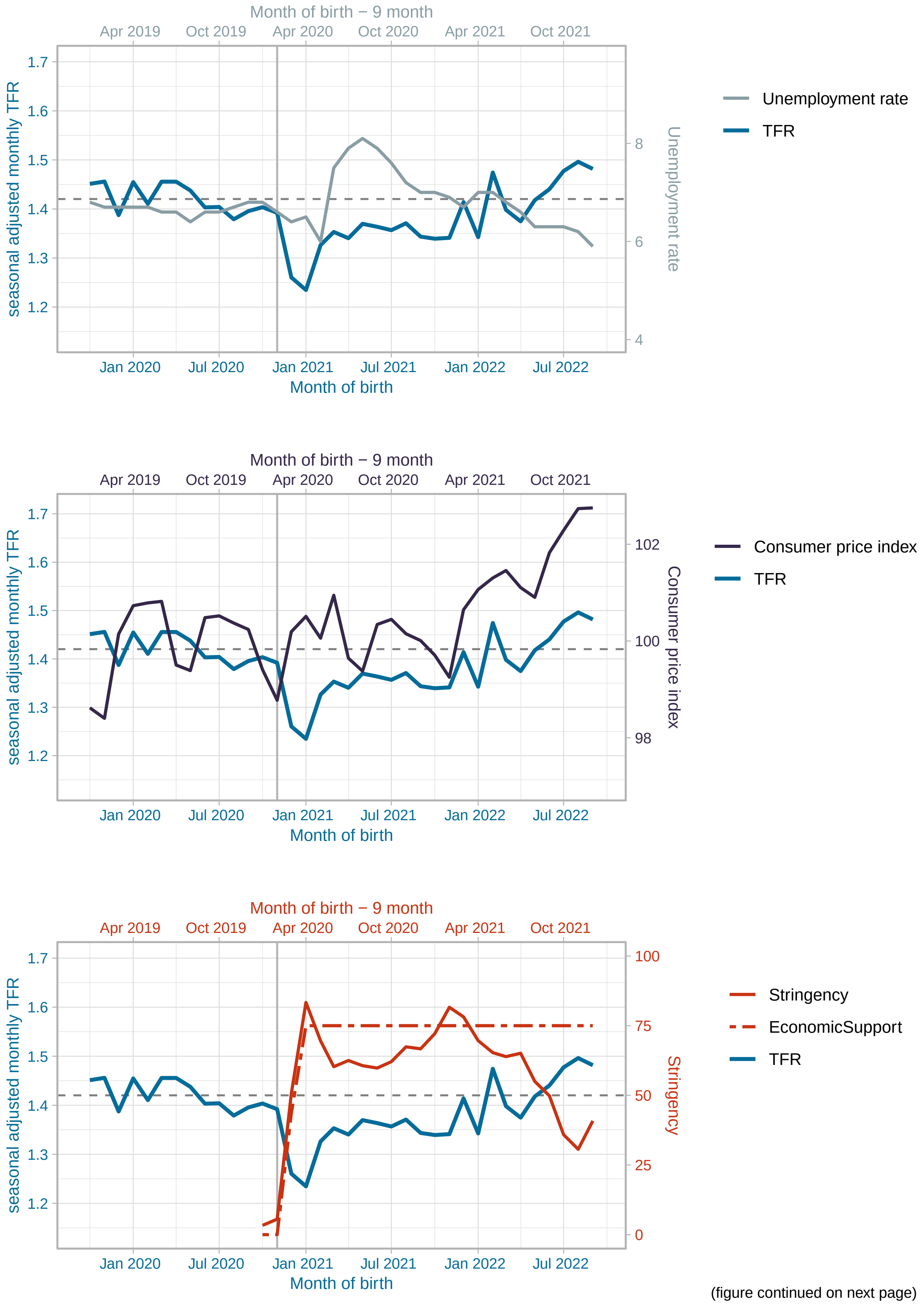


(figure continued on next page)

#### Portugal

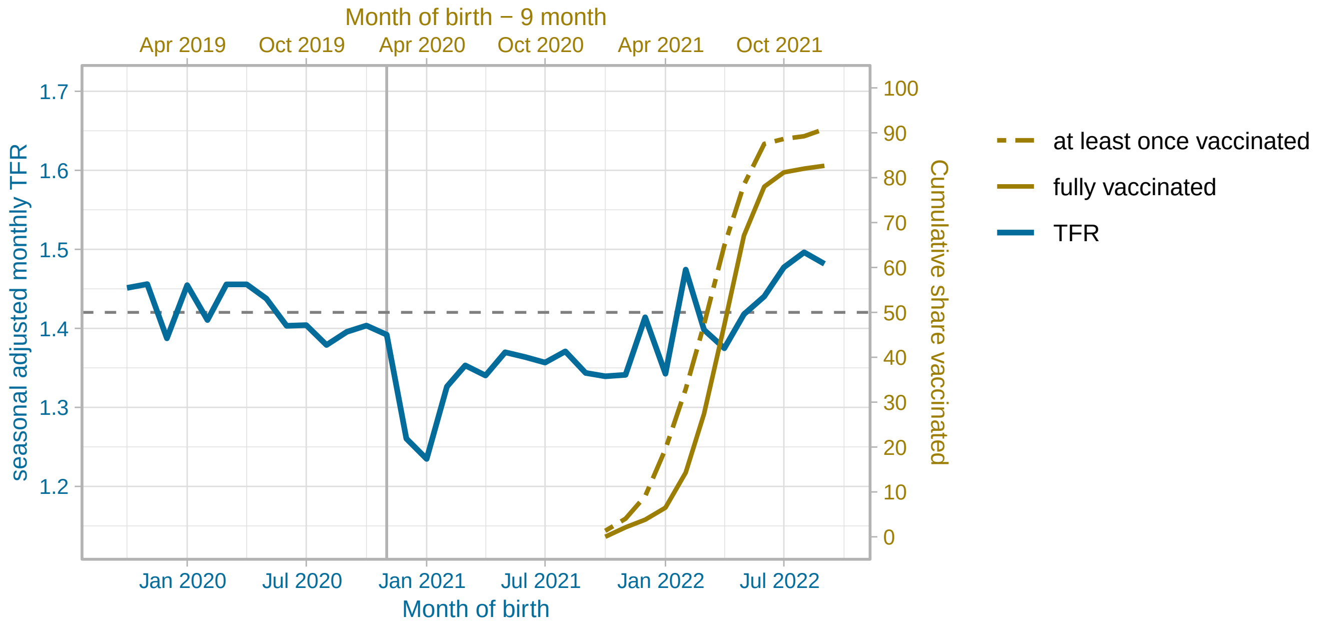


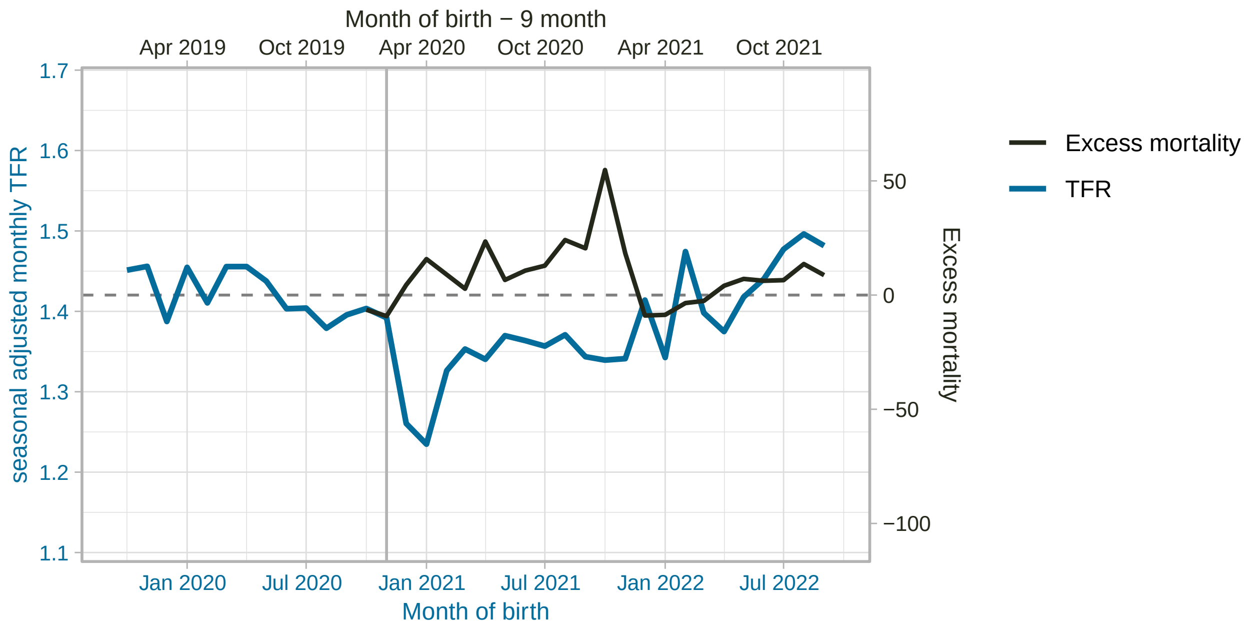


(figure continued on next page)

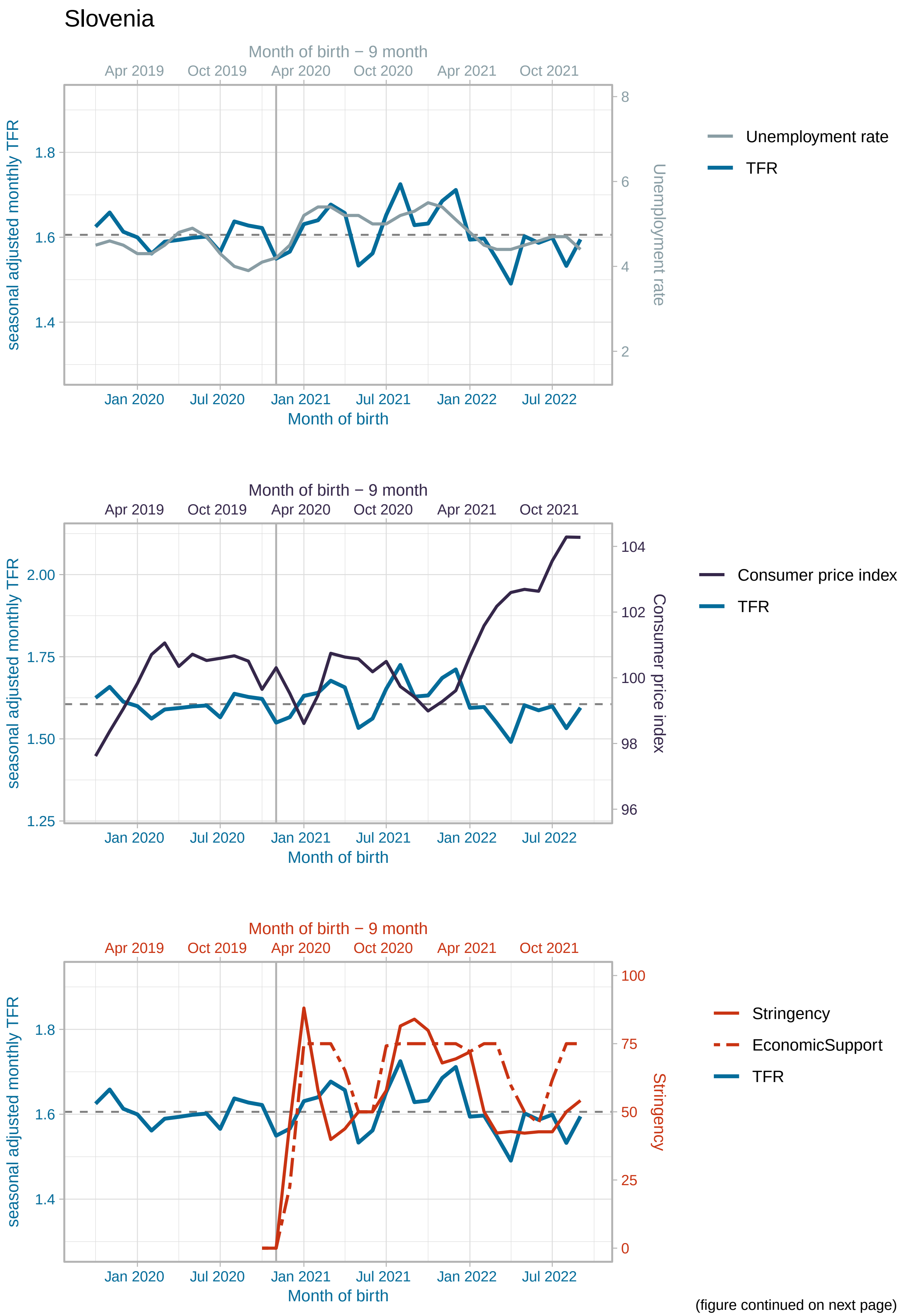


(figure continued on next page)

#### Slovenia

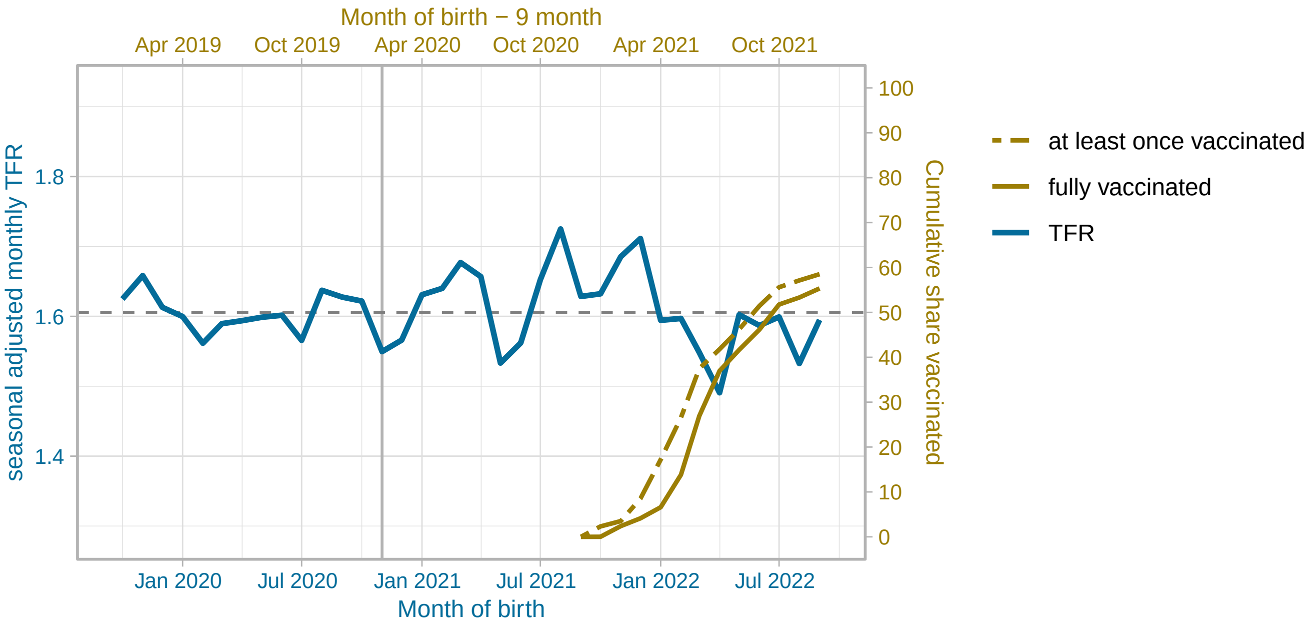


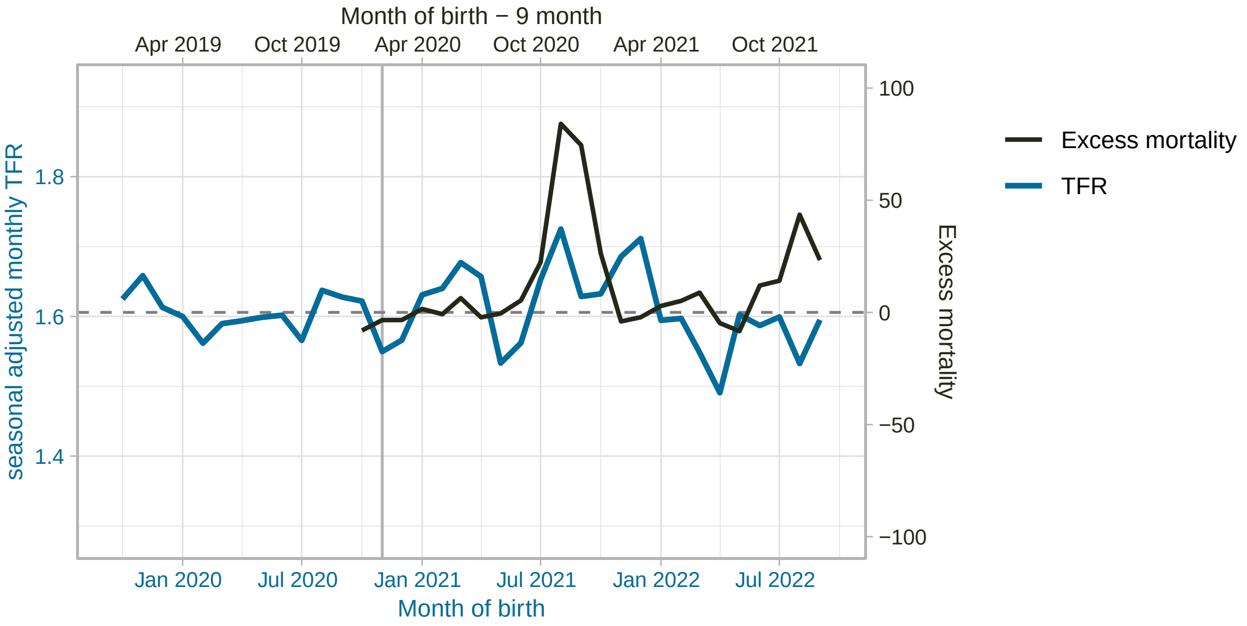


(figure continued on next page)

#### Sweden

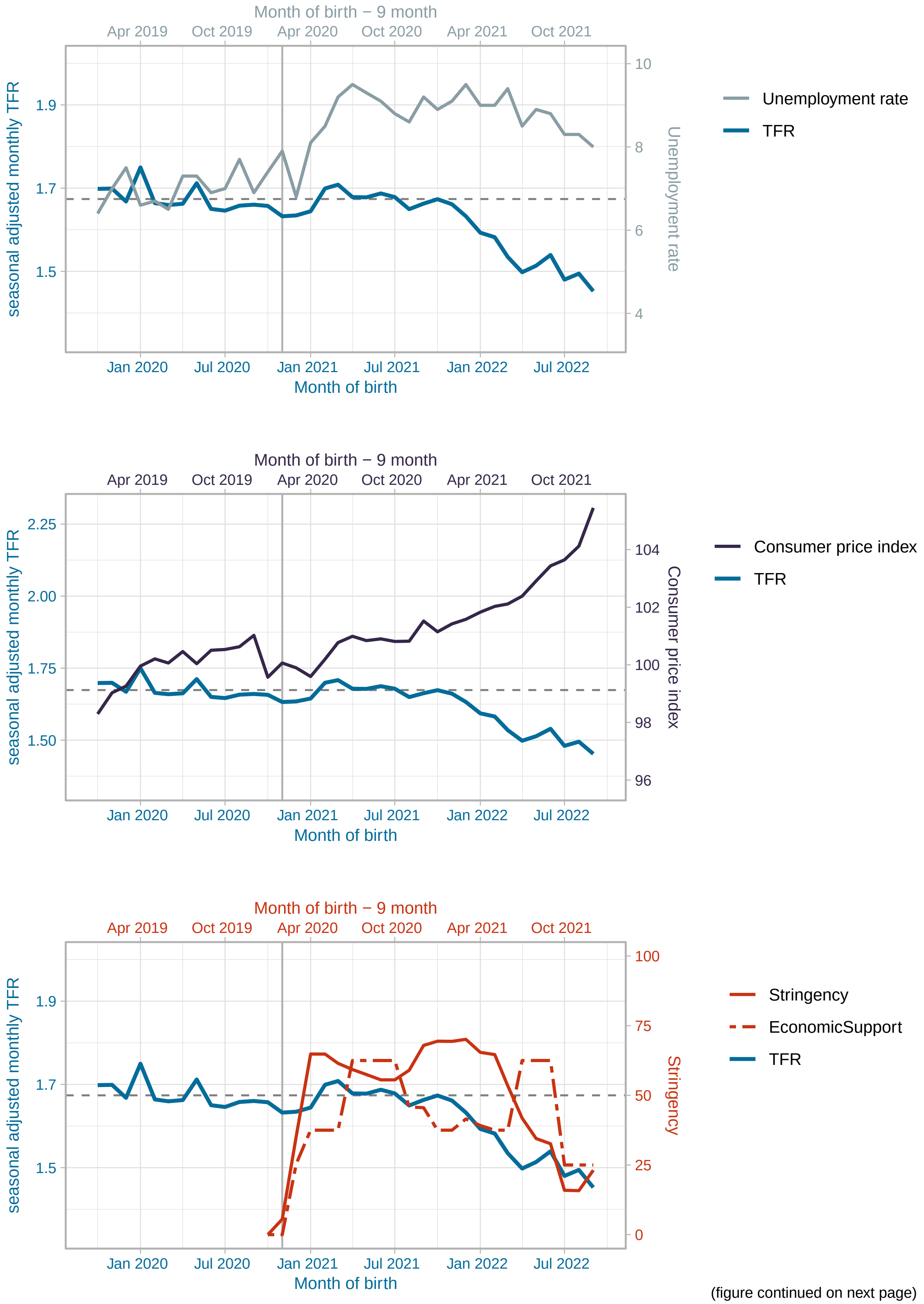


(figure continued on next page)

Sweden

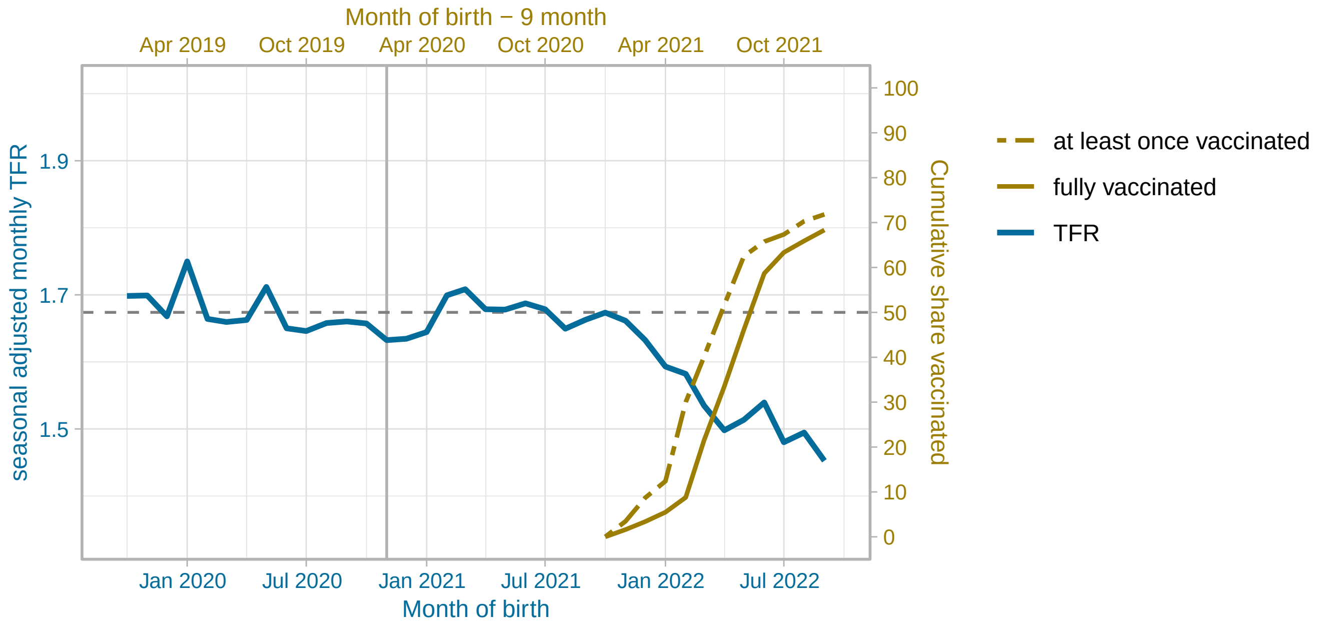


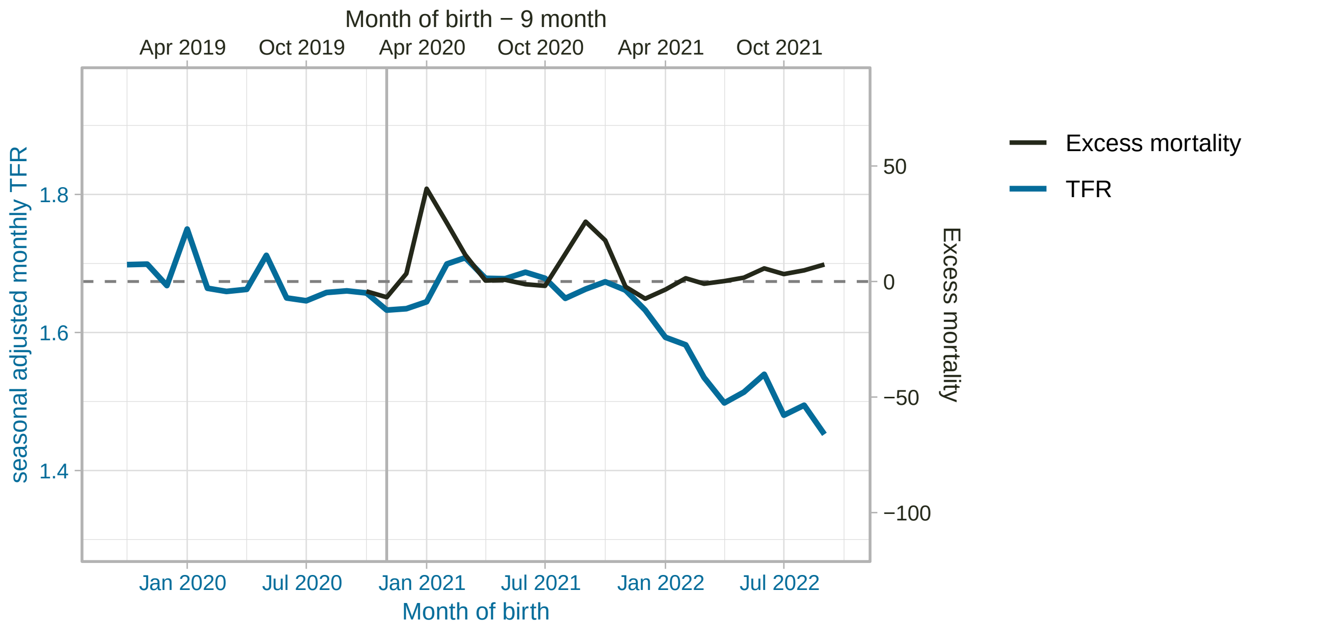


(figure continued on next page)

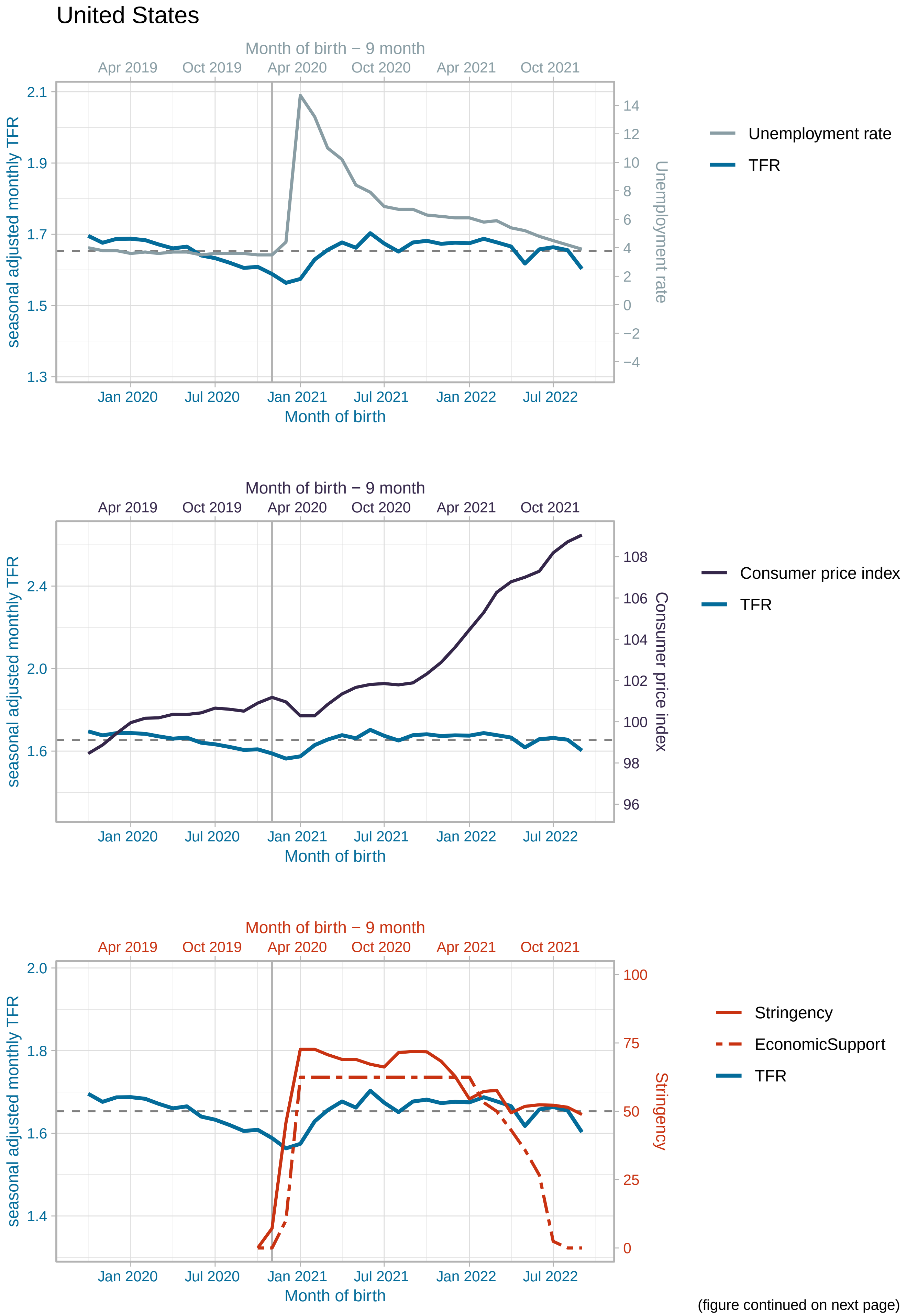


(figure continued on next page)

United States

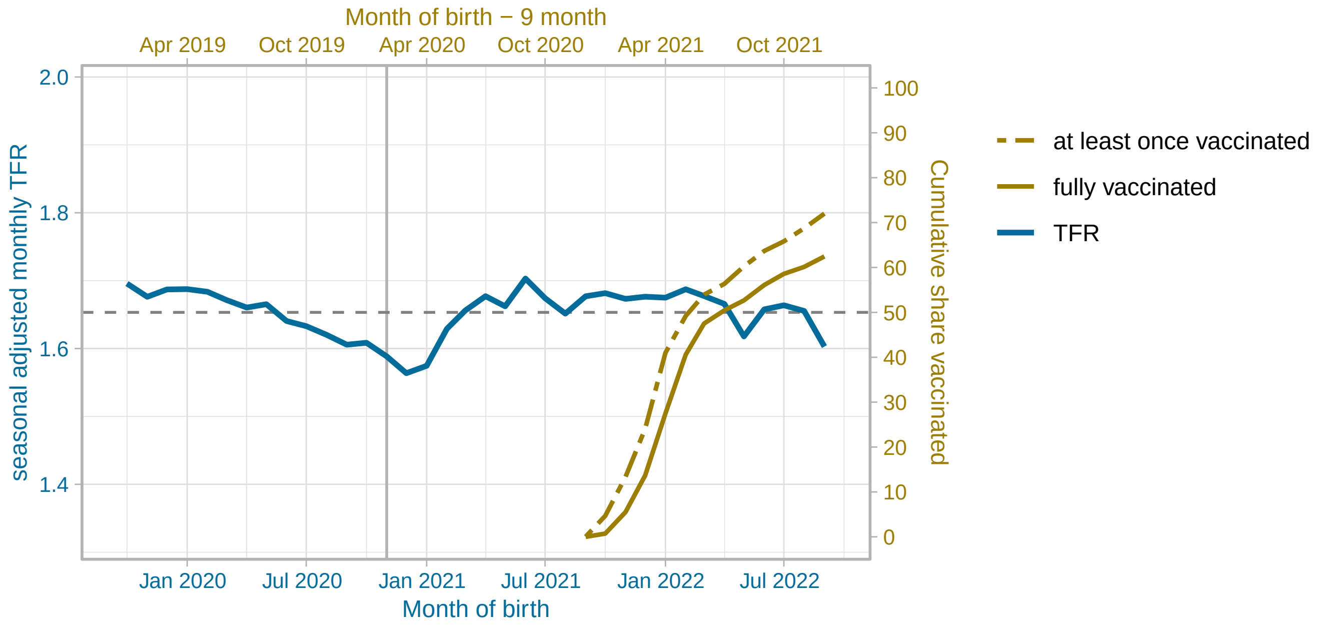


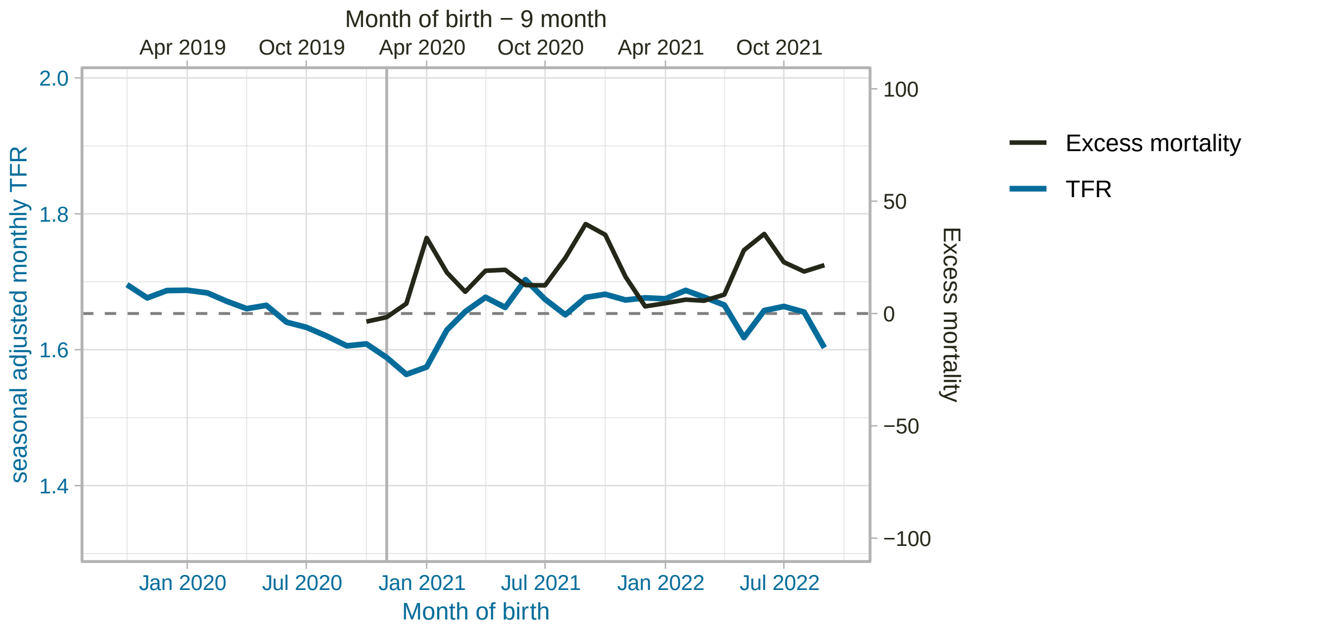
